# Change in Symptoms and Immune Response in People with Post-Acute Sequelae of SARS-Cov-2 Infection (*PASC*) After SARS-Cov-2 Vaccination

**DOI:** 10.1101/2021.07.21.21260391

**Authors:** Daisy Massey, Diana Berrent, Athena Akrami, Gina Assaf, Hannah Davis, Karen Harris, Lisa McCorkell, Aaron M Ring, Wade L Schulz, Hannah Wei, Harlan M Krumholz, Akiko Iwasaki

**Author notes:** **Address correspondence to:** Harlan M. Krumholz MD, SM, 1 Church Street, Suite 200, New Haven, CT 06510.

## Abstract

As more people are vaccinated against SARS-CoV-2, many of those already infected are still suffering from Post-Acute Sequelae (PASC). Although there is no current treatment for PASC, reports from patients that the vaccine itself improves, and in some reports, worsens, PASC symptoms may lead to a deeper understanding of the causes of PASC symptoms and viable treatments. As such, we are conducting a study that measures the changes in PASC symptoms after vaccination. We are collecting baseline self-report and biospecimens for immune assays and then are following up with participants to collect the same data at 2-weeks, 6-weeks, and 12-weeks post-vaccination (first dose). Immune assays using blood specimens will include B-cell, T-cell, and myeloid cell panels; evaluation of T-cell responsiveness to SARS-CoV-2 peptides and antigen specific response; autoantibody screening (of IgG, IgM, and IgA antibodies that attack human proteins); and TCR sequencing and antigen mapping of CD8+ T-cells. Mucosal immunity will be measured using saliva specimens. The study aims to provide answers for people with PASC, especially regarding the causes of their symptoms and how the vaccine may affect them, and clues for PASC treatment.

## BACKGROUND

With many millions of people infected with SARS-CoV-2, the world faces the prospect of immense numbers of people experiencing a chronic, post-infection disability and little information to guide clinical care. A substantial percentage of people infected with SARS-CoV-2 have persistent symptoms, with rates reported ranging from 10% to 30% and higher.^1–5^ These persistent symptoms are often debilitating, infringing on peoples’ ability to return to work and previous levels of activity.^6,7^ To date, there is no standard of care for people with suffering from Post-Acute Sequelae of SARS-CoV-2 Infection (PASC), but recent reports have noted that some people have found that their PASC symptoms decreased or disappeared after receiving at least one dose of a vaccine.^8^ The extent to which vaccination changes PASC symptoms, if at all, is unknown, as is the biological mechanism that would cause the improvement. Understanding the mechanism by which vaccination affects PASC symptoms would both provide advances in treatment and information as to how vaccination affects people with PASC.

The general purpose of the Yale COVID Recovery Study is to determine the change in immune responses in people with PASC after vaccination. The idea for the study came from an observation from Survivor Corps, a COVID-19 patient advocacy organization with over 170,000 members, many of whom have been infected with SARS-CoV-2 and are now struggling with PASC. <u>Survivor Corps</u> had been conducting research through citizen science collaborations that tracked the patient’s experience with Long COVID throughout the pandemic. With the goal of generating knowledge about people with prolonged recoveries, they posted a poll to their patient community about the effects of vaccination on PASC symptoms. The poll found that about 40% of people reported mild to full resolution of their symptoms after they were vaccinated while about 14% of people reported worsening of their symptoms after they were vaccinated. In response to this finding, Dr. Akiko Iwasaki independently developed and published hypotheses to explain how the vaccine might affect people with PASC and what might be causing PASC in the individuals for whom vaccination improves their symptoms.^9,10^ Conversations between Survivor Corps and Yale investigations then led to the decision to pursue the study.

We hypothesize that people with PASC exhibit heterogeneity in immune profile and viral reservoir, and that among people with PASC who are vaccinated, changes in immune parameters correlate with symptom response. Specifically, we hypothesize that those who have autoreactive T cells benefit from effector function diversion by vaccine-induced cytokines, while those with a viral reservoir may benefit from its elimination by vaccine-induced antiviral antibodies and T cells. Moreover, there is some evidence that women more often have a profile of more robust T cell immunity and men might be more likely to maintain a viral reservoir.^11^ Accordingly, we will investigate differences in the immune profile and viral load (and its surrogates) of people with PASC and compare with those immune profiles in people without PASC but evidence of prior infection. We will also study changes in symptoms and immune profiles after vaccination for people with PASC. We specifically will determine whether changes occur through antigen-specific adaptive immune stimulation or innate immune signals leading to transient immunomodulation and whether there are sex differences. Thus, the objective of this study is to determine changes in clinical and immune responses to vaccination and their correlation, using both participant self-report through surveys and immune analyses from biospecimen samples provided by participants.

Accordingly, we are conducting a study that combines digital participant self-report to characterize participants’ symptoms and assays to characterize participants’ immune profiles. We will characterize how the changes in symptoms correspond to changes in aspects of the immune response. We will enroll participants with moderate-to-severe PASC prior to vaccination so that we may measure their immune and symptom profiles both before and after vaccination.

## METHODS

### Design Overview

We are conducting an observational study of people aged 12 and over who have moderate-to-severe PASC and who are planning to receive a vaccine against SARS-CoV-2. The study seeks primarily to describe the heterogeneity in symptoms and immune profile among people with PASC, and once participants have been vaccinated, to determine changes in clinical and immune responses to vaccination and their correlation.

The study will include 4 surveys to monitor participants’ symptom profile and experience with COVID-19 as well as 3 separate biospecimen collection appointments. The surveys and study enrollment will be conducted through REDCap at Yale, a platform that is designed to securely collect survey data from people in a user-friendly design setting. The biospecimen collection will occur as follows: the first will occur as soon as a participant is enrolled, the second will occur 6 weeks after the participant receives the first dose of a COVID-19 vaccine, and the third will occur 12 weeks after the participant receives the first dose of a COVID-19 vaccine. This study will be conducted both online and in person for biospecimen collection.

The engagement required by each participant will last approximately 15 weeks, depending on when the participant receives the first dose of a COVID-19 vaccine.

The study protocol has been reviewed and approved by the Institutional Review Board at the Yale School of Medicine.

### Study Population

The study inclusion and exclusion criteria are listed in **Table 1**. Study participants will include people aged 12 and over who either have been diagnosed with COVID-19, hospitalized due to COVID-19, or who have tested positive for SARS-CoV-2 more than 2 months prior to joining the study (PCR, antigen, or a positive T-test or antibody test at any point), and who have proof of the diagnosis, hospitalization, or positive test; who intend to receive the vaccine for SARS-CoV-2, but who have not yet received any doses of the vaccine; and who self-report that they have persistent moderate-to-severe symptoms or disability (by answering “yes” to the question: In your opinion, do you have symptoms more than 2 months after your initial COVID-19 infection (Long COVID) that interfere with your quality of life or with your ability to do your normal activities?). Additionally, they must be fluent in English or Spanish and have the ability and willingness to participate in a digital study and give blood samples at a Yale site 3 times on a Monday or Wednesday morning (before 12 pm). Minors (ages 12-17) are eligible for the study, but will require a parent or guardian to also consent to the study and be present for all study visits.

**Table 1.** Study inclusion and exclusion criteria

|  |  |  |
| --- | --- | --- |
| Inclusion criteria | ☐ | Fluent in English or Spanish |
|  | ☐ | Age 12 years and over (if ages 12-17, must have a parent or guardian join with participant) |
|  | ☐ | Test positive for SARS-CoV-2 (defined as a patient who reports receiving any screening or diagnostic test used to detect the presence of SARS-CoV-2 including any FDA-approved or authorized molecular or antigen-based assay) at least 2 months prior to the start of this study OR test positive for antibodies to SARS-CoV-2 at any point OR receive a positive T-test at any point OR receive a clinical diagnoses of COVID-19 at any point OR was hospitalized due to COVID-19 at any point |
|  | ☐ | Self-report moderate to severe symptoms or disability due to PASC |
|  | ☐ | Self-report planning to receive a vaccine against SARS-CoV-2 |
|  | ☐ | Self-report has not yet received any doses of a vaccine against SARS-CoV-2 |
| Exclusion criteria | ☐ | Unable to provide informed consent |
|  | ☐ | Unable to provide 3 blood and saliva samples at one of the 2 participating Yale New Haven Health blood draw sites on a Monday or Wednesday morning (before 12 pm) |
|  | ☐ | Does not have access to a hand-held device or computer that would allow for digital participation in the study |
|  | ☐ | Do not have and do not want to set up an email address |
|  | ☐ | Individuals who are prisoners |

### Recruitment

Potential participants will be provided a link for the study enrollment as well as an email address where they can reach out for more information about the study. Participants will complete a brief screening questionnaire (Appendix 2) to determine study eligibility and to provide their email address and/or phone number. Study coordinators will then reach out, by email to schedule a phone consultation, or directly by phone, as is the participant’s preference. During the phone consultation, a study coordinator will read a script designed to inform the participant of the study design and risks to ensure that the participant understands the study prior to consent. The participant will also be asked if they have any questions for the study coordinator. Once the phone consultation is complete, the study coordinator will email the participant a link to the e-consent (Appendix 1). Once the participant consents to the study, which will require an e-signature, an email will automatically be sent to the participant containing the link for the first survey (Appendix 3) and information as to how to provide the first biospecimen sample. We will also email participants a copy of their completed consent form as well as contact information for the study team should they have any questions or issues throughout the study. The research team will also contact the participant within the next 2-3 days by phone to provide information regarding the timing and location for biospecimen collection and to answer any questions the participant has. Participants will provide blood and saliva samples 3 times.

The study has expectations for participants and academic researchers.

i. *<u>Participants:</u>* Participants will agree to contribute to knowledge generation by filling out 4 surveys and providing biospecimens 3 times at Yale draw sites in a timely manner. Participants will share their ideas, wisdom, and suggestions with researchers, including by responding to open-ended survey questions as they feel appropriate. They will freely express any concerns and contribute to continually improving the study. They will ask any questions, knowing their comments are welcome and they are valued members of the team.
ii. *<u>Academic Researchers</u>*: Researchers will treat all participants as teammates and protect their data and privacy. Researchers will work hard, in partnership with participants, to produce research that will improve people’s health.

Diversity Goal: The goal is that a minimum of 25% of the participants are from Black and Latinx/Hispanic populations.

### Team Involvement

This research team is committed to involving patient communities in the study design and the distribution of results to participants. As noted above, the early observation by <u>Survivor Corps</u> that some people with Long COVID improved after vaccination stimulated the development of a set of hypotheses that ultimately became the basis of the COVID Vaccine Recovery study. In addition, Survivor Corps generated press coverage aimed at driving enrollment and interest in the study. Survivor Corps advocated for including those without positive PCR tests for SARS-CoV-2. They also reviewed and contributed to the surveys and advocated for asking when symptoms began by start date instead of by length in weeks.

We also worked with the <u>Patient-Led Research Collaborative</u>, a self-organized group of Long COVID patient-researchers working on patient-led research around the Long COVID experience. The Yale team had been engaged with the Patient-led Research Collaborative in the development of a set of surveys for an observational study of Long COVID and strategies to promote patient engagement when the idea for this study emerged. The survey that was in development was well positioned to be used for this study, so that effort was re-positioned for this study. The group then iteratively contributed to the surveys for this study. They also raised the issue of including people without positive PCR tests for SARS-CoV-2. The Patient-Led Research Collaborative was paid for their contributions.

This study’s surveys also drew on previous survey studies.^6,12–16^

The academic researchers developed the study hypotheses, and were responsible for study oversight, management, execution and funding. The academic researchers are also responsible for the laboratory testing. The academic researchers were also responsible for the IRB approval – and will be responsible for leading the academic papers.

### Data Collection Overview

Data collected will include participant self-report survey data and biospecimens (blood and saliva). The overall study flow has been described in **Figure 1**.

**Figure 1.**
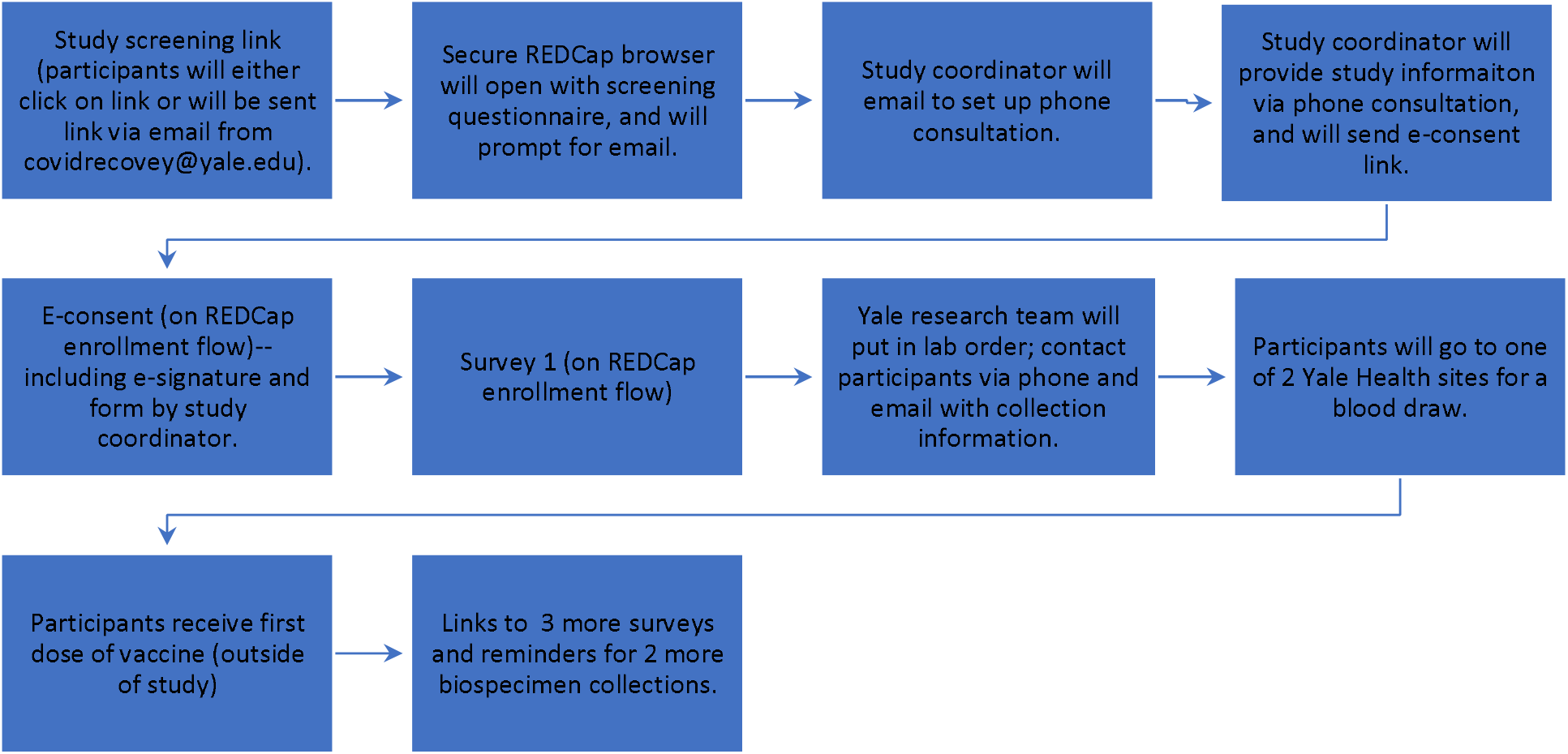
Flow chart of study. (Describe the key partners and their roles in enrollment, data collection, data cleaning and analysis).

#### Participant Surveys

The study will include 4 surveys to describe participants’ experience with PASC. The 4 main surveys delivered to participants will be delivered on a timetable based on vaccination: Survey 1 will be delivered upon enrollment, prior to participants receiving any vaccine doses; Survey 2 will be delivered 2 weeks post-vaccine (first dose); Survey 3 will be delivered 6 weeks post-vaccine (first dose); and Survey 4 will be delivered 12 weeks post-vaccine (first dose).

Survey 1, delivered upon study enrollment, will solicit basic demographic information, including participant’s zip code, gender, and race/ethnicity. Survey 1 will also solicit information about the participant’s symptoms due to COVID-19 and the severity, timing, and waxing or waning of the symptoms; health status; and SARS-CoV-2 testing. Survey 1 is estimated to take 15-20 minutes. Survey 1 can be found in Appendix 3.

Surveys 2, 3, and 4 will be delivered 2, 6, and 12 weeks after a participant receives their first vaccine dose and will solicit information regarding the participant’s symptoms due to COVID-19 and the severity, timing, and changes of the symptoms; and health status.

Surveys 2, 3, and 4 are estimated to take 10-15 minutes each. Surveys 2, 3, and 4 can be found in Appendices 4, 5, and 6, respectively.

All surveys will also include an open-ended response option to capture information that the participant deems essential, but that the survey did not capture.

Follow-up reminders will be sent at 48 hours, if the survey is not completed.

The surveys were created using patient input. Due to the novelty of PASC and the lack of definition surrounding the condition and its symptoms, we did not use validated questions in our surveys. Instead, to generate the list of relevant medical conditions and PASC symptoms, we created a combined list of PASC symptoms by starting with symptoms listed by the Johns Hopkins Long COVID Survey,^16^ and then adding PASC symptoms identified through survey research with people with PASC^6,7^ to include symptoms not yet captured by studies of people with PASC who were hospitalized or outpatient, since many people with PASC do not receive medical care. Once the combined lists of medical conditions and symptoms were created, we solicited patient feedback by sharing the survey drafts with five members of Patient-Led Research Collaborative, a member of Survivor Corps, and two individual patients, all of whom were people with PASC.

#### Blood and Saliva Collection

The biospecimen collection will occur as follows: the first will occur as soon as a participant is enrolled, the second will occur 6 weeks after the participant receives the first dose of a COVID-19 vaccine, and the third will occur 12 weeks after the participant receives the first dose of a COVID-19 vaccine. Participants will be asked to provide 45 mL of blood and a saliva sample (up to 5 mL) for each biospecimen collection. Participants will be able to give a blood sample within a 2-week window (e.g. the collection may be within 5-7 weeks after their first COVID-19 vaccine dose). The survey collection will occur as follows: the first will occur as soon as the participant is enrolled, the second will occur 2 weeks after the participant receives the first dose of a COVID-19 vaccine, the third will occur 6 weeks after the participant receives the first dose of a COVID-19 vaccine, and the fourth will occur 12 weeks after the participant receives the first dose of COVID-19 vaccine. Survey links and reminders will be sent via text and/or email.

The biospecimen collection will require 45cc of blood and a saliva sample (5 mL) to be given 3 times by each participant. The blood sample will be collected by a trained technician at a Yale New Haven Health blood draw site. The blood sample will immediately undergo mild agitation and remain stored under controlled temperature. Saliva samples will be self-collected by the participant, who will receive instructions upon arriving at the draw site.

### Data Management

We will assemble an advisory group of some participants and the researchers for guidance on data collection, data analysis and interpretation, and results dissemination. The PIs will form the decision-making group for the study.

Data for this study will be collected, recorded, and stored using REDCap, a secure web application designed to support data capture for research studies. It includes features for HIPAA compliance including real-time data entry validation (e.g. for data types and range checks), a full audit trail, user-based privileges, de-identified data export mechanism to statistical packages (SPSS, SAS, Stata and R), and integration with the institutional Active Directory.

Only approved Yale investigators will have access to the open responses qualitative survey data saved in YNHH/CORE-administered servers. Participants can download their survey responses or request to receive them in an encrypted email. These team members will use Yale owned hardware to access PHI and PII data held in YNHH/CORE secured servers. These data will be deidentified by removing all PII. Additionally, REDCap survey data will be deidentified prior to being exported for use in analyses.

All biospecimens will be collected at Yale New Haven Health blood draw sites. Biospecimens will be stored in the Iwasaki biorepository, and plasma extracted from biospecimen samples will be shared with the Ring biorepository. Serum will be isolated after centrifugation and will be stored at −80C. PBMCs will be isolated and either used immediately and/or frozen and stored at −80C. Plasma will be stored at −80C.

The principal investigators are responsible for monitoring the data, assuring protocol compliance, and conducting the safety reviews at the specified frequency regularly. During the review process the principal investigators will evaluate whether the study should continue unchanged, require modification/amendment, or close to enrollment.

### Data Elements

The data elements have been summarized in Table 2. Data outlined in Appendices 3-6 below details the types of information that participants will share with research sites using REDCap. In addition, participants will provide 45 mL of blood specimens 3 times and saliva samples (up to 5 mL in a sterile self-collection cup), which will be used to understand participants’ immune profiles. The blood collection tubes will be as follows: 2 gold topped 5 mL serum tubes (BD), 3 green topped 10 mL plasma heparin-coated tubes (BD), and 1 lavender topped 3-5 mL EDTA-coated tube.

**Table 2.**
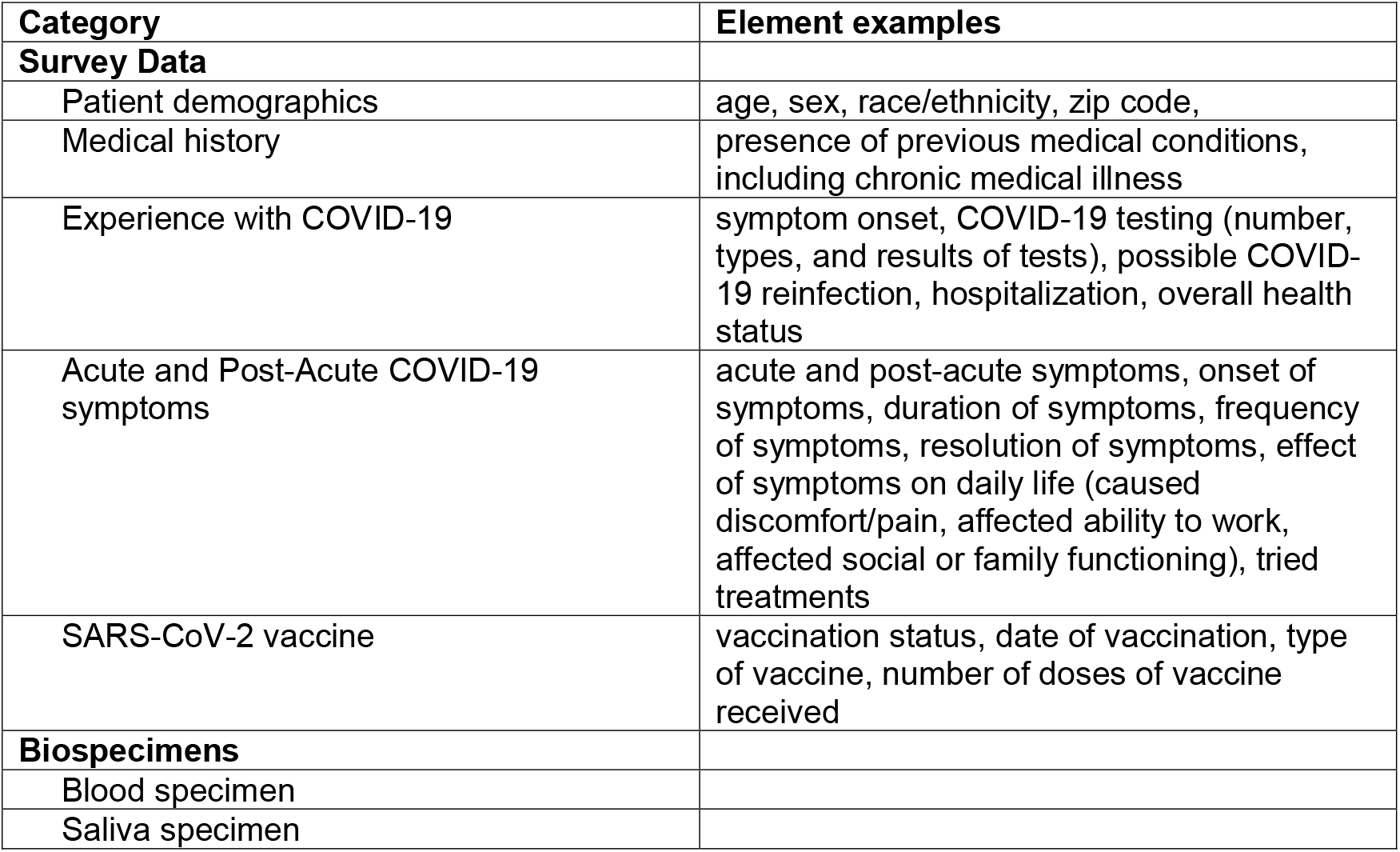
Data elements

| **Category** | **Element examples** |
| --- | --- |
| **Survey Data** |  |
| Patient demographics | age, sex, race/ethnicity, zip code, |
| Medical history | presence of previous medical conditions, including chronic medical illness |
| Experience with COVID-19 | symptom onset, COVID-19 testing (number, types, and results of tests), possible COVID-19 reinfection, hospitalization, overall health status |
| Acute and Post-Acute COVID-19 symptoms | acute and post-acute symptoms, onset of symptoms, duration of symptoms, frequency of symptoms, resolution of symptoms, effect of symptoms on daily life (caused discomfort/pain, affected ability to work, affected social or family functioning), tried treatments |
| SARS-CoV-2 vaccine | vaccination status, date of vaccination, type of vaccine, number of doses of vaccine received |
| **Biospecimens** |  |
| Blood specimen |  |
| Saliva specimen |  |

## Statistical Analyses

### Self-report Survey Data Analyses

The primary outcome for the study is whether people’s overall health condition improves, does not change, or worsens after receiving the COVID-19 vaccine. We will measure this outcome with the question: “Would you say that your overall health, as compared to your health before the vaccine, is worse, better, or the same?” We will ask this question at 2, 6, and 12 weeks after the first dose of the vaccine. As our primary outcome, therefore, we will report the number of people who felt better, worse, or the same by 2 week, 6 week, and 12 week intervals. We will also report the number of people who felt better, worse, or the same at the end of the study (according to the 12 week survey). For each group of people by overall health condition change, we will also report any immune response trends that were found through saliva-based or blood-based analyses (below). In order to report this primary outcome, we will also report the average number of symptoms experienced by people at each time point (pre-vaccine, 2-weeks-post-vaccine, 6-weeks-post-vaccine, and 12-weeks-post-vaccine).

Our secondary outcome for the study is whether specific symptoms or clusters of symptoms are more likely than others to either improve or worsen after vaccination. Symptom changes will be described by whether participants report that symptoms have improved, changed, or stayed the same since before the vaccine. Symptom changes will also be described by changes in their severity and in their frequency and duration. The groups of symptoms will be data-driven; symptoms will not be grouped based on pre-specified categories. As for the primary outcome, we will report the overall outcome, whether symptoms were likely to improve or worsen overall after the entire study, and the longitudinal outcome, how symptoms changed over the 12-week period after vaccination.

Standard descriptive summaries will summarize baseline and demographic characteristics (e.g., means and standard deviations for continuous variables such as age and percentages for categorical variables such as gender, medians, and quartiles for skewed data). Missing values will be handled by exclusion or multiple imputation. We will also examine the effect of age, sex, and race on the type, presence, severity, and timing of various symptom complexes and their relationship with outcomes. Hospitalizations will also be reported longitudinally, and the association between participant symptom complexes and hospitalizations will be reported.

### Saliva-based Analyses

Through longitudinal studies of groups of mild, moderate, and severe COVID-19 patients, our group has found the mucosal viral load (as measured through saliva studies) is a powerful indicator of clinical trajectory in COVID-19. We hypothesize that the mucosal host response may also reveal unique insights into the immune response that may not be evident in our other serologic analyses. We will measure antibody responses and perform cytokine studies using ELISA from saliva samples at all planned time points. Additionally, we will utilize a 71-plex cytokine array (Eve technologies) as previously described by our group that is commercially available.

### Blood-based Analyses

Immunological profiling, immunophenotyping, antibody and T cell assays have been described in **Table 3**.

**Table 3.**
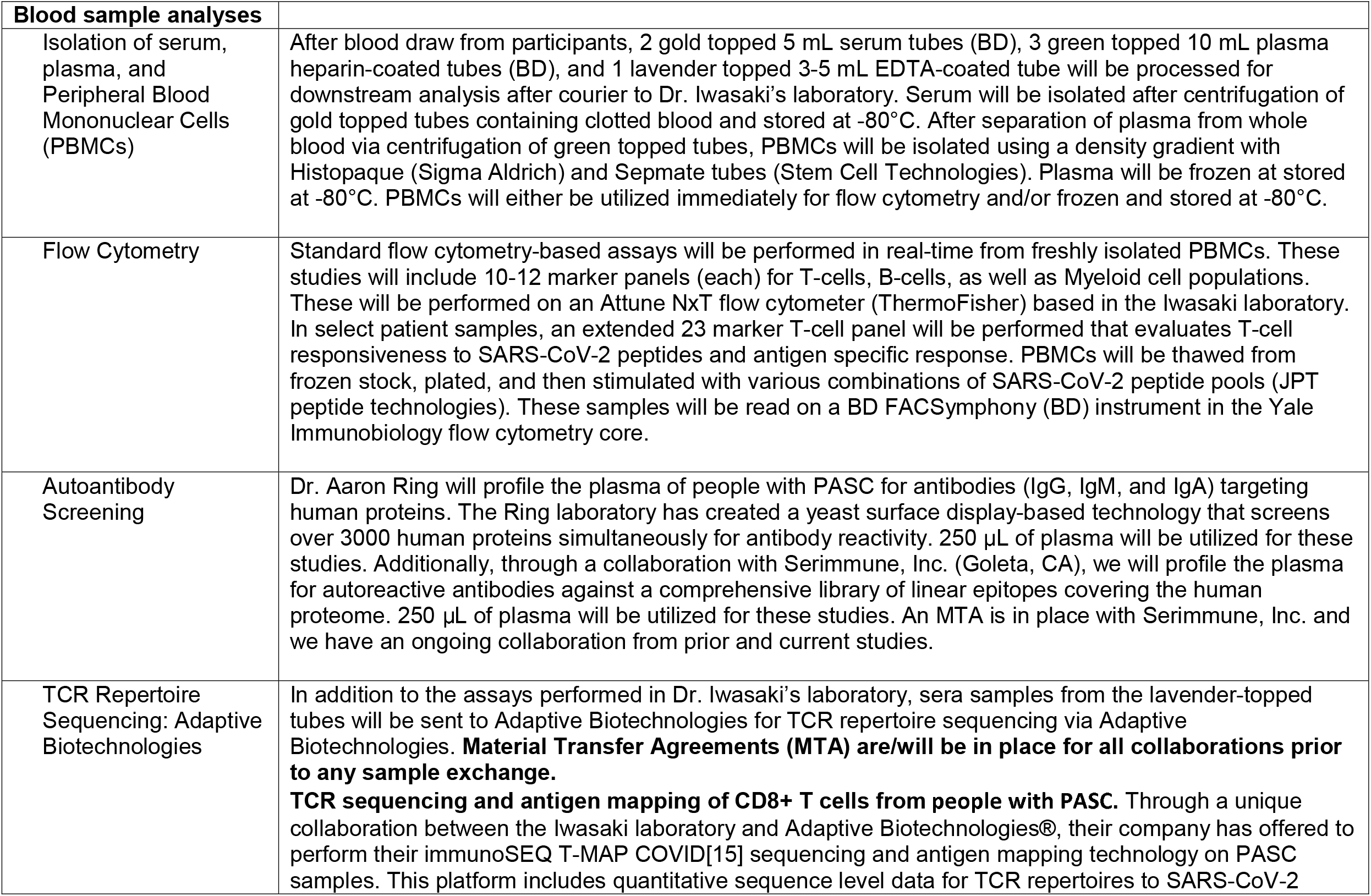
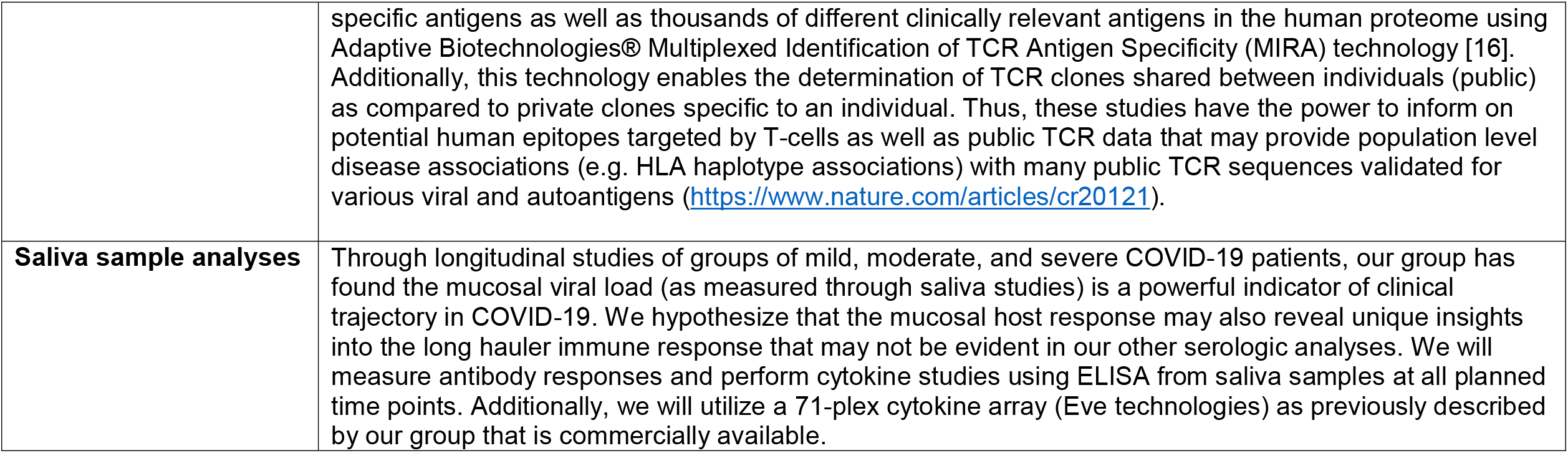
Details of immune tests to be conducted on blood and saliva specimens (immunological profiling, immunophenotyping, and antibody and T cell assays.)

⍰ *Isolation of serum, plasma, and Peripheral Blood Mononuclear Cells (PBMCs)* After blood draw from participants, 2 gold topped 5 mL serum tubes (BD), 3 green topped 10 mL plasma heparin-coated tubes (BD), and 1 lavender topped 3-5 mL EDTA-coated tube will be processed for downstream analysis after courier to Dr. Iwasaki’s laboratory. Serum will be isolated after centrifugation of gold topped tubes containing clotted blood and stored at −80°C. After separation of plasma from whole blood via centrifugation of green topped tubes, PBMCs will be isolated using a density gradient with Histopaque (Sigma Aldrich) and Sepmate tubes (Stem Cell Technologies). Plasma will be frozen at stored at −80°C. PBMCs will either be utilized immediately for flow cytometry and/or frozen and stored at −80°C.
⍰ *Flow Cytometry* Standard flow cytometry-based assays will be performed in real-time from freshly isolated PBMCs. These studies will include 10-12 marker panels (each) for T-cells, B-cells, as well as Myeloid cell populations. These will be performed on an Attune NxT flow cytometer (ThermoFisher) based in the Iwasaki laboratory. In select patient samples, an extended 23 marker T-cell panel will be performed that evaluates T-cell responsiveness to SARS-CoV-2 peptides and antigen specific response. PBMCs will be thawed from frozen stock, plated, and then stimulated with various combinations of SARS-CoV-2 peptide pools (JPT peptide technologies). These samples will be read on a BD FACSymphony (BD) instrument in the Yale Immunobiology flow cytometry core.
⍰ *Autoantibody screening* Dr. Ring will profile the plasma of people with PASC for antibodies (IgG, IgM, and IgA) targeting human proteins. The Ring laboratory has created a yeast surface display-based technology that screens over 3000 human proteins simultaneously for antibody reactivity. 250 μL of plasma will be utilized for these studies. Additionally, through a collaboration with Serimmune, Inc. (Goleta, CA), we will profile the plasma for autoreactive antibodies against a comprehensive library of linear epitopes covering the human proteome. 250 μL of plasma will be utilized for these studies. An MTA is in place with Serimmune, Inc. and we have an ongoing collaboration from prior and current studies.
⍰ *TCR Repertoire Sequencing: Adaptive Biotechnologies* In addition to the assays performed in Dr. Iwasaki’s laboratory, sera samples from the lavender-topped tubes will be sent to Adaptive Biotechnologies for TCR repertoire sequencing via Adaptive Biotechnologies. Material Transfer Agreements (MTA) are/will be in place for all collaborations prior to any sample exchange. *TCR sequencing and antigen mapping of CD8+ T cells from people with PASC*. Through a unique collaboration between the Iwasaki laboratory and Adaptive Biotechnologies®, their company has offered to perform their immunoSEQ T-MAP COVID[15] sequencing and antigen mapping technology on PASC samples. This platform includes quantitative sequence level data for TCR repertoires to SARS-CoV-2 specific antigens as well as thousands of different clinically relevant antigens in the human proteome using Adaptive Biotechnologies® Multiplexed Identification of TCR Antigen Specificity (MIRA) technology [16]. Additionally, this technology enables the determination of TCR clones shared between individuals (public) as compared to private clones specific to an individual. Thus, these studies have the power to inform on potential human epitopes targeted by T-cells as well as public TCR data that may provide population level disease associations (e.g. HLA haplotype associations) with many public TCR sequences validated for various viral and autoantigens (https://www.nature.com/articles/cr20121). We will compare the deep immune profiling among people who improved, did not change and worsened, based on their overall impression of their symptoms.

## SUMMARY

We are conducting the Yale COVID Recovery Study to understand the trends in how the COVID-19 vaccine affects people with PASC, and the mechanisms behind these changes. By collecting information from participants before and after the vaccine, using self-report surveys and immune analyses with blood and saliva specimens, we hope to provide a holistic account of how the vaccine changes both the health trajectory and immune system of participants. We are partnering with patients and patient groups and the study design and surveys reflect their contributions and input. These groups have also contributed to ensure that the study answers patient questions about the vaccine and its effects, especially given the lack of clinical information for people with PASC. While study results will be disseminated, our purpose in sharing the surveys and study design now is to share information that may be used for other studies of PASC, especially given the lack of consensus and knowledge on the condition and the importance of patient input in better understanding how to measure changes in the experience of PASC.

## Supporting information

Appendix 1: Consent Form

Appendices 2-6: Questionnaires

## Data Availability

This protocol and surveys are available to any researcher who requests them and can be found, in addition to MedRxiv, by emailing.

---

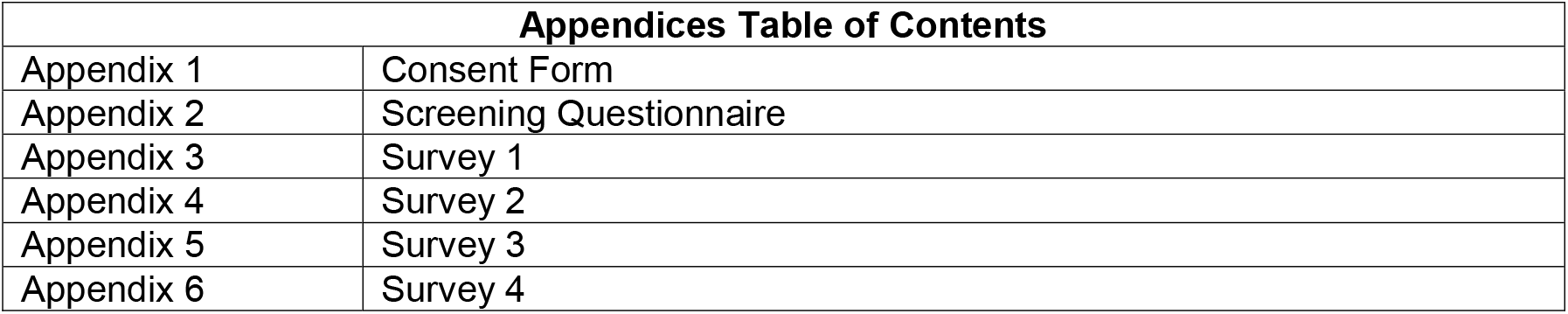

| **Appendices Table of Contents** |  |
| --- | --- |
| Appendix 1 | Consent Form |
| Appendix 2 | Screening Questionnaire |
| Appendix 3 | Survey 1 |
| Appendix 4 | Survey 2 |
| Appendix 5 | Survey 3 |
| Appendix 6 | Survey 4 |

