## Appendix 1: Consent Form for "Change in Symptoms and Immune Response in People with Post-Acute Sequelae of SARS-Cov-2 Infection (*PASC*) After SARS-Cov-2 Vaccination"

### Consent to Participate in the Yale COVID Recovery Study

Please read the following consent form carefully and add your name and signature at the bottom if you agree to participate. If you have any questions, please. Thank you for your interest!

Key Information:

We invite you to participate in a research study in association with the Yale University School of Medicine. The purpose of this study is to measure changes in immune response and Long Covid symptoms before and after getting the COVID-19 vaccine by collecting blood and saliva samples, as well as having participants fill out 4 symptom surveys. In this study we promise to tell you about all that we learn. In this study, we will ask you to give blood and saliva samples 3 times at either of 2 Yale blood draw sites located in New Haven County. You will be able to provide the blood and saliva samples on either a Monday or Wednesday morning (between 8 am and 11 am) and can pick which day is most convenient for you. In addition to providing samples, we will ask you to complete 4 surveys, which you can do at home on a computer, smartphone, or tablet. If you have been to a Yale New Haven Health system site for medical treatment before, then we will also use the information in your clinical record to understand your medical history.

We are conducting the study using REDCap at Yale, which is a secure online tool designed for research that we will use to enroll participants and send surveys without needing you to come into the office or to travel to any site.

Taking part in this research study is your choice. You do not have to join this study and can choose to leave it at any time for any reason. If you decide not to join this study or choose to leave the study at a later time, your health care, benefits, or relationship with your doctors will not change or be affected, and you will still have access to medical care and your medical records as you would normally do.

The study will continue for as long as the researchers and participants are continuing to learn from the data. We anticipate that the study will continue for no more than 1 year. The study will involve giving blood and saliva samples 3 times and answering 4 surveys. All surveys will take less than 20 minutes to complete. You can use a smart phone, tablet or computer to answer the questions and will receive a link to these surveys by email. You can decide not to answer questions at any time, though we may send a reminder to be sure you have not forgotten. Your information from the study will not be shared with your medical team. If you have a medical issue, you should contact your doctor. If you have an emergency, you should call 911. The study will not be monitored for medical problems or care.

For a detailed description of study procedures, please see the "What are the activities you will be doing if you participate in this study?" section of this consent form.

You may not directly benefit from taking part in this study. Still, we hope that this study will produce knowledge that may help you and others. If you are a Connecticut resident who does not already have a vaccination appointment and would like to be vaccinated, we will share information with you regarding how to make a vaccine appointment at a Yale site.

Detailed Information: Please review the rest of this document for details about the above topics and additional information you should know before deciding whether or not you will participate in this study.

Why are you being invited to participate in this study?

We invite you to be part of this study because you had a SARS-CoV-2 infection, suffer from long covid symptoms, and are planning to receive the SARS-CoV-2 vaccine but have not yet received any doses. Also, to be in this study, you must be 18 years of age or older, English-or-Spanish speaking, and able and willing to consent. You also need access to a personal mobile smartphone (i.e., iPhone or Android phone), tablet (i.e., iPad or Android device or similar), or computer with Internet access, and the ability to go to a Yale blood draw site in New Haven County 3 times.

How many participants will take part in this study?

We will enroll up to 1,000 participants but envision the ability to enroll 100.

Who is paying for the study?

The study is currently being supported by the researchers. We are seeking funding but believe this is too important to wait.

What activities will you be doing if you participate in this study?

People who participate in this study will give blood and saliva samples 3 times, once before you get the vaccine, 6 weeks after getting the vaccine, and then 12 weeks after getting the vaccine. There are 2 Yale labs that you can visit to get your blood drawn. You choose which lab and day is most convenient for you, and we'll send you reminders when it's time to give another sample. You will be able to provide the samples on either a Monday or Wednesday morning (between 8 am and 11 am). A trained phlebotomist will draw 45 mLs of blood from you each time, and a total of 135 mL of blood will be collected from you across the 3 study visits.

We will also ask you to fill out 4 surveys using REDCap at Yale, a secure platform for surveys. We use REDCap at Yale to do our research because the platform does not share your data, and when we download your data to do research, it will not have your name or any identifying information on it.

If you agree to participate in this study, the initial set-up process should take no longer than 20 minutes. You will need to do the following steps:

Enroll in the Study. By agreeing to this consent form, you will be granting your e-consent to this study. We will email you a copy of your consent form, with your name only. You will be directed to the first survey, which should be completed before you receive any dose of the vaccine. We may send you survey reminders by email. In your email inbox, you will have an email from the research team with information as to where and how to provide blood and saliva samples at a Yale site. We may also call you within 2-3 days to share this information with you and answer any questions that you may have. At any point until you receive a dose of the vaccine, you may go to any of the Yale blood draw sites to provide the first blood and saliva samples. Please note: Researchers will not be watching or evaluating your symptoms as part of this study, including your responses to the questionnaires. The researchers will not share the information with your medical team. If you begin to experience new symptoms or any medical issues, please contact your doctor. In case of a life-threatening emergency, call 911 immediately.

Continuous study process: You will give blood and saliva a total of 3 times, once before vaccination and twice after vaccination. You will answer surveys a total of 4 times, once before vaccination and three times after vaccination. You will receive an email when it is time to complete the next survey or to provide samples again. If you do not yet know when your vaccine appointment will be, we will be in touch via email or calling to see when you are getting your vaccine in order to know when it is time to schedule your next sample collection or survey. You may always ask us any questions by emailing.

Will your information be used for research in the future?

Information collected from you for this study may be used for future research or shared with other researchers. If this happens, we will remove any information that could identify you before it is shared. Since identifying information will be removed, we will not ask you for additional consent. We are asking now for your permission to do that. The goal is to have as many good scientists working on these data as possible to accelerate progress. The shared data will always be de-identified to protect your identity.

Will you be contacted about participating in future research?

We may contact you about participating in future research, but the decision to participate will always be up to you.

What are the risks and discomforts of participating in this study?

While giving blood may be uncomfortable, the risks are minimal and may include pain, a bruise at the point where the blood is taken, redness and swelling of the vein and in rare case infection, and fainting.

All research studies involve some risk to participant privacy. As with every study, you share your personal health information with researchers, and there is a risk that someone might access that information improperly. However, the research team has strict protocols in place to control access to your data, including keeping audit logs of who has accessed your data on REDCap at Yale. You will be able to withdraw from the study and stop sharing data through REDCap at Yale at any time for any reason. The researchers will not give your doctors any information from the questionnaires you fill out. The data will be kept confidential and will not be part of your medical record. In addition, access to your medical record is through the Yale New Haven Health EPIC system, which is a secure system that only the study coordinators and researchers will have access to for the study. Any information extracted from your medical record will also be kept on Yale secure servers. All email communication between you and our study team will be secure. All emails to participants will be sent from a Yale email address and will be encrypted.

While participating in this study, we will ask you to fill out multiple surveys during the study period, which will take some of your time. Some of the surveys may include sensitive questions that you feel uncomfortable answering. We encourage you to answer all the questions as we can learn better that way, but the decision is up to you.

What if there is new information that may affect your decision to participate in this study?

We will tell you about important findings (either good or bad). The study risks are low, and we do not expect that to change.

Will you receive your results from the study?

We will share the results of the T-Detect test, which is a test from Adaptive Biotechnologies to determine whether there are T cells present in your body. We will share with you the results from each of the 3 study visits, but we will share all results after your last study visit. We encourage you to please visit your doctor for more information regarding the study results. The other laboratory tests used in this study are research tests and are not approved for clinical use. Officially, the laboratory for the other tests is not approved under the Clinical Laboratory Improvement Amendments (CLIA), which regulates laboratory testing performed on humans. The results of a research test conducted in a non-CLIA-certified laboratory cannot be shared with study participants. As such, we cannot share your individual test results with you for all tests, but we will share the results of the study and our findings with everyone who participates as they become available. You will also receive notification of scientific presentations and paper publications from the study. You will also be able to download a record of all of your survey responses.

Can you leave or be removed from this study?

You have the right to leave a study at any time without penalty. If you do become a study participant, you are free to quit and withdraw from this study at any time during its course. You can withdraw from the study at any point by emailing. When you withdraw from this study, we will not gather any new information after that date. Information we already collected will be retained and used as noted above. Withdrawing from the study will involve no penalty or loss of benefits to which you are otherwise entitled. It will not harm your relationship with your doctors.

The researchers have the right to stop your participation in this study without your consent if:

They believe it is in your best interests You do not follow the instructions They cancel the study for any reason

What about the confidentiality of your medical information?

This authorization is voluntary. The Yale University School of Medicine and its affiliate ("Yale New Haven Health") will not withhold or refuse your treatment, payment, enrollment, or eligibility for benefits if you do not sign this authorization. You do not have to sign this authorization, but not signing this authorization will exclude you from the study.

The study team will collect PHI about you for this research during the study. PHI is your health information, which includes your medical history and new information obtained from this study. This new information includes answers you provide to survey questions and results from laboratory tests using the blood samples you provide.

The study team may also use your medical record information from the Yale New Haven Health system's electronic health record (also known as the EPIC system) to see whether you have received any dose of the vaccine. (If a person is vaccinated in Connecticut and has received medical care from any Yale New Haven Health provider, then the vaccination appointment will be noted in the Yale New Haven Health electronic health record.) We will only use this information to know when to send reminders to schedule a sample collection.

The study team may also use your medical record information from the Yale New Haven Health system's electronic health record (also known as the EPIC system) to see your medical history and to better understand your experience with COVID-19 and any relevant prior medical conditions. We plan to collect data including date of birth, medical record number, vital signs, provider encounters, family history, laboratory findings, procedures, problem list, ICD diagnoses, medicines administered and prescribed. We will also be comparing this information with biological findings to determine whether anything predicts how the vaccine changes Long Covid symptoms.

Records identifying you (such as containing your name or phone number) will be deleted at the conclusion of the study, at which point we will keep on secure Yale servers only deidentified information for up to 10 years.

The information we are asking to use and share is called "Protected Health Information." It is protected by a federal law called the Privacy Rule of the Health Insurance Portability and Accountability Act (HIPAA). In general, we cannot use or share your health information for research without your permission. If you want, we can give you more information about the Privacy Rule. Also, if you have any questions about the Privacy Rule and your rights, you can speak to Yale Privacy Officer at 203-432-5919.

How will you use and share my information?

We will use your information to conduct the study described in this consent form. In the spirit of open science, we may also share de-identified data with other scientists to support further investigations.

You have a right to inspect and copy the information to be disclosed with this authorization. You may obtain a copy of the data by contacting Dr. Harlan Krumholz at.

Suppose you no longer want to be in the study and do not want your future health information to be used. In that case, you may change your mind and revoke (take back) this authorization at any time by contacting Dr. Harlan Krumholz at. If you revoke your authorization, you will no longer be allowed to participate in the study. Previously authorized individuals/entities may still use or disclose health information that they have already obtained about you as necessary to maintain the integrity or reliability of the current study.

This authorization is valid for the entirety of this research study. It will expire upon completion of the study or if you revoke (take back) it. If you withdraw from this study, the data already collected from you may remain in the study records.

Dr. Harlan Krumholz, the Principal Investigatory of the study, and the co-investigators and study coordinators may see your health information in connection with this study. We may also share your information with trained phlebotomists who will provide services to you in connection with this study and laboratories that will analyze your deidentified blood and saliva samples. We will do our best to make sure your information stays private. But, if we share information with people who do not have to follow the Privacy Rule, your information will no longer be protected by the Privacy Rule. Let us know if you have questions about this. However, to better protect your health information, agreements are in place with these individuals and/or companies that require that they keep your information confidential.

Records of participation in this study will be maintained and kept confidential as required by law. Representatives from Yale University, the Yale Human Research Protection Program and the Institutional Review Board (the committee that reviews, approves, and monitors research on human participants), who are responsible for ensuring research compliance. These individuals are required to keep all information confidential.

What are the benefits to participate in this study?

The study might benefit you by giving you information about your body that you can share with your doctor. We will tell you after each visit whether we found T-cells, which are a special kind of fighter cell in your body that try to fight an invader, like COVID-19. Sometimes these cells can hurt the body if they fight for a long time or if there are too many of them. When we share your results with you, we will give you a sheet with information about what it means.

What are the costs to participate in this study?

There are no costs to you for participating in this research.

Will you be paid for your participation in this study?

There is currently no payment for the study.

What if you are injured due to your participation in this study?

We do not expect you to be potentially injured by participating.

The Yale University School of Medicine has no program for financial compensation or other forms of payment for injuries, which you may incur due to participation in this study. By signing this form, you are not giving up any legal rights to seek injury compensation.

Who can you contact for more information about this study?

Questions are encouraged. If you have further questions about this study, you may email Dr. Harlan Krumholz at. We will make every attempt to respond to all questions in a timely manner, however, please note that this email address is not monitored all the time and that you should contact your healthcare provided for any questions regarding your health.

Whom can you contact if you have concerns about your rights as a study participant?

You may address questions about the rights of research participants to the Yale Human Research Protection Program at 1-203-785-4688.

What are your rights as a study participant?

Taking part in this study is voluntary. If you choose not to participate in this study or to leave the study at any time for any reason, your health care, benefits, or relationship with your doctors will not change or be affected, and you will still have access to medical care and your medical records as you would normally do.

---

Authorization and Permission

I have read this form and have decided to participate in the Yale COVID Recovery Study as described above. Its general purposes, teh particulars of my involvement and possible hazards and inconveniences have been explained to my satisfaction. I understand that I will be provided with a copy of this consent form.

By signing this form, I give permission to the researchers to use and give out information about me for the purposes described in this form. By refusing to give permission, I understand that I will not be able to participate in this research.

---

1) Today's Date

______________________________

2) Participant First Name

______________________________

3) Participant Last Name

______________________________

4) Please add your signature

____________________________________
