## Appendices 2-6: Questionnaires for "Change in Symptoms and Immune Response in People with Post-Acute Sequelae of SARS-Cov-2 Infection (*PASC*) After SARS-Cov-2 Vaccination"

### Screening Questionnaire

Please complete the survey below to determine eligibility for the Yale COVID Recovery Study.

If you have any questions about the survey, please.

Thank you for your interest!

| | |
|---|---|
| Please enter your first name. | __________________ |
| Please enter your last name. | __________________ |
| Please provide an email address that we may use to contact you. | __________________ |
| What is your birthdate? | __________________ |
| What is a phone number that we may use to reach you? | __________________ |
| Are you currently 18 years or older? | ◯ Yes<br>◯ No |
| Have you ever tested positive for COVID-19? (The type of test may have been PCR, antigen, antibody, or T-test.) | ◯ Yes<br>◯ No |
| Have you ever been hospitalized due to COVID-19? | ◯ Yes<br>◯ No |
| Have you ever been diagnosed with COVID-19 by a clinician? | ◯ Yes<br>◯ No |
| In your opinion, do you have symptoms more than 2 months after your initial COVID-19 infection (long covid) that interfere with your quality of life or with your ability to do your normal activities? | ◯ Yes<br>◯ No |
| Have you received any dose(s) of a COVID-19 vaccine? | ◯ Yes<br>◯ No |
| Do you plan to get the COVID-19 vaccine? (Note: You do not need to currently have a vaccine appointment to be planning to get the vaccine.) | ◯ Yes<br>◯ No |
| Are you willing to give a blood and saliva sample 3 times at one of the following locations on a Monday or Wednesday morning (between 8 am and 11 am):<br><br>North Haven Medical Center<br><br>Yale New Haven Hospital, Smilow Center | ◯ Yes<br>◯ No |
| Has consent been completed and verified? | ☐ Yes<br>☐ No |

### Survey 1

Thank you for taking Survey 1 for the Yale COVID Recovery Study!

Please remember that you may stop this survey and come back to it at any point.

You may also find it helpful to have the below items ready as you complete the survey. If you do not have these items, please still fill in the information as best you can.

Testing results: dates, type (PCR, antigen) and result of tests. If you do not remember the exact date, the estimated date is enough.Symptom time and severity: your symptom log. You will be asked to pick symptoms you have had since you began having COVID-19 symptoms.

**There are 3 sections in this survey. In this section, we will ask for your contact information so that we may follow up with future research opportunities. We will also ask demographic questions so that we can understand how Long COVID is affecting the population.**

What is your birthdate? ______________________

What is your zip code? ______________________

Do you plan on getting the vaccine?
- ◯ Yes, and I have an appointment scheduled
- ◯ Yes, but I don't have an appointment yet
- ◯ I'm not planning on getting the vaccine
- ◯ I have already gotten at least one dose of the COVID-19 vaccine

When is your vaccine appointment? ______________________

What is your gender?
- ◯ Male
- ◯ Female
- ◯ Not listed ______

Please specify (optional): ______________________

What race(s) or ethnicity do you consider yourself to be?
*Please choose 1 or more
- ☐ American Indian/Alaska Native
- ☐ Asian ______
- ☐ Black or African American
- ☐ Hispanic or Latino ______
- ☐ Native Hawaiian/Other Pacific Islander ______
- ☐ White
- ☐ Some other race ______

Please specify your race and/or ethnicity ______________________

Please specify (select as many as apply to you)
- ☐ Chinese
- ☐ Filipino
- ☐ Asian Indian
- ☐ Vietnamese
- ☐ Korean
- ☐ Japanese
- ☐ Other Asian (e.g., Pakistani, Cambodian, Hmong)

Please specify (select as many as apply to you):

- ☐ Native Hawaiian
- ☐ Samoan
- ☐ Chamorro
- ☐ Other Pacific Islander (e.g., Tongan, Fijian, Mashallese)

---

Please specify (select as many as apply to you)

- ☐ Mexican/Mexican American
- ☐ Puerto Rican
- ☐ Dominican (Republic)
- ☐ Cuban/Cuban American
- ☐ Central or South American
- ☐ Other Latino/Hispanic

---

Have you ever been told by a doctor that you have any of the following?

- ☐ Any allergies
- ☐ Arthritis (including rheumatoid arthritis, gout, lupus, or fibromyalgia)
- ☐ Asthma
- ☐ Blood disorder (including sickle cell disease or Thalassemia)
- ☐ Cancer or malignancy of any kind
- ☐ Cerebrovascular conditions affecting blood vessels to or in the brain (including stroke)
- ☐ Chronic lung disease (including emphysema, chronic bronchitis, chronic obstructive pulmonary disease (COPD), or pulmonary fibrosis)
- ☐ Cystic fibrosis
- ☐ Diabetes
- ☐ Gastrointestinal issues (including IBS or acid reflux)
- ☐ Heart attack, also called myocardial infarction
- ☐ Heart conditions (including coronary artery disease or cardiomyopathies)
- ☐ Heart failure
- ☐ High cholesterol
- ☐ History of organ transplant (including kidney, liver, heart, or lung)
- ☐ Hypertension or high blood pressure
- ☐ Immunocompromised state (including weakened immun system from blood or bone marrow transplant, immune deficiencies, HIV, use of corticosteroids, or use of other immune-weakening medicines)
- ☐ Kidney disease
- ☐ Liver disease
- ☐ Mental health conditions (including anxiety, depression, bipolar disorder, schizophrenia, obsessive or compulsive disorder)
- ☐ Neurologic conditions (including seizures or dementia)
- ☐ Spinal disorder(s)
- ☐ Other

---

Please describe "Other":

______________________________

**Now we are going to ask you questions about your experience with COVID-19. Please answer as best you can, and we understand that dates may not be exact. You have 2 survey sections to go.**

When did your symptoms due to COVID-19 begin? __________________________________

Have you been tested for COVID-19 at least once?
- ◯ Yes
- ◯ No

What was the date of the first time you were tested for COVID-19? __________________________________

What was the type of test?
- ◯ PCR test
- ◯ Antigen test
- ◯ Not sure

What was the result?
- ◯ Positive
- ◯ Negative
- ◯ Inconclusive
- ◯ Awaiting result

Were you tested for COVID more than once?
- ◯ Yes
- ◯ No

How many times were you tested for COVID-19? __________________________________

What was the date of your second COVID-19 test? __________________________________

What was the result of your second test?
- ◯ Positive
- ◯ Negative
- ◯ Inconclusive
- ◯ Awaiting result

What was the date of your most recent COVID-19 test? __________________________________

What was the result of your most recent COVID-19 test?
- ◯ Positive
- ◯ Negative
- ◯ Inconclusive
- ◯ Awaiting result

If you ever tested positive for COVID-19, when was your most recent positive test? __________________________________

Do you believe that you were reinfected with COVID-19?
- ◯ Yes
- ◯ No

Were you ever hospitalized due to COVID-19?
- ◯ Yes
- ◯ No

Have you visited the hospital or been hospitalized for COVID-19 symptoms that began more than two weeks after you first got sick?
- ◯ Yes
- ◯ No

Which of the following symptoms have you experienced due to COVID-19? Select as many as apply. As a reminder, you stated that your symptoms began on [s1_symptom_date]. *select all that apply

Symptoms

Have you experienced this symptom (at any time)?

Approximately when did you begin experiencing this symptom?

Does/did this symptom come and go?

Has this symptom gone away completely?

Abdominal pain

______

______

______

______

Abnormally low temperature

______

______

______

______

Acid reflux

______

______

______

______

Afternoon or evening fevers/low-grade fevers

______

______

______

______

Anemia (low number of red blood cells)

______

______

______

Anxiety

______

______

______

______

Arrythmia (improper beating of the heart due to electrical impulse problems)

______

______

______

______

Bilateral neck throbbing around lymph nodes

______

______

______

______

Blurry vision

______

______

______

______

Bone aches in extremities

______

______

______

______

Brain fog

______

______

______

______

Brain pressure

______

______

______

______

Bulging veins

______

______

______

______

Burning sensations

______

______

______

______

Bruising of skin

______

______

______

______

Calf cramps

______

______

______

______

Change in nails (i.e. white spots, brittleness, change in moons)

______

______

______

______

Changes in voice

______

______

______

______

Changed sense of taste

______

______

______

______

Chills but no fever

______

______

______

______

Clogged ears

______

______

______

______

Cold burning feeling in lungs

______

______

______

______

Confusion

______

______

______

______

Congested or runny nose

______

______

______

______

Constant thirst

______

______

______

______

Costochondritis (inflammation of the cartilage that connects a rib to the breastbone)

______

______

______

______

Cough

______

______

______

______

Coughing up blood

______

______

______

______

Covid toes (tender or itchy rash or chilblains on the toes or foot)

______

______

______

______

Cracked or dry lips

______

______

______

______

Dental problems (i.e. chipped tooth, tooth loss)

______

______

______

______

Diarrhea

______

______

______

______

Difficulty concentrating or focusing

______

______

______

______

Difficulty sleeping

______

______

______

______

Difficulty speaking properly

______

______

______

______

Discoloration of the skin (for example: purple or blue on the hands or feet, no blistering)

______

______

______

______

Dizziness

______

______

______

______

Dry eyes

______

______

______

______

Dry or peeling skin

______

______

______

______

Dry scalp or dandruff

______

______

______

______

Dry throat

______

______

______

______

Ear pain/earache

______

______

______

______

Elevated thyroid hormones

______

______

______

______

Extreme pressure at base of head or occipital nerve

______

______

______

______

Eye stye or infection

______

______

______

______

Fatigue

______

______

______

______

Feeling irritable

______

______

______

______

Feeling of burning skin

______

______

______

______

Fever or chills

______

______

______

______

Floaters or flashes of light in vision

______

______

______

______

Foot pain

______

______

______

______

GERD (acid reflux) with excessive salivation

______

______

______

______

Goiter or lump in throat

______

______

______

______

Hair loss

______

______

______

______

Hand or wrist pain

______

______

______

______

Headache

______

______

______

______

Heart palpitations (heart skipping a beat or racing)

______

______

______

______

Heat intolerance

______

______

______

______

High blood pressure

______

______

______

______

Hormone imbalances

______

______

______

______

“Hot” blood rush

______

______

______

______

Inability to cry

______

______

______

______

Inability to exercise or be active

______

______

______

______

Inability to yawn

______

______

______

______

Internal tremors or buzzing/vibration

______

______

______

______

Irregular or skipped menstrual cycles

______

______

______

______

Jaw pain

______

______

______

______

Joint pain

______

______

______

______

Kidney issues or protein in urine

______

______

______

______

Kidney pain

______

______

______

______

Low blood oxygen

______

______

______

______

Low blood pressure

______

______

______

______

Lower back pain

______

______

______

______

Loss of appetite

______

______

______

______

Loss of hearing

______

______

______

______

Loss or decrease in quality of vision

______

______

______

______

Lump in throat/difficulty swallowing

______

______

______

______

Memory problems

______

______

______

______

Menstrual cycles that are heavier or lighter than normal

______

______

______

______

Mid-back pain at base of ribs

______

______

______

______

Mouth sores or sore tongue

______

______

______

______

Muscle or body aches

______

______

______

______

Muscle twitching

______

______

______

______

Nausea or vomiting

______

______

______

______

Neck muscle pain

______

______

______

______

Nerve pain

______

______

______

______

Nerve sensations (tingling, pins and needles, numbness)

______

______

______

______

Neuropathy in feet and hands (weakness, numbness, and pain)

______

______

______

______

New allergies

______

______

______

______

Night sweats

______

______

______

______

Nightmares

______

______

______

______

Painful scalp

______

______

______

______

Partial or complete loss of sense of smell

______

______

______

______

Partial or complete loss of sense of taste

______

______

______

______

Persistent chest pain or pressure

______

______

______

______

Personality change (drastic)

______

______

______

______

Petechiae (pinpoint rash)

______

______

______

______

Phantom smells

______

______

______

______

Phlegm in back of throat

______

______

______

______

Post-exertional malaise (worsened symptoms or flu-like symptoms after exertion)

______

______

______

______

Postnasal drip

______

______

______

______

Rash

______

______

______

______

Reflux or heartburn

______

______

______

______

Runny nose

______

______

______

______

Sadness

______

______

______

______

Seizures

______

______

______

______

Sharp or sudden chest pain

______

______

______

______

Shortness of breath or difficulty breathing

______

______

______

______

Shortness of breath or exhaustion from bending over

______

______

______

______

Sleeping more than normal

______

______

______

______

Sore throat

______

______

______

______

Spinal issues

______

______

______

______

Spikes in blood pressure

______

______

______

______

Swollen hands or feet

______

______

______

______

Swollen lymph nodes

______

______

______

______

Syncope (fainting)

______

______

______

______

Tachycardia (rapid heartbeat) at rest

______

______

______

______

Tachycardia (rapid heartbeat) after standing up

______

______

______

______

Thrush (white fungal infection in the mouth or throat)

______

______

______

______

Tinnitus or humming in ears

______

______

______

______

Tremors or shakiness

______

______

______

______

Upper back pain

______

______

______

______

UTI (urinary tract infection)

______

______

______

______

Weakened neck

______

______

______

______

Weight gain

______

______

______

______

Other symptom(s)

______

| | |
|---|---|
| Please specify "Other symptoms" | ______________________ |
| Have you experienced this symptom (at any time)? | ☐ Yes |
| When did the symptom begin? | ______________________ |
| Does/Did this symptom come and go? | ◯ Yes<br>◯ No |
| Has the symptom gone away completely? | ◯ Yes<br>◯ No |
| Have you experienced this symptom (at any time)? | ☐ Yes |

**While experiencing these symptoms, how much do/did they bother you in terms of discomfort or pain?**

| | Not at all | A little bit | Somewhat | Quite a bit | Very much |
|---|---|---|---|---|---|
| Abdominal pain | ◯ | ◯ | ◯ | ◯ | ◯ |
| Abnormally low temperature | ◯ | ◯ | ◯ | ◯ | ◯ |
| Acid reflux | ◯ | ◯ | ◯ | ◯ | ◯ |
| Afternoon or evening fevers/low-grade fevers | ◯ | ◯ | ◯ | ◯ | ◯ |
| Anemia (low number of red blood cells) | ◯ | ◯ | ◯ | ◯ | ◯ |
| Anxiety | ◯ | ◯ | ◯ | ◯ | ◯ |
| Arrythmia (improper beating of the heart due to electrical impulse problems) | ◯ | ◯ | ◯ | ◯ | ◯ |
| Bilateral neck throbbing around lymph nodes | ◯ | ◯ | ◯ | ◯ | ◯ |
| Blurry vision | ◯ | ◯ | ◯ | ◯ | ◯ |
| Bone aches in extremities | ◯ | ◯ | ◯ | ◯ | ◯ |
| Brain fog | ◯ | ◯ | ◯ | ◯ | ◯ |
| Brain pressure | ◯ | ◯ | ◯ | ◯ | ◯ |
| Bulging veins | ◯ | ◯ | ◯ | ◯ | ◯ |
| Burning sensations | ◯ | ◯ | ◯ | ◯ | ◯ |
| Bruising of skin | ◯ | ◯ | ◯ | ◯ | ◯ |
| Calf cramps | ◯ | ◯ | ◯ | ◯ | ◯ |
| Change in nails (i.e. white spots, brittleness, change in moons) | ◯ | ◯ | ◯ | ◯ | ◯ |
| Changes in voice | ◯ | ◯ | ◯ | ◯ | ◯ |
| Changed sense of taste | ◯ | ◯ | ◯ | ◯ | ◯ |
| Chills but no fever | ◯ | ◯ | ◯ | ◯ | ◯ |
| Clogged ears | ◯ | ◯ | ◯ | ◯ | ◯ |
| Cold burning feeling in lungs | ◯ | ◯ | ◯ | ◯ | ◯ |
| Confusion | ◯ | ◯ | ◯ | ◯ | ◯ |
| Congested or runny nose | ◯ | ◯ | ◯ | ◯ | ◯ |
| Constant thirst | ◯ | ◯ | ◯ | ◯ | ◯ |
| Costochondritis (inflammation of the cartilage that connects a rib to the breastbone) | ◯ | ◯ | ◯ | ◯ | ◯ |
| Cough | ◯ | ◯ | ◯ | ◯ | ◯ |
| Coughing up blood | ◯ | ◯ | ◯ | ◯ | ◯ |

| | | | | | |
|---|---|---|---|---|---|
| Covid toes (tender or itchy rash or chilblains on the toes or foot) | ○ | ○ | ○ | ○ | ○ |
| Cracked or dry lips | ○ | ○ | ○ | ○ | ○ |
| Dental problems (i.e. chipped tooth, tooth loss) | ○ | ○ | ○ | ○ | ○ |
| Diarrhea | ○ | ○ | ○ | ○ | ○ |
| Difficulty concentrating or focusing | ○ | ○ | ○ | ○ | ○ |
| Difficulty sleeping | ○ | ○ | ○ | ○ | ○ |
| Difficulty speaking properly | ○ | ○ | ○ | ○ | ○ |
| Discoloration of the skin (for example: purple or blue on the hands or feet, no blistering) | ○ | ○ | ○ | ○ | ○ |
| Dizziness | ○ | ○ | ○ | ○ | ○ |
| Dry eyes | ○ | ○ | ○ | ○ | ○ |
| Dry or peeling skin | ○ | ○ | ○ | ○ | ○ |
| Dry scalp or dandruff | ○ | ○ | ○ | ○ | ○ |
| Dry throat | ○ | ○ | ○ | ○ | ○ |
| Ear pain/earache | ○ | ○ | ○ | ○ | ○ |
| Elevated thyroid hormones | ○ | ○ | ○ | ○ | ○ |
| Extreme pressure at base of head or occipital nerve | ○ | ○ | ○ | ○ | ○ |
| Eye stye or infection | ○ | ○ | ○ | ○ | ○ |
| Fatigue | ○ | ○ | ○ | ○ | ○ |
| Feeling irritable | ○ | ○ | ○ | ○ | ○ |
| Feeling of burning skin | ○ | ○ | ○ | ○ | ○ |
| Fever or chills | ○ | ○ | ○ | ○ | ○ |
| Floaters or flashes of light in vision | ○ | ○ | ○ | ○ | ○ |
| Foot pain | ○ | ○ | ○ | ○ | ○ |
| GERD (acid reflux) with excessive salivation | ○ | ○ | ○ | ○ | ○ |
| Goiter or lump in throat | ○ | ○ | ○ | ○ | ○ |
| Hair loss | ○ | ○ | ○ | ○ | ○ |
| Hand or wrist pain | ○ | ○ | ○ | ○ | ○ |
| Headache | ○ | ○ | ○ | ○ | ○ |
| Heart palpitations (heart skipping a beat or racing) | ○ | ○ | ○ | ○ | ○ |
| Heat intolerance | ○ | ○ | ○ | ○ | ○ |
| High blood pressure | ○ | ○ | ○ | ○ | ○ |
| Hormone imbalances | ○ | ○ | ○ | ○ | ○ |

| | | | | | |
|---|---|---|---|---|---|
| "Hot" blood rush | ○ | ○ | ○ | ○ | ○ |
| Inability to cry | ○ | ○ | ○ | ○ | ○ |
| Inability to exercise or be active | ○ | ○ | ○ | ○ | ○ |
| Inability to yawn | ○ | ○ | ○ | ○ | ○ |
| Internal tremors or buzzing/vibration | ○ | ○ | ○ | ○ | ○ |
| Irregular or skipped menstrual cycles | ○ | ○ | ○ | ○ | ○ |
| Jaw pain | ○ | ○ | ○ | ○ | ○ |
| Joint pain | ○ | ○ | ○ | ○ | ○ |
| Kidney issues or protein in urine | ○ | ○ | ○ | ○ | ○ |
| Kidney pain | ○ | ○ | ○ | ○ | ○ |
| Low blood oxygen | ○ | ○ | ○ | ○ | ○ |
| Low blood pressure | ○ | ○ | ○ | ○ | ○ |
| Lower back pain | ○ | ○ | ○ | ○ | ○ |
| Loss of appetite | ○ | ○ | ○ | ○ | ○ |
| Loss of hearing | ○ | ○ | ○ | ○ | ○ |
| Loss or decrease in quality of vision | ○ | ○ | ○ | ○ | ○ |
| Lump in throat/difficulty swallowing | ○ | ○ | ○ | ○ | ○ |
| Memory problems | ○ | ○ | ○ | ○ | ○ |
| Menstrual cycles that are heavier or lighter than normal | ○ | ○ | ○ | ○ | ○ |
| Mid-back pain at base of ribs | ○ | ○ | ○ | ○ | ○ |
| Mouth sores or sore tongue | ○ | ○ | ○ | ○ | ○ |
| Muscle or body aches | ○ | ○ | ○ | ○ | ○ |
| Muscle twitching | ○ | ○ | ○ | ○ | ○ |
| Nausea or vomiting | ○ | ○ | ○ | ○ | ○ |
| Neck muscle pain | ○ | ○ | ○ | ○ | ○ |
| Nerve pain | ○ | ○ | ○ | ○ | ○ |
| Nerve sensations (tingling, pins and needles, numbness) | ○ | ○ | ○ | ○ | ○ |
| Neuropathy in feet and hands (weakness, numbness, and pain) | ○ | ○ | ○ | ○ | ○ |
| New allergies | ○ | ○ | ○ | ○ | ○ |
| Night sweats | ○ | ○ | ○ | ○ | ○ |
| Nightmares | ○ | ○ | ○ | ○ | ○ |
| Painful scalp | ○ | ○ | ○ | ○ | ○ |
| Partial or complete loss of sense of smell | ○ | ○ | ○ | ○ | ○ |

| | | | | | |
|---|---|---|---|---|---|
| Partial or complete loss of sense of taste | ○ | ○ | ○ | ○ | ○ |
| Persistent chest pain or pressure | ○ | ○ | ○ | ○ | ○ |
| Personality change (drastic) | ○ | ○ | ○ | ○ | ○ |
| Petechiae (pinpoint rash) | ○ | ○ | ○ | ○ | ○ |
| Phantom smells | ○ | ○ | ○ | ○ | ○ |
| Phlegm in back of throat | ○ | ○ | ○ | ○ | ○ |
| Post-exertional malaise (worsened symptoms or flu-like symptoms after exertion) | ○ | ○ | ○ | ○ | ○ |
| Postnasal drip | ○ | ○ | ○ | ○ | ○ |
| Rash | ○ | ○ | ○ | ○ | ○ |
| Reflux or heartburn | ○ | ○ | ○ | ○ | ○ |
| Runny nose | ○ | ○ | ○ | ○ | ○ |
| Sadness | ○ | ○ | ○ | ○ | ○ |
| Seizures | ○ | ○ | ○ | ○ | ○ |
| Sharp or sudden chest pain | ○ | ○ | ○ | ○ | ○ |
| Shortness of breath or difficulty breathing | ○ | ○ | ○ | ○ | ○ |
| Shortness of breath or exhaustion from bending over | ○ | ○ | ○ | ○ | ○ |
| Sleeping more than normal | ○ | ○ | ○ | ○ | ○ |
| Sore throat | ○ | ○ | ○ | ○ | ○ |
| Spinal issues | ○ | ○ | ○ | ○ | ○ |
| Spikes in blood pressure | ○ | ○ | ○ | ○ | ○ |
| Swollen hands or feet | ○ | ○ | ○ | ○ | ○ |
| Swollen lymph nodes | ○ | ○ | ○ | ○ | ○ |
| Syncope (fainting) | ○ | ○ | ○ | ○ | ○ |
| Tachycardia (rapid heartbeat) at rest | ○ | ○ | ○ | ○ | ○ |
| Tachycardia (rapid heartbeat) after standing up | ○ | ○ | ○ | ○ | ○ |
| Thrush (white fungal infection in the mouth or throat) | ○ | ○ | ○ | ○ | ○ |
| Tinnitus or humming in ears | ○ | ○ | ○ | ○ | ○ |
| Tremors or shakiness | ○ | ○ | ○ | ○ | ○ |
| Upper back pain | ○ | ○ | ○ | ○ | ○ |
| UTI (urinary tract infection) | ○ | ○ | ○ | ○ | ○ |
| Weakened neck | ○ | ○ | ○ | ○ | ○ |
| Weight gain | ○ | ○ | ○ | ○ | ○ |

**After quarantine, how much does/did the symptom impair your ability to work compared to pre-COVID?**

Abdominal pain

Abnormally low temperature

Acid reflux

Afternoon or evening fevers/low-grade fevers

Anemia (low number of red blood cells)

Anxiety

Arrythmia (improper beating of the heart due to electrical impulse problems)

Bilateral neck throbbing around lymph nodes

Blurry vision

Bone aches in extremities

Brain fog

Brain pressure

Bulging veins

Burning sensations

Bruising of skin

Calf cramps

Change in nails (i.e. white spots, brittleness, change in moons)

Changes in voice

Changed sense of taste

Chills but no fever

Clogged ears

Cold burning feeling in lungs

Confusion

Congested or runny nose

Constant thirst

Costochondritis (inflammation of the cartilage that connects a rib to the breastbone)

Cough

Coughing up blood

Covid toes (tender or itchy rash or chilblains on the toes or foot)

| | | | | | |
|---|---|---|---|---|---|
| Cracked or dry lips | ○ | ○ | ○ | ○ | ○ |
| Dental problems (i.e. chipped tooth, tooth loss) | ○ | ○ | ○ | ○ | ○ |
| Diarrhea | ○ | ○ | ○ | ○ | ○ |
| Difficulty concentrating or focusing | ○ | ○ | ○ | ○ | ○ |
| Difficulty sleeping | ○ | ○ | ○ | ○ | ○ |
| Difficulty speaking properly | ○ | ○ | ○ | ○ | ○ |
| Discoloration of the skin (for example: purple or blue on the hands or feet, no blistering) | ○ | ○ | ○ | ○ | ○ |
| Dizziness | ○ | ○ | ○ | ○ | ○ |
| Dry eyes | ○ | ○ | ○ | ○ | ○ |
| Dry or peeling skin | ○ | ○ | ○ | ○ | ○ |
| Dry scalp or dandruff | ○ | ○ | ○ | ○ | ○ |
| Dry throat | ○ | ○ | ○ | ○ | ○ |
| Ear pain/earache | ○ | ○ | ○ | ○ | ○ |
| Elevated thyroid hormones | ○ | ○ | ○ | ○ | ○ |
| Extreme pressure at base of head or occipital nerve | ○ | ○ | ○ | ○ | ○ |
| Eye stye or infection | ○ | ○ | ○ | ○ | ○ |
| Fatigue | ○ | ○ | ○ | ○ | ○ |
| Feeling irritable | ○ | ○ | ○ | ○ | ○ |
| Feeling of burning skin | ○ | ○ | ○ | ○ | ○ |
| Fever or chills | ○ | ○ | ○ | ○ | ○ |
| Floaters or flashes of light in vision | ○ | ○ | ○ | ○ | ○ |
| Foot pain | ○ | ○ | ○ | ○ | ○ |
| GERD (acid reflux) with excessive salivation | ○ | ○ | ○ | ○ | ○ |
| Goiter or lump in throat | ○ | ○ | ○ | ○ | ○ |
| Hair loss | ○ | ○ | ○ | ○ | ○ |
| Hand or wrist pain | ○ | ○ | ○ | ○ | ○ |
| Headache | ○ | ○ | ○ | ○ | ○ |
| Heart palpitations (heart skipping a beat or racing) | ○ | ○ | ○ | ○ | ○ |
| Heat intolerance | ○ | ○ | ○ | ○ | ○ |
| High blood pressure | ○ | ○ | ○ | ○ | ○ |
| Hormone imbalances | ○ | ○ | ○ | ○ | ○ |
| "Hot" blood rush | ○ | ○ | ○ | ○ | ○ |
| Inability to cry | ○ | ○ | ○ | ○ | ○ |

| | | | | | |
|---|---|---|---|---|---|
| Inability to exercise or be active | ○ | ○ | ○ | ○ | ○ |
| Inability to yawn | ○ | ○ | ○ | ○ | ○ |
| Internal tremors or buzzing/vibration | ○ | ○ | ○ | ○ | ○ |
| Irregular or skipped menstrual cycles | ○ | ○ | ○ | ○ | ○ |
| Jaw pain | ○ | ○ | ○ | ○ | ○ |
| Joint pain | ○ | ○ | ○ | ○ | ○ |
| Kidney issues or protein in urine | ○ | ○ | ○ | ○ | ○ |
| Kidney pain | ○ | ○ | ○ | ○ | ○ |
| Low blood oxygen | ○ | ○ | ○ | ○ | ○ |
| Low blood pressure | ○ | ○ | ○ | ○ | ○ |
| Lower back pain | ○ | ○ | ○ | ○ | ○ |
| Loss of appetite | ○ | ○ | ○ | ○ | ○ |
| Loss of hearing | ○ | ○ | ○ | ○ | ○ |
| Loss or decrease in quality of vision | ○ | ○ | ○ | ○ | ○ |
| Lump in throat/difficulty swallowing | ○ | ○ | ○ | ○ | ○ |
| Memory problems | ○ | ○ | ○ | ○ | ○ |
| Menstrual cycles that are heavier or lighter than normal | ○ | ○ | ○ | ○ | ○ |
| Mid-back pain at base of ribs | ○ | ○ | ○ | ○ | ○ |
| Mouth sores or sore tongue | ○ | ○ | ○ | ○ | ○ |
| Muscle or body aches | ○ | ○ | ○ | ○ | ○ |
| Muscle twitching | ○ | ○ | ○ | ○ | ○ |
| Nausea or vomiting | ○ | ○ | ○ | ○ | ○ |
| Neck muscle pain | ○ | ○ | ○ | ○ | ○ |
| Nerve pain | ○ | ○ | ○ | ○ | ○ |
| Nerve sensations (tingling, pins and needles, numbness) | ○ | ○ | ○ | ○ | ○ |
| Neuropathy in feet and hands (weakness, numbness, and pain) | ○ | ○ | ○ | ○ | ○ |
| New allergies | ○ | ○ | ○ | ○ | ○ |
| Night sweats | ○ | ○ | ○ | ○ | ○ |
| Nightmares | ○ | ○ | ○ | ○ | ○ |
| Painful scalp | ○ | ○ | ○ | ○ | ○ |
| Partial or complete loss of sense of smell | ○ | ○ | ○ | ○ | ○ |
| Partial or complete loss of sense of taste | ○ | ○ | ○ | ○ | ○ |

| | | | | | |
|---|---|---|---|---|---|
| Persistent chest pain or pressure | ○ | ○ | ○ | ○ | ○ |
| Personality change (drastic) | ○ | ○ | ○ | ○ | ○ |
| Petechiae (pinpoint rash) | ○ | ○ | ○ | ○ | ○ |
| Phantom smells | ○ | ○ | ○ | ○ | ○ |
| Phlegm in back of throat | ○ | ○ | ○ | ○ | ○ |
| Post-exertional malaise (worsened symptoms or flu-like symptoms after exertion) | ○ | ○ | ○ | ○ | ○ |
| Postnasal drip | ○ | ○ | ○ | ○ | ○ |
| Rash | ○ | ○ | ○ | ○ | ○ |
| Reflux or heartburn | ○ | ○ | ○ | ○ | ○ |
| Runny nose | ○ | ○ | ○ | ○ | ○ |
| Sadness | ○ | ○ | ○ | ○ | ○ |
| Seizures | ○ | ○ | ○ | ○ | ○ |
| Sharp or sudden chest pain | ○ | ○ | ○ | ○ | ○ |
| Shortness of breath or difficulty breathing | ○ | ○ | ○ | ○ | ○ |
| Shortness of breath or exhaustion from bending over | ○ | ○ | ○ | ○ | ○ |
| Sleeping more than normal | ○ | ○ | ○ | ○ | ○ |
| Sore throat | ○ | ○ | ○ | ○ | ○ |
| Spinal issues | ○ | ○ | ○ | ○ | ○ |
| Spikes in blood pressure | ○ | ○ | ○ | ○ | ○ |
| Swollen hands or feet | ○ | ○ | ○ | ○ | ○ |
| Swollen lymph nodes | ○ | ○ | ○ | ○ | ○ |
| Syncope (fainting) | ○ | ○ | ○ | ○ | ○ |
| Tachycardia (rapid heartbeat) at rest | ○ | ○ | ○ | ○ | ○ |
| Tachycardia (rapid heartbeat) after standing up | ○ | ○ | ○ | ○ | ○ |
| Thrush (white fungal infection in the mouth or throat) | ○ | ○ | ○ | ○ | ○ |
| Tinnitus or humming in ears | ○ | ○ | ○ | ○ | ○ |
| Tremors or shakiness | ○ | ○ | ○ | ○ | ○ |
| Upper back pain | ○ | ○ | ○ | ○ | ○ |
| UTI (urinary tract infection) | ○ | ○ | ○ | ○ | ○ |
| Weakened neck | ○ | ○ | ○ | ○ | ○ |
| Weight gain | ○ | ○ | ○ | ○ | ○ |

**After quarantine, how much does/did the symptom impair your social or family functioning compared to pre-COVID?**

| | | | | | |
|---|---|---|---|---|---|
| Abdominal pain | ○ | ○ | ○ | ○ | ○ |
| Abnormally low temperature | ○ | ○ | ○ | ○ | ○ |
| Acid reflux | ○ | ○ | ○ | ○ | ○ |
| Afternoon or evening fevers/low-grade fevers | ○ | ○ | ○ | ○ | ○ |
| Anemia (low number of red blood cells) | ○ | ○ | ○ | ○ | ○ |
| Anxiety | ○ | ○ | ○ | ○ | ○ |
| Arrythmia (improper beating of the heart due to electrical impulse problems) | ○ | ○ | ○ | ○ | ○ |
| Bilateral neck throbbing around lymph nodes | ○ | ○ | ○ | ○ | ○ |
| Blurry vision | ○ | ○ | ○ | ○ | ○ |
| Bone aches in extremities | ○ | ○ | ○ | ○ | ○ |
| Brain fog | ○ | ○ | ○ | ○ | ○ |
| Brain pressure | ○ | ○ | ○ | ○ | ○ |
| Bulging veins | ○ | ○ | ○ | ○ | ○ |
| Burning sensations | ○ | ○ | ○ | ○ | ○ |
| Bruising of skin | ○ | ○ | ○ | ○ | ○ |
| Calf cramps | ○ | ○ | ○ | ○ | ○ |
| Change in nails (i.e. white spots, brittleness, change in moons) | ○ | ○ | ○ | ○ | ○ |
| Changes in voice | ○ | ○ | ○ | ○ | ○ |
| Changed sense of taste | ○ | ○ | ○ | ○ | ○ |
| Chills but no fever | ○ | ○ | ○ | ○ | ○ |
| Clogged ears | ○ | ○ | ○ | ○ | ○ |
| Cold burning feeling in lungs | ○ | ○ | ○ | ○ | ○ |
| Confusion | ○ | ○ | ○ | ○ | ○ |
| Congested or runny nose | ○ | ○ | ○ | ○ | ○ |
| Constant thirst | ○ | ○ | ○ | ○ | ○ |
| Costochondritis (inflammation of the cartilage that connects a rib to the breastbone) | ○ | ○ | ○ | ○ | ○ |
| Cough | ○ | ○ | ○ | ○ | ○ |
| Coughing up blood | ○ | ○ | ○ | ○ | ○ |
| Covid toes (tender or itchy rash or chilblains on the toes or foot) | ○ | ○ | ○ | ○ | ○ |

| | | | | | |
|---|---|---|---|---|---|
| Cracked or dry lips | ○ | ○ | ○ | ○ | ○ |
| Dental problems (i.e. chipped tooth, tooth loss) | ○ | ○ | ○ | ○ | ○ |
| Diarrhea | ○ | ○ | ○ | ○ | ○ |
| Difficulty concentrating or focusing | ○ | ○ | ○ | ○ | ○ |
| Difficulty sleeping | ○ | ○ | ○ | ○ | ○ |
| Difficulty speaking properly | ○ | ○ | ○ | ○ | ○ |
| Discoloration of the skin (for example: purple or blue on the hands or feet, no blistering) | ○ | ○ | ○ | ○ | ○ |
| Dizziness | ○ | ○ | ○ | ○ | ○ |
| Dry eyes | ○ | ○ | ○ | ○ | ○ |
| Dry or peeling skin | ○ | ○ | ○ | ○ | ○ |
| Dry scalp or dandruff | ○ | ○ | ○ | ○ | ○ |
| Dry throat | ○ | ○ | ○ | ○ | ○ |
| Ear pain/earache | ○ | ○ | ○ | ○ | ○ |
| Elevated thyroid hormones | ○ | ○ | ○ | ○ | ○ |
| Extreme pressure at base of head or occipital nerve | ○ | ○ | ○ | ○ | ○ |
| Eye stye or infection | ○ | ○ | ○ | ○ | ○ |
| Fatigue | ○ | ○ | ○ | ○ | ○ |
| Feeling irritable | ○ | ○ | ○ | ○ | ○ |
| Feeling of burning skin | ○ | ○ | ○ | ○ | ○ |
| Fever or chills | ○ | ○ | ○ | ○ | ○ |
| Floaters or flashes of light in vision | ○ | ○ | ○ | ○ | ○ |
| Foot pain | ○ | ○ | ○ | ○ | ○ |
| GERD (acid reflux) with excessive salivation | ○ | ○ | ○ | ○ | ○ |
| Goiter or lump in throat | ○ | ○ | ○ | ○ | ○ |
| Hair loss | ○ | ○ | ○ | ○ | ○ |
| Hand or wrist pain | ○ | ○ | ○ | ○ | ○ |
| Headache | ○ | ○ | ○ | ○ | ○ |
| Heart palpitations (heart skipping a beat or racing) | ○ | ○ | ○ | ○ | ○ |
| Heat intolerance | ○ | ○ | ○ | ○ | ○ |
| High blood pressure | ○ | ○ | ○ | ○ | ○ |
| Hormone imbalances | ○ | ○ | ○ | ○ | ○ |
| "Hot" blood rush | ○ | ○ | ○ | ○ | ○ |
| Inability to cry | ○ | ○ | ○ | ○ | ○ |

| | | | | | |
|---|---|---|---|---|---|
| Inability to exercise or be active | ○ | ○ | ○ | ○ | ○ |
| Inability to yawn | ○ | ○ | ○ | ○ | ○ |
| Internal tremors or buzzing/vibration | ○ | ○ | ○ | ○ | ○ |
| Irregular or skipped menstrual cycles | ○ | ○ | ○ | ○ | ○ |
| Jaw pain | ○ | ○ | ○ | ○ | ○ |
| Joint pain | ○ | ○ | ○ | ○ | ○ |
| Kidney issues or protein in urine | ○ | ○ | ○ | ○ | ○ |
| Kidney pain | ○ | ○ | ○ | ○ | ○ |
| Low blood oxygen | ○ | ○ | ○ | ○ | ○ |
| Low blood pressure | ○ | ○ | ○ | ○ | ○ |
| Lower back pain | ○ | ○ | ○ | ○ | ○ |
| Loss of appetite | ○ | ○ | ○ | ○ | ○ |
| Loss of hearing | ○ | ○ | ○ | ○ | ○ |
| Loss or decrease in quality of vision | ○ | ○ | ○ | ○ | ○ |
| Lump in throat/difficulty swallowing | ○ | ○ | ○ | ○ | ○ |
| Memory problems | ○ | ○ | ○ | ○ | ○ |
| Menstrual cycles that are heavier or lighter than normal | ○ | ○ | ○ | ○ | ○ |
| Mid-back pain at base of ribs | ○ | ○ | ○ | ○ | ○ |
| Mouth sores or sore tongue | ○ | ○ | ○ | ○ | ○ |
| Muscle or body aches | ○ | ○ | ○ | ○ | ○ |
| Muscle twitching | ○ | ○ | ○ | ○ | ○ |
| Nausea or vomiting | ○ | ○ | ○ | ○ | ○ |
| Neck muscle pain | ○ | ○ | ○ | ○ | ○ |
| Nerve pain | ○ | ○ | ○ | ○ | ○ |
| Nerve sensations (tingling, pins and needles, numbness) | ○ | ○ | ○ | ○ | ○ |
| Neuropathy in feet and hands (weakness, numbness, and pain) | ○ | ○ | ○ | ○ | ○ |
| New allergies | ○ | ○ | ○ | ○ | ○ |
| Night sweats | ○ | ○ | ○ | ○ | ○ |
| Nightmares | ○ | ○ | ○ | ○ | ○ |
| Painful scalp | ○ | ○ | ○ | ○ | ○ |
| Partial or complete loss of sense of smell | ○ | ○ | ○ | ○ | ○ |
| Partial or complete loss of sense of taste | ○ | ○ | ○ | ○ | ○ |

| | | | | | |
|---|---|---|---|---|---|
| Persistent chest pain or pressure | ○ | ○ | ○ | ○ | ○ |
| Personality change (drastic) | ○ | ○ | ○ | ○ | ○ |
| Petechiae (pinpoint rash) | ○ | ○ | ○ | ○ | ○ |
| Phantom smells | ○ | ○ | ○ | ○ | ○ |
| Phlegm in back of throat | ○ | ○ | ○ | ○ | ○ |
| Post-exertional malaise (worsened symptoms or flu-like symptoms after exertion) | ○ | ○ | ○ | ○ | ○ |
| Postnasal drip | ○ | ○ | ○ | ○ | ○ |
| Rash | ○ | ○ | ○ | ○ | ○ |
| Reflux or heartburn | ○ | ○ | ○ | ○ | ○ |
| Runny nose | ○ | ○ | ○ | ○ | ○ |
| Sadness | ○ | ○ | ○ | ○ | ○ |
| Seizures | ○ | ○ | ○ | ○ | ○ |
| Sharp or sudden chest pain | ○ | ○ | ○ | ○ | ○ |
| Shortness of breath or difficulty breathing | ○ | ○ | ○ | ○ | ○ |
| Shortness of breath or exhaustion from bending over | ○ | ○ | ○ | ○ | ○ |
| Sleeping more than normal | ○ | ○ | ○ | ○ | ○ |
| Sore throat | ○ | ○ | ○ | ○ | ○ |
| Spinal issues | ○ | ○ | ○ | ○ | ○ |
| Spikes in blood pressure | ○ | ○ | ○ | ○ | ○ |
| Swollen hands or feet | ○ | ○ | ○ | ○ | ○ |
| Swollen lymph nodes | ○ | ○ | ○ | ○ | ○ |
| Syncope (fainting) | ○ | ○ | ○ | ○ | ○ |
| Tachycardia (rapid heartbeat) at rest | ○ | ○ | ○ | ○ | ○ |
| Tachycardia (rapid heartbeat) after standing up | ○ | ○ | ○ | ○ | ○ |
| Thrush (white fungal infection in the mouth or throat) | ○ | ○ | ○ | ○ | ○ |
| Tinnitus or humming in ears | ○ | ○ | ○ | ○ | ○ |
| Tremors or shakiness | ○ | ○ | ○ | ○ | ○ |
| Upper back pain | ○ | ○ | ○ | ○ | ○ |
| UTI (urinary tract infection) | ○ | ○ | ○ | ○ | ○ |
| Weakened neck | ○ | ○ | ○ | ○ | ○ |
| Weight gain | ○ | ○ | ○ | ○ | ○ |

---

What treatments (medicine, therapy, vitamins, body positioning, at-home care, etc.) have you found effective for reducing your symptoms, if any? ______________________________

| | 76-100% of health before COVID-19 | 51-75% of health before COVID-19 | 26-50% of health before COVID-19 | 0-25% of health before COVID-19 |
|---|---|---|---|---|
| On your best days, would you say you that you are at | ○ | ○ | ○ | ○ |
| On your worst days, would you say that you are at | ○ | ○ | ○ | ○ |

Please, in the space below, share any information about your experience with COVID that we might have missed. ________________________________________

### Survey 2

Thank you for taking Survey 2 for the Yale COVID Recovery Study!

Please remember that you may stop this survey and come back to it at any point. To return to the survey, please return to the link in your browser (or reclick the link in your email).

You may also find it helpful to have the below items ready as you complete the survey. If you do not have these items, please still fill in the information as best you can.

Testing results: dates, type (PCR, antigen) and result of tests. If you do not remember the exact date, the estimated date is enough.Symptom time and severity: your symptom log. You will be asked to pick symptoms you have had since you began having COVID-19 symptoms.

| | |
|---|---|
| Please verify that you received the first dose of your vaccine on [vaccine_arm_1][v_v_vax_date] | ◯ This is correct<br>◯ This is not correct ______ |
| On what date did you receive the first dose of your COVID-19 vaccine? | __________________________________ |
| Which COVID-19 vaccine did you receive? | ◯ Pfizer-BioNTech<br>◯ Moderna<br>◯ Johnson & Johnson<br>◯ AstraZeneca or Covishield<br>◯ Other<br>◯ Don't know |
| Have you received your second dose of the vaccine? | ◯ Yes ______<br>◯ No, but it's scheduled<br>◯ No, and I don't have an appointment scheduled<br>◯ No, but the vaccine I received requires only 1 dose |
| When did you receive your second dose of the COVID-19 vaccine? | __________________________________ |

Before you received the vaccine, you completed Survey 1 and marked the symptoms that you had experienced. Now, we will ask you a series of questions about these symptoms, and how they have changed (if at all). Please answer these questions based on your experience since you received the COVID-19 vaccine, or for the time period from [vaccine_arm_1][v_v_vax_date] until now.

**Since [vaccine_arm_1][v_v_vax_date], this symptom has been**

| | Better | Worse | The same |
|---|---|---|---|
| Abdominal pain | ○ | ○ | ○ |
| Abnormally low temperature | ○ | ○ | ○ |
| Acid reflux | ○ | ○ | ○ |
| Afternoon or evening fevers/low-grade fevers | ○ | ○ | ○ |
| Anemia (low number of red blood cells) | ○ | ○ | ○ |
| Anxiety | ○ | ○ | ○ |
| Arrythmia (improper beating of the heart due to electrical impulse problems) | ○ | ○ | ○ |
| Bilateral neck throbbing around lymph nodes | ○ | ○ | ○ |
| Blurry vision | ○ | ○ | ○ |
| Bone aches in extremities | ○ | ○ | ○ |
| Brain fog | ○ | ○ | ○ |
| Brain pressure | ○ | ○ | ○ |
| Bulging veins | ○ | ○ | ○ |
| Burning sensations | ○ | ○ | ○ |
| Bruising of skin | ○ | ○ | ○ |
| Calf cramps | ○ | ○ | ○ |
| Change in nails (i.e. white spots, brittleness, change in moons) | ○ | ○ | ○ |
| Changes in voice | ○ | ○ | ○ |
| Changed sense of taste | ○ | ○ | ○ |
| Chills but no fever | ○ | ○ | ○ |
| Clogged ears | ○ | ○ | ○ |
| Cold burning feeling in lungs | ○ | ○ | ○ |
| Confusion | ○ | ○ | ○ |
| Congested or runny nose | ○ | ○ | ○ |
| Constant thirst | ○ | ○ | ○ |
| Costochondritis (inflammation of the cartilage that connects a rib to the breastbone) | ○ | ○ | ○ |
| Cough | ○ | ○ | ○ |
| Coughing up blood | ○ | ○ | ○ |
| Covid toes (tender or itchy rash or chilblains on the toes or foot) | ○ | ○ | ○ |

| | | | |
|---|---|---|---|
| Cracked or dry lips | ○ | ○ | ○ |
| Dental problems (i.e. chipped tooth, tooth loss) | ○ | ○ | ○ |
| Diarrhea | ○ | ○ | ○ |
| Difficulty concentrating or focusing | ○ | ○ | ○ |
| Difficulty sleeping | ○ | ○ | ○ |
| Difficulty speaking properly | ○ | ○ | ○ |
| Discoloration of the skin (for example: purple or blue on the hands or feet, no blistering) | ○ | ○ | ○ |
| Dizziness | ○ | ○ | ○ |
| Dry eyes | ○ | ○ | ○ |
| Dry or peeling skin | ○ | ○ | ○ |
| Dry scalp or dandruff | ○ | ○ | ○ |
| Dry throat | ○ | ○ | ○ |
| Ear pain/earache | ○ | ○ | ○ |
| Elevated thyroid hormones | ○ | ○ | ○ |
| Extreme pressure at base of head or occipital nerve | ○ | ○ | ○ |
| Eye stye or infection | ○ | ○ | ○ |
| Fatigue | ○ | ○ | ○ |
| Feeling irritable | ○ | ○ | ○ |
| Feeling of burning skin | ○ | ○ | ○ |
| Fever or chills | ○ | ○ | ○ |
| Floaters or flashes of light in vision | ○ | ○ | ○ |
| Foot pain | ○ | ○ | ○ |
| GERD (acid reflux) with excessive salivation | ○ | ○ | ○ |
| Goiter or lump in throat | ○ | ○ | ○ |
| Hair loss | ○ | ○ | ○ |
| Hand or wrist pain | ○ | ○ | ○ |
| Headache | ○ | ○ | ○ |
| Heart palpitations (heart skipping a beat or racing) | ○ | ○ | ○ |
| Heat intolerance | ○ | ○ | ○ |
| High blood pressure | ○ | ○ | ○ |
| Hormone imbalances | ○ | ○ | ○ |
| "Hot" blood rush | ○ | ○ | ○ |
| Inability to cry | ○ | ○ | ○ |

| | | | |
|---|---|---|---|
| Inability to exercise or be active | ○ | ○ | ○ |
| Inability to yawn | ○ | ○ | ○ |
| Internal tremors or buzzing/vibration | ○ | ○ | ○ |
| Irregular or skipped menstrual cycles | ○ | ○ | ○ |
| Jaw pain | ○ | ○ | ○ |
| Joint pain | ○ | ○ | ○ |
| Kidney issues or protein in urine | ○ | ○ | ○ |
| Kidney pain | ○ | ○ | ○ |
| Low blood oxygen | ○ | ○ | ○ |
| Low blood pressure | ○ | ○ | ○ |
| Lower back pain | ○ | ○ | ○ |
| Loss of appetite | ○ | ○ | ○ |
| Loss of hearing | ○ | ○ | ○ |
| Loss or decrease in quality of vision | ○ | ○ | ○ |
| Lump in throat/difficulty swallowing | ○ | ○ | ○ |
| Memory problems | ○ | ○ | ○ |
| Menstrual cycles that are heavier or lighter than normal | ○ | ○ | ○ |
| Mid-back pain at base of ribs | ○ | ○ | ○ |
| Mouth sores or sore tongue | ○ | ○ | ○ |
| Muscle or body aches | ○ | ○ | ○ |
| Muscle twitching | ○ | ○ | ○ |
| Nausea or vomiting | ○ | ○ | ○ |
| Neck muscle pain | ○ | ○ | ○ |
| Nerve pain | ○ | ○ | ○ |
| Nerve sensations (tingling, pins and needles, numbness) | ○ | ○ | ○ |
| Neuropathy in feet and hands (weakness, numbness, and pain) | ○ | ○ | ○ |
| New allergies | ○ | ○ | ○ |
| Night sweats | ○ | ○ | ○ |
| Nightmares | ○ | ○ | ○ |
| Painful scalp | ○ | ○ | ○ |
| Partial or complete loss of sense of smell | ○ | ○ | ○ |
| Partial or complete loss of sense of taste | ○ | ○ | ○ |

| | | | |
|---|---|---|---|
| Persistent chest pain or pressure | ○ | ○ | ○ |
| Personality change (drastic) | ○ | ○ | ○ |
| Petechiae (pinpoint rash) | ○ | ○ | ○ |
| Phantom smells | ○ | ○ | ○ |
| Phlegm in back of throat | ○ | ○ | ○ |
| Post-exertional malaise (worsened symptoms or flu-like symptoms after exertion) | ○ | ○ | ○ |
| Postnasal drip | ○ | ○ | ○ |
| Rash | ○ | ○ | ○ |
| Reflux or heartburn | ○ | ○ | ○ |
| Runny nose | ○ | ○ | ○ |
| Sadness | ○ | ○ | ○ |
| Seizures | ○ | ○ | ○ |
| Sharp or sudden chest pain | ○ | ○ | ○ |
| Shortness of breath or difficulty breathing | ○ | ○ | ○ |
| Shortness of breath or exhaustion from bending over | ○ | ○ | ○ |
| Sleeping more than normal | ○ | ○ | ○ |
| Sore throat | ○ | ○ | ○ |
| Spinal issues | ○ | ○ | ○ |
| Spikes in blood pressure | ○ | ○ | ○ |
| Swollen hands or feet | ○ | ○ | ○ |
| Swollen lymph nodes | ○ | ○ | ○ |
| Syncope (fainting) | ○ | ○ | ○ |
| Tachycardia (rapid heartbeat) at rest | ○ | ○ | ○ |
| Tachycardia (rapid heartbeat) after standing up | ○ | ○ | ○ |
| Thrush (white fungal infection in the mouth or throat) | ○ | ○ | ○ |
| Tinnitus or humming in ears | ○ | ○ | ○ |
| Tremors or shakiness | ○ | ○ | ○ |
| Upper back pain | ○ | ○ | ○ |
| UTI (urinary tract infection) | ○ | ○ | ○ |
| Weakened neck | ○ | ○ | ○ |
| Weight gain | ○ | ○ | ○ |

**Since [vaccine_arm_1][v_v_vax_date], I have experienced this symptom, on average**

| | Never--it has gone away completely | Once a month | Once or twice a week | 3 or 4 times a week | Most days | All the time |
|---|---|---|---|---|---|---|
| Abdominal pain | ○ | ○ | ○ | ○ | ○ | ○ |
| Abnormally low temperature | ○ | ○ | ○ | ○ | ○ | ○ |
| Acid reflux | ○ | ○ | ○ | ○ | ○ | ○ |
| Afternoon or evening fevers/low-grade fevers | ○ | ○ | ○ | ○ | ○ | ○ |
| Anemia (low number of red blood cells) | ○ | ○ | ○ | ○ | ○ | ○ |
| Anxiety | ○ | ○ | ○ | ○ | ○ | ○ |
| Arrythmia (improper beating of the heart due to electrical impulse problems) | ○ | ○ | ○ | ○ | ○ | ○ |
| Bilateral neck throbbing around lymph nodes | ○ | ○ | ○ | ○ | ○ | ○ |
| Blurry vision | ○ | ○ | ○ | ○ | ○ | ○ |
| Bone aches in extremities | ○ | ○ | ○ | ○ | ○ | ○ |
| Brain fog | ○ | ○ | ○ | ○ | ○ | ○ |
| Brain pressure | ○ | ○ | ○ | ○ | ○ | ○ |
| Bulging veins | ○ | ○ | ○ | ○ | ○ | ○ |
| Burning sensations | ○ | ○ | ○ | ○ | ○ | ○ |
| Bruising of skin | ○ | ○ | ○ | ○ | ○ | ○ |
| Calf cramps | ○ | ○ | ○ | ○ | ○ | ○ |
| Change in nails (i.e. white spots, brittleness, change in moons) | ○ | ○ | ○ | ○ | ○ | ○ |
| Changes in voice | ○ | ○ | ○ | ○ | ○ | ○ |
| Changed sense of taste | ○ | ○ | ○ | ○ | ○ | ○ |
| Chills but no fever | ○ | ○ | ○ | ○ | ○ | ○ |
| Clogged ears | ○ | ○ | ○ | ○ | ○ | ○ |
| Cold burning feeling in lungs | ○ | ○ | ○ | ○ | ○ | ○ |
| Confusion | ○ | ○ | ○ | ○ | ○ | ○ |
| Congested or runny nose | ○ | ○ | ○ | ○ | ○ | ○ |
| Constant thirst | ○ | ○ | ○ | ○ | ○ | ○ |
| Costochondritis (inflammation of the cartilage that connects a rib to the breastbone) | ○ | ○ | ○ | ○ | ○ | ○ |
| Cough | ○ | ○ | ○ | ○ | ○ | ○ |
| Coughing up blood | ○ | ○ | ○ | ○ | ○ | ○ |

| | | | | | | |
|---|---|---|---|---|---|---|
| Covid toes (tender or itchy rash or chilblains on the toes or foot) | ◯ | ◯ | ◯ | ◯ | ◯ | ◯ |
| Cracked or dry lips | ◯ | ◯ | ◯ | ◯ | ◯ | ◯ |
| Dental problems (i.e. chipped tooth, tooth loss) | ◯ | ◯ | ◯ | ◯ | ◯ | ◯ |
| Diarrhea | ◯ | ◯ | ◯ | ◯ | ◯ | ◯ |
| Difficulty concentrating or focusing | ◯ | ◯ | ◯ | ◯ | ◯ | ◯ |
| Difficulty sleeping | ◯ | ◯ | ◯ | ◯ | ◯ | ◯ |
| Difficulty speaking properly | ◯ | ◯ | ◯ | ◯ | ◯ | ◯ |
| Discoloration of the skin (for example: purple or blue on the hands or feet, no blistering) | ◯ | ◯ | ◯ | ◯ | ◯ | ◯ |
| Dizziness | ◯ | ◯ | ◯ | ◯ | ◯ | ◯ |
| Dry eyes | ◯ | ◯ | ◯ | ◯ | ◯ | ◯ |
| Dry or peeling skin | ◯ | ◯ | ◯ | ◯ | ◯ | ◯ |
| Dry scalp or dandruff | ◯ | ◯ | ◯ | ◯ | ◯ | ◯ |
| Dry throat | ◯ | ◯ | ◯ | ◯ | ◯ | ◯ |
| Ear pain/earache | ◯ | ◯ | ◯ | ◯ | ◯ | ◯ |
| Elevated thyroid hormones | ◯ | ◯ | ◯ | ◯ | ◯ | ◯ |
| Extreme pressure at base of head or occipital nerve | ◯ | ◯ | ◯ | ◯ | ◯ | ◯ |
| Eye stye or infection | ◯ | ◯ | ◯ | ◯ | ◯ | ◯ |
| Fatigue | ◯ | ◯ | ◯ | ◯ | ◯ | ◯ |
| Feeling irritable | ◯ | ◯ | ◯ | ◯ | ◯ | ◯ |
| Feeling of burning skin | ◯ | ◯ | ◯ | ◯ | ◯ | ◯ |
| Fever or chills | ◯ | ◯ | ◯ | ◯ | ◯ | ◯ |
| Floaters or flashes of light in vision | ◯ | ◯ | ◯ | ◯ | ◯ | ◯ |
| Foot pain | ◯ | ◯ | ◯ | ◯ | ◯ | ◯ |
| GERD (acid reflux) with excessive salivation | ◯ | ◯ | ◯ | ◯ | ◯ | ◯ |
| Goiter or lump in throat | ◯ | ◯ | ◯ | ◯ | ◯ | ◯ |
| Hair loss | ◯ | ◯ | ◯ | ◯ | ◯ | ◯ |
| Hand or wrist pain | ◯ | ◯ | ◯ | ◯ | ◯ | ◯ |
| Headache | ◯ | ◯ | ◯ | ◯ | ◯ | ◯ |
| Heart palpitations (heart skipping a beat or racing) | ◯ | ◯ | ◯ | ◯ | ◯ | ◯ |
| Heat intolerance | ◯ | ◯ | ◯ | ◯ | ◯ | ◯ |
| High blood pressure | ◯ | ◯ | ◯ | ◯ | ◯ | ◯ |
| Hormone imbalances | ◯ | ◯ | ◯ | ◯ | ◯ | ◯ |

| | | | | | | |
|---|---|---|---|---|---|---|
| "Hot" blood rush | ○ | ○ | ○ | ○ | ○ | ○ |
| Inability to cry | ○ | ○ | ○ | ○ | ○ | ○ |
| Inability to exercise or be active | ○ | ○ | ○ | ○ | ○ | ○ |
| Inability to yawn | ○ | ○ | ○ | ○ | ○ | ○ |
| Internal tremors or buzzing/vibration | ○ | ○ | ○ | ○ | ○ | ○ |
| Irregular or skipped menstrual cycles | ○ | ○ | ○ | ○ | ○ | ○ |
| Jaw pain | ○ | ○ | ○ | ○ | ○ | ○ |
| Joint pain | ○ | ○ | ○ | ○ | ○ | ○ |
| Kidney issues or protein in urine | ○ | ○ | ○ | ○ | ○ | ○ |
| Kidney pain | ○ | ○ | ○ | ○ | ○ | ○ |
| Low blood oxygen | ○ | ○ | ○ | ○ | ○ | ○ |
| Low blood pressure | ○ | ○ | ○ | ○ | ○ | ○ |
| Lower back pain | ○ | ○ | ○ | ○ | ○ | ○ |
| Loss of appetite | ○ | ○ | ○ | ○ | ○ | ○ |
| Loss of hearing | ○ | ○ | ○ | ○ | ○ | ○ |
| Loss or decrease in quality of vision | ○ | ○ | ○ | ○ | ○ | ○ |
| Lump in throat/difficulty swallowing | ○ | ○ | ○ | ○ | ○ | ○ |
| Memory problems | ○ | ○ | ○ | ○ | ○ | ○ |
| Menstrual cycles that are heavier or lighter than normal | ○ | ○ | ○ | ○ | ○ | ○ |
| Mid-back pain at base of ribs | ○ | ○ | ○ | ○ | ○ | ○ |
| Mouth sores or sore tongue | ○ | ○ | ○ | ○ | ○ | ○ |
| Muscle or body aches | ○ | ○ | ○ | ○ | ○ | ○ |
| Muscle twitching | ○ | ○ | ○ | ○ | ○ | ○ |
| Nausea or vomiting | ○ | ○ | ○ | ○ | ○ | ○ |
| Neck muscle pain | ○ | ○ | ○ | ○ | ○ | ○ |
| Nerve pain | ○ | ○ | ○ | ○ | ○ | ○ |
| Nerve sensations (tingling, pins and needles, numbness) | ○ | ○ | ○ | ○ | ○ | ○ |
| Neuropathy in feet and hands (weakness, numbness, and pain) | ○ | ○ | ○ | ○ | ○ | ○ |
| New allergies | ○ | ○ | ○ | ○ | ○ | ○ |
| Night sweats | ○ | ○ | ○ | ○ | ○ | ○ |
| Nightmares | ○ | ○ | ○ | ○ | ○ | ○ |
| Painful scalp | ○ | ○ | ○ | ○ | ○ | ○ |
| Partial or complete loss of sense of smell | ○ | ○ | ○ | ○ | ○ | ○ |

| | | | | | | |
|---|---|---|---|---|---|---|
| Partial or complete loss of sense of taste | ○ | ○ | ○ | ○ | ○ | ○ |
| Persistent chest pain or pressure | ○ | ○ | ○ | ○ | ○ | ○ |
| Personality change (drastic) | ○ | ○ | ○ | ○ | ○ | ○ |
| Petechiae (pinpoint rash) | ○ | ○ | ○ | ○ | ○ | ○ |
| Phantom smells | ○ | ○ | ○ | ○ | ○ | ○ |
| Phlegm in back of throat | ○ | ○ | ○ | ○ | ○ | ○ |
| Post-exertional malaise (worsened symptoms or flu-like symptoms after exertion) | ○ | ○ | ○ | ○ | ○ | ○ |
| Postnasal drip | ○ | ○ | ○ | ○ | ○ | ○ |
| Rash | ○ | ○ | ○ | ○ | ○ | ○ |
| Reflux or heartburn | ○ | ○ | ○ | ○ | ○ | ○ |
| Runny nose | ○ | ○ | ○ | ○ | ○ | ○ |
| Sadness | ○ | ○ | ○ | ○ | ○ | ○ |
| Seizures | ○ | ○ | ○ | ○ | ○ | ○ |
| Sharp or sudden chest pain | ○ | ○ | ○ | ○ | ○ | ○ |
| Shortness of breath or difficulty breathing | ○ | ○ | ○ | ○ | ○ | ○ |
| Shortness of breath or exhaustion from bending over | ○ | ○ | ○ | ○ | ○ | ○ |
| Sleeping more than normal | ○ | ○ | ○ | ○ | ○ | ○ |
| Sore throat | ○ | ○ | ○ | ○ | ○ | ○ |
| Spinal issues | ○ | ○ | ○ | ○ | ○ | ○ |
| Spikes in blood pressure | ○ | ○ | ○ | ○ | ○ | ○ |
| Swollen hands or feet | ○ | ○ | ○ | ○ | ○ | ○ |
| Swollen lymph nodes | ○ | ○ | ○ | ○ | ○ | ○ |
| Syncope (fainting) | ○ | ○ | ○ | ○ | ○ | ○ |
| Tachycardia (rapid heartbeat) at rest | ○ | ○ | ○ | ○ | ○ | ○ |
| Tachycardia (rapid heartbeat) after standing up | ○ | ○ | ○ | ○ | ○ | ○ |
| Thrush (white fungal infection in the mouth or throat) | ○ | ○ | ○ | ○ | ○ | ○ |
| Tinnitus or humming in ears | ○ | ○ | ○ | ○ | ○ | ○ |
| Tremors or shakiness | ○ | ○ | ○ | ○ | ○ | ○ |
| Upper back pain | ○ | ○ | ○ | ○ | ○ | ○ |
| UTI (urinary tract infection) | ○ | ○ | ○ | ○ | ○ | ○ |
| Weakened neck | ○ | ○ | ○ | ○ | ○ | ○ |
| Weight gain | ○ | ○ | ○ | ○ | ○ | ○ |

**Since [vaccine_arm_1][v_v_vax_date], while experiencing the symptom, how much did it bother you in terms of discomfort or pain?**

***If you have not experienced the symptom since your vaccine, please choose "Not at all"**

| | Not at all | A little bit | Somewhat | Quite a bit | Very much |
|---|---|---|---|---|---|
| Abdominal pain | ○ | ○ | ○ | ○ | ○ |
| Abnormally low temperature | ○ | ○ | ○ | ○ | ○ |
| Acid reflux | ○ | ○ | ○ | ○ | ○ |
| Afternoon or evening fevers/low-grade fevers | ○ | ○ | ○ | ○ | ○ |
| Anemia (low number of red blood cells) | ○ | ○ | ○ | ○ | ○ |
| Anxiety | ○ | ○ | ○ | ○ | ○ |
| Arrythmia (improper beating of the heart due to electrical impulse problems) | ○ | ○ | ○ | ○ | ○ |
| Bilateral neck throbbing around lymph nodes | ○ | ○ | ○ | ○ | ○ |
| Blurry vision | ○ | ○ | ○ | ○ | ○ |
| Bone aches in extremities | ○ | ○ | ○ | ○ | ○ |
| Brain fog | ○ | ○ | ○ | ○ | ○ |
| Brain pressure | ○ | ○ | ○ | ○ | ○ |
| Bulging veins | ○ | ○ | ○ | ○ | ○ |
| Burning sensations | ○ | ○ | ○ | ○ | ○ |
| Bruising of skin | ○ | ○ | ○ | ○ | ○ |
| Calf cramps | ○ | ○ | ○ | ○ | ○ |
| Change in nails (i.e. white spots, brittleness, change in moons) | ○ | ○ | ○ | ○ | ○ |
| Changes in voice | ○ | ○ | ○ | ○ | ○ |
| Changed sense of taste | ○ | ○ | ○ | ○ | ○ |
| Chills but no fever | ○ | ○ | ○ | ○ | ○ |
| Clogged ears | ○ | ○ | ○ | ○ | ○ |
| Cold burning feeling in lungs | ○ | ○ | ○ | ○ | ○ |
| Confusion | ○ | ○ | ○ | ○ | ○ |
| Congested or runny nose | ○ | ○ | ○ | ○ | ○ |
| Constant thirst | ○ | ○ | ○ | ○ | ○ |
| Costochondritis (inflammation of the cartilage that connects a rib to the breastbone) | ○ | ○ | ○ | ○ | ○ |
| Cough | ○ | ○ | ○ | ○ | ○ |

| | | | | | |
|---|---|---|---|---|---|
| Coughing up blood | ○ | ○ | ○ | ○ | ○ |
| Covid toes (tender or itchy rash or chilblains on the toes or foot) | ○ | ○ | ○ | ○ | ○ |
| Cracked or dry lips | ○ | ○ | ○ | ○ | ○ |
| Dental problems (i.e. chipped tooth, tooth loss) | ○ | ○ | ○ | ○ | ○ |
| Diarrhea | ○ | ○ | ○ | ○ | ○ |
| Difficulty concentrating or focusing | ○ | ○ | ○ | ○ | ○ |
| Difficulty sleeping | ○ | ○ | ○ | ○ | ○ |
| Difficulty speaking properly | ○ | ○ | ○ | ○ | ○ |
| Discoloration of the skin (for example: purple or blue on the hands or feet, no blistering) | ○ | ○ | ○ | ○ | ○ |
| Dizziness | ○ | ○ | ○ | ○ | ○ |
| Dry eyes | ○ | ○ | ○ | ○ | ○ |
| Dry or peeling skin | ○ | ○ | ○ | ○ | ○ |
| Dry scalp or dandruff | ○ | ○ | ○ | ○ | ○ |
| Dry throat | ○ | ○ | ○ | ○ | ○ |
| Ear pain/earache | ○ | ○ | ○ | ○ | ○ |
| Elevated thyroid hormones | ○ | ○ | ○ | ○ | ○ |
| Extreme pressure at base of head or occipital nerve | ○ | ○ | ○ | ○ | ○ |
| Eye stye or infection | ○ | ○ | ○ | ○ | ○ |
| Fatigue | ○ | ○ | ○ | ○ | ○ |
| Feeling irritable | ○ | ○ | ○ | ○ | ○ |
| Feeling of burning skin | ○ | ○ | ○ | ○ | ○ |
| Fever or chills | ○ | ○ | ○ | ○ | ○ |
| Floaters or flashes of light in vision | ○ | ○ | ○ | ○ | ○ |
| Foot pain | ○ | ○ | ○ | ○ | ○ |
| GERD (acid reflux) with excessive salivation | ○ | ○ | ○ | ○ | ○ |
| Goiter or lump in throat | ○ | ○ | ○ | ○ | ○ |
| Hair loss | ○ | ○ | ○ | ○ | ○ |
| Hand or wrist pain | ○ | ○ | ○ | ○ | ○ |
| Headache | ○ | ○ | ○ | ○ | ○ |
| Heart palpitations (heart skipping a beat or racing) | ○ | ○ | ○ | ○ | ○ |
| Heat intolerance | ○ | ○ | ○ | ○ | ○ |
| High blood pressure | ○ | ○ | ○ | ○ | ○ |

| | | | | | |
|---|---|---|---|---|---|
| Hormone imbalances | ○ | ○ | ○ | ○ | ○ |
| "Hot" blood rush | ○ | ○ | ○ | ○ | ○ |
| Inability to cry | ○ | ○ | ○ | ○ | ○ |
| Inability to exercise or be active | ○ | ○ | ○ | ○ | ○ |
| Inability to yawn | ○ | ○ | ○ | ○ | ○ |
| Internal tremors or buzzing/vibration | ○ | ○ | ○ | ○ | ○ |
| Irregular or skipped menstrual cycles | ○ | ○ | ○ | ○ | ○ |
| Jaw pain | ○ | ○ | ○ | ○ | ○ |
| Joint pain | ○ | ○ | ○ | ○ | ○ |
| Kidney issues or protein in urine | ○ | ○ | ○ | ○ | ○ |
| Kidney pain | ○ | ○ | ○ | ○ | ○ |
| Low blood oxygen | ○ | ○ | ○ | ○ | ○ |
| Low blood pressure | ○ | ○ | ○ | ○ | ○ |
| Lower back pain | ○ | ○ | ○ | ○ | ○ |
| Loss of appetite | ○ | ○ | ○ | ○ | ○ |
| Loss of hearing | ○ | ○ | ○ | ○ | ○ |
| Loss or decrease in quality of vision | ○ | ○ | ○ | ○ | ○ |
| Lump in throat/difficulty swallowing | ○ | ○ | ○ | ○ | ○ |
| Memory problems | ○ | ○ | ○ | ○ | ○ |
| Menstrual cycles that are heavier or lighter than normal | ○ | ○ | ○ | ○ | ○ |
| Mid-back pain at base of ribs | ○ | ○ | ○ | ○ | ○ |
| Mouth sores or sore tongue | ○ | ○ | ○ | ○ | ○ |
| Muscle or body aches | ○ | ○ | ○ | ○ | ○ |
| Muscle twitching | ○ | ○ | ○ | ○ | ○ |
| Nausea or vomiting | ○ | ○ | ○ | ○ | ○ |
| Neck muscle pain | ○ | ○ | ○ | ○ | ○ |
| Nerve pain | ○ | ○ | ○ | ○ | ○ |
| Nerve sensations (tingling, pins and needles, numbness) | ○ | ○ | ○ | ○ | ○ |
| Neuropathy in feet and hands (weakness, numbness, and pain) | ○ | ○ | ○ | ○ | ○ |
| New allergies | ○ | ○ | ○ | ○ | ○ |
| Night sweats | ○ | ○ | ○ | ○ | ○ |
| Nightmares | ○ | ○ | ○ | ○ | ○ |
| Painful scalp | ○ | ○ | ○ | ○ | ○ |

| | | | | | |
|---|---|---|---|---|---|
| Partial or complete loss of sense of smell | ◯ | ◯ | ◯ | ◯ | ◯ |
| Partial or complete loss of sense of taste | ◯ | ◯ | ◯ | ◯ | ◯ |
| Persistent chest pain or pressure | ◯ | ◯ | ◯ | ◯ | ◯ |
| Personality change (drastic) | ◯ | ◯ | ◯ | ◯ | ◯ |
| Petechiae (pinpoint rash) | ◯ | ◯ | ◯ | ◯ | ◯ |
| Phantom smells | ◯ | ◯ | ◯ | ◯ | ◯ |
| Phlegm in back of throat | ◯ | ◯ | ◯ | ◯ | ◯ |
| Post-exertional malaise (worsened symptoms or flu-like symptoms after exertion) | ◯ | ◯ | ◯ | ◯ | ◯ |
| Postnasal drip | ◯ | ◯ | ◯ | ◯ | ◯ |
| Rash | ◯ | ◯ | ◯ | ◯ | ◯ |
| Reflux or heartburn | ◯ | ◯ | ◯ | ◯ | ◯ |
| Runny nose | ◯ | ◯ | ◯ | ◯ | ◯ |
| Sadness | ◯ | ◯ | ◯ | ◯ | ◯ |
| Seizures | ◯ | ◯ | ◯ | ◯ | ◯ |
| Sharp or sudden chest pain | ◯ | ◯ | ◯ | ◯ | ◯ |
| Shortness of breath or difficulty breathing | ◯ | ◯ | ◯ | ◯ | ◯ |
| Shortness of breath or exhaustion from bending over | ◯ | ◯ | ◯ | ◯ | ◯ |
| Sleeping more than normal | ◯ | ◯ | ◯ | ◯ | ◯ |
| Sore throat | ◯ | ◯ | ◯ | ◯ | ◯ |
| Spinal issues | ◯ | ◯ | ◯ | ◯ | ◯ |
| Spikes in blood pressure | ◯ | ◯ | ◯ | ◯ | ◯ |
| Swollen hands or feet | ◯ | ◯ | ◯ | ◯ | ◯ |
| Swollen lymph nodes | ◯ | ◯ | ◯ | ◯ | ◯ |
| Syncope (fainting) | ◯ | ◯ | ◯ | ◯ | ◯ |
| Tachycardia (rapid heartbeat) at rest | ◯ | ◯ | ◯ | ◯ | ◯ |
| Tachycardia (rapid heartbeat) after standing up | ◯ | ◯ | ◯ | ◯ | ◯ |
| Thrush (white fungal infection in the mouth or throat) | ◯ | ◯ | ◯ | ◯ | ◯ |
| Tinnitus or humming in ears | ◯ | ◯ | ◯ | ◯ | ◯ |
| Tremors or shakiness | ◯ | ◯ | ◯ | ◯ | ◯ |
| Upper back pain | ◯ | ◯ | ◯ | ◯ | ◯ |
| UTI (urinary tract infection) | ◯ | ◯ | ◯ | ◯ | ◯ |
| Weakened neck | ◯ | ◯ | ◯ | ◯ | ◯ |

| | | | | | |
|---|---|---|---|---|---|
| Weight gain | ○ | ○ | ○ | ○ | ○ |

**Since [vaccine_arm_1][v_v_vax_date], while experiencing the symptom, how much did it impair your ability to work compared to pre-COVID?**

***If you have not experienced the symptom since your vaccine, please choose "Not at all."**

| | Not at all | A little bit | Somewhat | Quite a bit | Very much |
|---|---|---|---|---|---|
| Abdominal pain | ○ | ○ | ○ | ○ | ○ |
| Abnormally low temperature | ○ | ○ | ○ | ○ | ○ |
| Acid reflux | ○ | ○ | ○ | ○ | ○ |
| Afternoon or evening fevers/low-grade fevers | ○ | ○ | ○ | ○ | ○ |
| Anemia (low number of red blood cells) | ○ | ○ | ○ | ○ | ○ |
| Anxiety | ○ | ○ | ○ | ○ | ○ |
| Arrythmia (improper beating of the heart due to electrical impulse problems) | ○ | ○ | ○ | ○ | ○ |
| Bilateral neck throbbing around lymph nodes | ○ | ○ | ○ | ○ | ○ |
| Blurry vision | ○ | ○ | ○ | ○ | ○ |
| Bone aches in extremities | ○ | ○ | ○ | ○ | ○ |
| Brain fog | ○ | ○ | ○ | ○ | ○ |
| Brain pressure | ○ | ○ | ○ | ○ | ○ |
| Bulging veins | ○ | ○ | ○ | ○ | ○ |
| Burning sensations | ○ | ○ | ○ | ○ | ○ |
| Bruising of skin | ○ | ○ | ○ | ○ | ○ |
| Calf cramps | ○ | ○ | ○ | ○ | ○ |
| Change in nails (i.e. white spots, brittleness, change in moons) | ○ | ○ | ○ | ○ | ○ |
| Changes in voice | ○ | ○ | ○ | ○ | ○ |
| Changed sense of taste | ○ | ○ | ○ | ○ | ○ |
| Chills but no fever | ○ | ○ | ○ | ○ | ○ |
| Clogged ears | ○ | ○ | ○ | ○ | ○ |
| Cold burning feeling in lungs | ○ | ○ | ○ | ○ | ○ |
| Confusion | ○ | ○ | ○ | ○ | ○ |
| Congested or runny nose | ○ | ○ | ○ | ○ | ○ |
| Constant thirst | ○ | ○ | ○ | ○ | ○ |
| Costochondritis (inflammation of the cartilage that connects a rib to the breastbone) | ○ | ○ | ○ | ○ | ○ |
| Cough | ○ | ○ | ○ | ○ | ○ |

| | | | | | |
|---|---|---|---|---|---|
| Coughing up blood | ○ | ○ | ○ | ○ | ○ |
| Covid toes (tender or itchy rash or chilblains on the toes or foot) | ○ | ○ | ○ | ○ | ○ |
| Cracked or dry lips | ○ | ○ | ○ | ○ | ○ |
| Dental problems (i.e. chipped tooth, tooth loss) | ○ | ○ | ○ | ○ | ○ |
| Diarrhea | ○ | ○ | ○ | ○ | ○ |
| Difficulty concentrating or focusing | ○ | ○ | ○ | ○ | ○ |
| Difficulty sleeping | ○ | ○ | ○ | ○ | ○ |
| Difficulty speaking properly | ○ | ○ | ○ | ○ | ○ |
| Discoloration of the skin (for example: purple or blue on the hands or feet, no blistering) | ○ | ○ | ○ | ○ | ○ |
| Dizziness | ○ | ○ | ○ | ○ | ○ |
| Dry eyes | ○ | ○ | ○ | ○ | ○ |
| Dry or peeling skin | ○ | ○ | ○ | ○ | ○ |
| Dry scalp or dandruff | ○ | ○ | ○ | ○ | ○ |
| Dry throat | ○ | ○ | ○ | ○ | ○ |
| Ear pain/earache | ○ | ○ | ○ | ○ | ○ |
| Elevated thyroid hormones | ○ | ○ | ○ | ○ | ○ |
| Extreme pressure at base of head or occipital nerve | ○ | ○ | ○ | ○ | ○ |
| Eye stye or infection | ○ | ○ | ○ | ○ | ○ |
| Fatigue | ○ | ○ | ○ | ○ | ○ |
| Feeling irritable | ○ | ○ | ○ | ○ | ○ |
| Feeling of burning skin | ○ | ○ | ○ | ○ | ○ |
| Fever or chills | ○ | ○ | ○ | ○ | ○ |
| Floaters or flashes of light in vision | ○ | ○ | ○ | ○ | ○ |
| Foot pain | ○ | ○ | ○ | ○ | ○ |
| GERD (acid reflux) with excessive salivation | ○ | ○ | ○ | ○ | ○ |
| Goiter or lump in throat | ○ | ○ | ○ | ○ | ○ |
| Hair loss | ○ | ○ | ○ | ○ | ○ |
| Hand or wrist pain | ○ | ○ | ○ | ○ | ○ |
| Headache | ○ | ○ | ○ | ○ | ○ |
| Heart palpitations (heart skipping a beat or racing) | ○ | ○ | ○ | ○ | ○ |
| Heat intolerance | ○ | ○ | ○ | ○ | ○ |
| High blood pressure | ○ | ○ | ○ | ○ | ○ |

| | | | | | |
|---|---|---|---|---|---|
| Hormone imbalances | ○ | ○ | ○ | ○ | ○ |
| "Hot" blood rush | ○ | ○ | ○ | ○ | ○ |
| Inability to cry | ○ | ○ | ○ | ○ | ○ |
| Inability to exercise or be active | ○ | ○ | ○ | ○ | ○ |
| Inability to yawn | ○ | ○ | ○ | ○ | ○ |
| Internal tremors or buzzing/vibration | ○ | ○ | ○ | ○ | ○ |
| Irregular or skipped menstrual cycles | ○ | ○ | ○ | ○ | ○ |
| Jaw pain | ○ | ○ | ○ | ○ | ○ |
| Joint pain | ○ | ○ | ○ | ○ | ○ |
| Kidney issues or protein in urine | ○ | ○ | ○ | ○ | ○ |
| Kidney pain | ○ | ○ | ○ | ○ | ○ |
| Low blood oxygen | ○ | ○ | ○ | ○ | ○ |
| Low blood pressure | ○ | ○ | ○ | ○ | ○ |
| Lower back pain | ○ | ○ | ○ | ○ | ○ |
| Loss of appetite | ○ | ○ | ○ | ○ | ○ |
| Loss of hearing | ○ | ○ | ○ | ○ | ○ |
| Loss or decrease in quality of vision | ○ | ○ | ○ | ○ | ○ |
| Lump in throat/difficulty swallowing | ○ | ○ | ○ | ○ | ○ |
| Memory problems | ○ | ○ | ○ | ○ | ○ |
| Menstrual cycles that are heavier or lighter than normal | ○ | ○ | ○ | ○ | ○ |
| Mid-back pain at base of ribs | ○ | ○ | ○ | ○ | ○ |
| Mouth sores or sore tongue | ○ | ○ | ○ | ○ | ○ |
| Muscle or body aches | ○ | ○ | ○ | ○ | ○ |
| Muscle twitching | ○ | ○ | ○ | ○ | ○ |
| Nausea or vomiting | ○ | ○ | ○ | ○ | ○ |
| Neck muscle pain | ○ | ○ | ○ | ○ | ○ |
| Nerve pain | ○ | ○ | ○ | ○ | ○ |
| Nerve sensations (tingling, pins and needles, numbness) | ○ | ○ | ○ | ○ | ○ |
| Neuropathy in feet and hands (weakness, numbness, and pain) | ○ | ○ | ○ | ○ | ○ |
| New allergies | ○ | ○ | ○ | ○ | ○ |
| Night sweats | ○ | ○ | ○ | ○ | ○ |
| Nightmares | ○ | ○ | ○ | ○ | ○ |
| Painful scalp | ○ | ○ | ○ | ○ | ○ |

| | | | | | |
|---|---|---|---|---|---|
| Partial or complete loss of sense of smell | ○ | ○ | ○ | ○ | ○ |
| Partial or complete loss of sense of taste | ○ | ○ | ○ | ○ | ○ |
| Persistent chest pain or pressure | ○ | ○ | ○ | ○ | ○ |
| Personality change (drastic) | ○ | ○ | ○ | ○ | ○ |
| Petechiae (pinpoint rash) | ○ | ○ | ○ | ○ | ○ |
| Phantom smells | ○ | ○ | ○ | ○ | ○ |
| Phlegm in back of throat | ○ | ○ | ○ | ○ | ○ |
| Post-exertional malaise (worsened symptoms or flu-like symptoms after exertion) | ○ | ○ | ○ | ○ | ○ |
| Postnasal drip | ○ | ○ | ○ | ○ | ○ |
| Rash | ○ | ○ | ○ | ○ | ○ |
| Reflux or heartburn | ○ | ○ | ○ | ○ | ○ |
| Runny nose | ○ | ○ | ○ | ○ | ○ |
| Sadness | ○ | ○ | ○ | ○ | ○ |
| Seizures | ○ | ○ | ○ | ○ | ○ |
| Sharp or sudden chest pain | ○ | ○ | ○ | ○ | ○ |
| Shortness of breath or difficulty breathing | ○ | ○ | ○ | ○ | ○ |
| Shortness of breath or exhaustion from bending over | ○ | ○ | ○ | ○ | ○ |
| Sleeping more than normal | ○ | ○ | ○ | ○ | ○ |
| Sore throat | ○ | ○ | ○ | ○ | ○ |
| Spinal issues | ○ | ○ | ○ | ○ | ○ |
| Spikes in blood pressure | ○ | ○ | ○ | ○ | ○ |
| Swollen hands or feet | ○ | ○ | ○ | ○ | ○ |
| Swollen lymph nodes | ○ | ○ | ○ | ○ | ○ |
| Syncope (fainting) | ○ | ○ | ○ | ○ | ○ |
| Tachycardia (rapid heartbeat) at rest | ○ | ○ | ○ | ○ | ○ |
| Tachycardia (rapid heartbeat) after standing up | ○ | ○ | ○ | ○ | ○ |
| Thrush (white fungal infection in the mouth or throat) | ○ | ○ | ○ | ○ | ○ |
| Tinnitus or humming in ears | ○ | ○ | ○ | ○ | ○ |
| Tremors or shakiness | ○ | ○ | ○ | ○ | ○ |
| Upper back pain | ○ | ○ | ○ | ○ | ○ |
| UTI (urinary tract infection) | ○ | ○ | ○ | ○ | ○ |
| Weakened neck | ○ | ○ | ○ | ○ | ○ |

| | | | | | |
|---|---|---|---|---|---|
| Weight gain | ◯ | ◯ | ◯ | ◯ | ◯ |

**Since [vaccine_arm_1][v_v_vax_date], while experiencing the symptom, how much did it impair your social or family functioning compared to pre-COVID?**

***If you have not experienced these symptoms since your vaccine, please choose "Not at all"**

| | Not at all | A little bit | Somewhat | Quite a bit | Very much |
|---|---|---|---|---|---|
| Abdominal pain | ○ | ○ | ○ | ○ | ○ |
| Abnormally low temperature | ○ | ○ | ○ | ○ | ○ |
| Acid reflux | ○ | ○ | ○ | ○ | ○ |
| Afternoon or evening fevers/low-grade fevers | ○ | ○ | ○ | ○ | ○ |
| Anemia (low number of red blood cells) | ○ | ○ | ○ | ○ | ○ |
| Anxiety | ○ | ○ | ○ | ○ | ○ |
| Arrythmia (improper beating of the heart due to electrical impulse problems) | ○ | ○ | ○ | ○ | ○ |
| Bilateral neck throbbing around lymph nodes | ○ | ○ | ○ | ○ | ○ |
| Blurry vision | ○ | ○ | ○ | ○ | ○ |
| Bone aches in extremities | ○ | ○ | ○ | ○ | ○ |
| Brain fog | ○ | ○ | ○ | ○ | ○ |
| Brain pressure | ○ | ○ | ○ | ○ | ○ |
| Bulging veins | ○ | ○ | ○ | ○ | ○ |
| Burning sensations | ○ | ○ | ○ | ○ | ○ |
| Bruising of skin | ○ | ○ | ○ | ○ | ○ |
| Calf cramps | ○ | ○ | ○ | ○ | ○ |
| Change in nails (i.e. white spots, brittleness, change in moons) | ○ | ○ | ○ | ○ | ○ |
| Changes in voice | ○ | ○ | ○ | ○ | ○ |
| Changed sense of taste | ○ | ○ | ○ | ○ | ○ |
| Chills but no fever | ○ | ○ | ○ | ○ | ○ |
| Clogged ears | ○ | ○ | ○ | ○ | ○ |
| Cold burning feeling in lungs | ○ | ○ | ○ | ○ | ○ |
| Confusion | ○ | ○ | ○ | ○ | ○ |
| Congested or runny nose | ○ | ○ | ○ | ○ | ○ |
| Constant thirst | ○ | ○ | ○ | ○ | ○ |
| Costochondritis (inflammation of the cartilage that connects a rib to the breastbone) | ○ | ○ | ○ | ○ | ○ |
| Cough | ○ | ○ | ○ | ○ | ○ |

| | | | | | |
|---|---|---|---|---|---|
| Coughing up blood | ○ | ○ | ○ | ○ | ○ |
| Covid toes (tender or itchy rash or chilblains on the toes or foot) | ○ | ○ | ○ | ○ | ○ |
| Cracked or dry lips | ○ | ○ | ○ | ○ | ○ |
| Dental problems (i.e. chipped tooth, tooth loss) | ○ | ○ | ○ | ○ | ○ |
| Diarrhea | ○ | ○ | ○ | ○ | ○ |
| Difficulty concentrating or focusing | ○ | ○ | ○ | ○ | ○ |
| Difficulty sleeping | ○ | ○ | ○ | ○ | ○ |
| Difficulty speaking properly | ○ | ○ | ○ | ○ | ○ |
| Discoloration of the skin (for example: purple or blue on the hands or feet, no blistering) | ○ | ○ | ○ | ○ | ○ |
| Dizziness | ○ | ○ | ○ | ○ | ○ |
| Dry eyes | ○ | ○ | ○ | ○ | ○ |
| Dry or peeling skin | ○ | ○ | ○ | ○ | ○ |
| Dry scalp or dandruff | ○ | ○ | ○ | ○ | ○ |
| Dry throat | ○ | ○ | ○ | ○ | ○ |
| Ear pain/earache | ○ | ○ | ○ | ○ | ○ |
| Elevated thyroid hormones | ○ | ○ | ○ | ○ | ○ |
| Extreme pressure at base of head or occipital nerve | ○ | ○ | ○ | ○ | ○ |
| Eye stye or infection | ○ | ○ | ○ | ○ | ○ |
| Fatigue | ○ | ○ | ○ | ○ | ○ |
| Feeling irritable | ○ | ○ | ○ | ○ | ○ |
| Feeling of burning skin | ○ | ○ | ○ | ○ | ○ |
| Fever or chills | ○ | ○ | ○ | ○ | ○ |
| Floaters or flashes of light in vision | ○ | ○ | ○ | ○ | ○ |
| Foot pain | ○ | ○ | ○ | ○ | ○ |
| GERD (acid reflux) with excessive salivation | ○ | ○ | ○ | ○ | ○ |
| Goiter or lump in throat | ○ | ○ | ○ | ○ | ○ |
| Hair loss | ○ | ○ | ○ | ○ | ○ |
| Hand or wrist pain | ○ | ○ | ○ | ○ | ○ |
| Headache | ○ | ○ | ○ | ○ | ○ |
| Heart palpitations (heart skipping a beat or racing) | ○ | ○ | ○ | ○ | ○ |
| Heat intolerance | ○ | ○ | ○ | ○ | ○ |
| High blood pressure | ○ | ○ | ○ | ○ | ○ |

| | | | | | |
|---|---|---|---|---|---|
| Hormone imbalances | ○ | ○ | ○ | ○ | ○ |
| "Hot" blood rush | ○ | ○ | ○ | ○ | ○ |
| Inability to cry | ○ | ○ | ○ | ○ | ○ |
| Inability to exercise or be active | ○ | ○ | ○ | ○ | ○ |
| Inability to yawn | ○ | ○ | ○ | ○ | ○ |
| Internal tremors or buzzing/vibration | ○ | ○ | ○ | ○ | ○ |
| Irregular or skipped menstrual cycles | ○ | ○ | ○ | ○ | ○ |
| Jaw pain | ○ | ○ | ○ | ○ | ○ |
| Joint pain | ○ | ○ | ○ | ○ | ○ |
| Kidney issues or protein in urine | ○ | ○ | ○ | ○ | ○ |
| Kidney pain | ○ | ○ | ○ | ○ | ○ |
| Low blood oxygen | ○ | ○ | ○ | ○ | ○ |
| Low blood pressure | ○ | ○ | ○ | ○ | ○ |
| Lower back pain | ○ | ○ | ○ | ○ | ○ |
| Loss of appetite | ○ | ○ | ○ | ○ | ○ |
| Loss of hearing | ○ | ○ | ○ | ○ | ○ |
| Loss or decrease in quality of vision | ○ | ○ | ○ | ○ | ○ |
| Lump in throat/difficulty swallowing | ○ | ○ | ○ | ○ | ○ |
| Memory problems | ○ | ○ | ○ | ○ | ○ |
| Menstrual cycles that are heavier or lighter than normal | ○ | ○ | ○ | ○ | ○ |
| Mid-back pain at base of ribs | ○ | ○ | ○ | ○ | ○ |
| Mouth sores or sore tongue | ○ | ○ | ○ | ○ | ○ |
| Muscle or body aches | ○ | ○ | ○ | ○ | ○ |
| Muscle twitching | ○ | ○ | ○ | ○ | ○ |
| Nausea or vomiting | ○ | ○ | ○ | ○ | ○ |
| Neck muscle pain | ○ | ○ | ○ | ○ | ○ |
| Nerve pain | ○ | ○ | ○ | ○ | ○ |
| Nerve sensations (tingling, pins and needles, numbness) | ○ | ○ | ○ | ○ | ○ |
| Neuropathy in feet and hands (weakness, numbness, and pain) | ○ | ○ | ○ | ○ | ○ |
| New allergies | ○ | ○ | ○ | ○ | ○ |
| Night sweats | ○ | ○ | ○ | ○ | ○ |
| Nightmares | ○ | ○ | ○ | ○ | ○ |
| Painful scalp | ○ | ○ | ○ | ○ | ○ |

| | | | | | |
|---|---|---|---|---|---|
| Partial or complete loss of sense of smell | ○ | ○ | ○ | ○ | ○ |
| Partial or complete loss of sense of taste | ○ | ○ | ○ | ○ | ○ |
| Persistent chest pain or pressure | ○ | ○ | ○ | ○ | ○ |
| Personality change (drastic) | ○ | ○ | ○ | ○ | ○ |
| Petechiae (pinpoint rash) | ○ | ○ | ○ | ○ | ○ |
| Phantom smells | ○ | ○ | ○ | ○ | ○ |
| Phlegm in back of throat | ○ | ○ | ○ | ○ | ○ |
| Post-exertional malaise (worsened symptoms or flu-like symptoms after exertion) | ○ | ○ | ○ | ○ | ○ |
| Postnasal drip | ○ | ○ | ○ | ○ | ○ |
| Rash | ○ | ○ | ○ | ○ | ○ |
| Reflux or heartburn | ○ | ○ | ○ | ○ | ○ |
| Runny nose | ○ | ○ | ○ | ○ | ○ |
| Sadness | ○ | ○ | ○ | ○ | ○ |
| Seizures | ○ | ○ | ○ | ○ | ○ |
| Sharp or sudden chest pain | ○ | ○ | ○ | ○ | ○ |
| Shortness of breath or difficulty breathing | ○ | ○ | ○ | ○ | ○ |
| Shortness of breath or exhaustion from bending over | ○ | ○ | ○ | ○ | ○ |
| Sleeping more than normal | ○ | ○ | ○ | ○ | ○ |
| Sore throat | ○ | ○ | ○ | ○ | ○ |
| Spinal issues | ○ | ○ | ○ | ○ | ○ |
| Spikes in blood pressure | ○ | ○ | ○ | ○ | ○ |
| Swollen hands or feet | ○ | ○ | ○ | ○ | ○ |
| Swollen lymph nodes | ○ | ○ | ○ | ○ | ○ |
| Syncope (fainting) | ○ | ○ | ○ | ○ | ○ |
| Tachycardia (rapid heartbeat) at rest | ○ | ○ | ○ | ○ | ○ |
| Tachycardia (rapid heartbeat) after standing up | ○ | ○ | ○ | ○ | ○ |
| Thrush (white fungal infection in the mouth or throat) | ○ | ○ | ○ | ○ | ○ |
| Tinnitus or humming in ears | ○ | ○ | ○ | ○ | ○ |
| Tremors or shakiness | ○ | ○ | ○ | ○ | ○ |
| Upper back pain | ○ | ○ | ○ | ○ | ○ |
| UTI (urinary tract infection) | ○ | ○ | ○ | ○ | ○ |
| Weakened neck | ○ | ○ | ○ | ○ | ○ |

| | | | | | |
|---|---|---|---|---|---|
| Weight gain | ○ | ○ | ○ | ○ | ○ |

Are there any symptoms not listed above that you have experienced since you received the first dose of your vaccine on [vaccine_arm_1][v_v_vax_date]? ______________________________

*Please list all if so

**Since you received the first dose of your vaccine on [vaccine_arm_1][v_v_vax_date]**

| | 76-100% of health before COVID-19 | 51-75% of health before COVID-19 | 26-50% of health before COVID-19 | 0-25% of health before COVID-19 |
|---|---|---|---|---|
| On your best days, would you say that you are at | ◯ | ◯ | ◯ | ◯ |
| On your worst days, would you say that you are at | ◯ | ◯ | ◯ | ◯ |

Since you received the first dose of your vaccine on [vaccine_arm_1][v_v_vax_date], would you say that your health is

◯ Better
◯ Worse
◯ The same
◯ Don't know

Please, in the space below, share any information about your experience with COVID-19 or the vaccine that we might have missed.

__________________________________

### Survey 3

Thank you for taking Survey 3 for the Yale COVID Recovery Study!

Please remember that you may stop this survey and come back to it at any point. To return to the survey, please return to the link in your browser (or reclick the link in your email).

You may also find it helpful to have the below items ready as you complete the survey. If you do not have these items, please still fill in the information as best you can.

Testing results: dates, type (PCR, antigen) and result of tests. If you do not remember the exact date, the estimated date is enough.Symptom time and severity: your symptom log. You will be asked to pick symptoms you have had since you began having COVID-19 symptoms.

| | |
|---|---|
| Have you received your second dose of the COVID-19 vaccine? | ◯ Yes ______<br>◯ No, but it's scheduled<br>◯ No, and I don't have an appointment scheduled |
| When did you receive your second dose of the COVID-19 vaccine? | ______________________________ |

On your previous survey, you responded that you have experienced the following symptoms due to COVID-19 at some point in the past.

The following questions will ask about how these symptoms have changed, if at all. Please respond based on your experiences during the past month.

**During the past month (as compared to before one month ago), this symptom has been**

| | Better | Worse | The same |
|---|---|---|---|
| Abdominal pain | ◯ | ◯ | ◯ |
| Abnormally low temperature | ◯ | ◯ | ◯ |
| Acid reflux | ◯ | ◯ | ◯ |
| Afternoon or evening fevers/low-grade fevers | ◯ | ◯ | ◯ |
| Anemia (low number of red blood cells) | ◯ | ◯ | ◯ |
| Anxiety | ◯ | ◯ | ◯ |
| Arrythmia (improper beating of the heart due to electrical impulse problems) | ◯ | ◯ | ◯ |
| Bilateral neck throbbing around lymph nodes | ◯ | ◯ | ◯ |
| Blurry vision | ◯ | ◯ | ◯ |
| Bone aches in extremities | ◯ | ◯ | ◯ |
| Brain fog | ◯ | ◯ | ◯ |
| Brain pressure | ◯ | ◯ | ◯ |
| Bulging veins | ◯ | ◯ | ◯ |
| Burning sensations | ◯ | ◯ | ◯ |
| Bruising of skin | ◯ | ◯ | ◯ |
| Calf cramps | ◯ | ◯ | ◯ |
| Change in nails (i.e. white spots, brittleness, change in moons) | ◯ | ◯ | ◯ |
| Changes in voice | ◯ | ◯ | ◯ |
| Changed sense of taste | ◯ | ◯ | ◯ |
| Chills but no fever | ◯ | ◯ | ◯ |
| Clogged ears | ◯ | ◯ | ◯ |
| Cold burning feeling in lungs | ◯ | ◯ | ◯ |
| Confusion | ◯ | ◯ | ◯ |
| Congested or runny nose | ◯ | ◯ | ◯ |
| Constant thirst | ◯ | ◯ | ◯ |
| Costochondritis (inflammation of the cartilage that connects a rib to the breastbone) | ◯ | ◯ | ◯ |
| Cough | ◯ | ◯ | ◯ |
| Coughing up blood | ◯ | ◯ | ◯ |
| Covid toes (tender or itchy rash or chilblains on the toes or foot) | ◯ | ◯ | ◯ |

| | | | |
|---|---|---|---|
| Cracked or dry lips | ○ | ○ | ○ |
| Dental problems (i.e. chipped tooth, tooth loss) | ○ | ○ | ○ |
| Diarrhea | ○ | ○ | ○ |
| Difficulty concentrating or focusing | ○ | ○ | ○ |
| Difficulty sleeping | ○ | ○ | ○ |
| Difficulty speaking properly | ○ | ○ | ○ |
| Discoloration of the skin (for example: purple or blue on the hands or feet, no blistering) | ○ | ○ | ○ |
| Dizziness | ○ | ○ | ○ |
| Dry eyes | ○ | ○ | ○ |
| Dry or peeling skin | ○ | ○ | ○ |
| Dry scalp or dandruff | ○ | ○ | ○ |
| Dry throat | ○ | ○ | ○ |
| Ear pain/earache | ○ | ○ | ○ |
| Elevated thyroid hormones | ○ | ○ | ○ |
| Extreme pressure at base of head or occipital nerve | ○ | ○ | ○ |
| Eye stye or infection | ○ | ○ | ○ |
| Fatigue | ○ | ○ | ○ |
| Feeling irritable | ○ | ○ | ○ |
| Feeling of burning skin | ○ | ○ | ○ |
| Fever or chills | ○ | ○ | ○ |
| Floaters or flashes of light in vision | ○ | ○ | ○ |
| Foot pain | ○ | ○ | ○ |
| GERD (acid reflux) with excessive salivation | ○ | ○ | ○ |
| Goiter or lump in throat | ○ | ○ | ○ |
| Hair loss | ○ | ○ | ○ |
| Hand or wrist pain | ○ | ○ | ○ |
| Headache | ○ | ○ | ○ |
| Heart palpitations (heart skipping a beat or racing) | ○ | ○ | ○ |
| Heat intolerance | ○ | ○ | ○ |
| High blood pressure | ○ | ○ | ○ |
| Hormone imbalances | ○ | ○ | ○ |
| "Hot" blood rush | ○ | ○ | ○ |
| Inability to cry | ○ | ○ | ○ |

| | | | |
|---|---|---|---|
| Inability to exercise or be active | ○ | ○ | ○ |
| Inability to yawn | ○ | ○ | ○ |
| Internal tremors or buzzing/vibration | ○ | ○ | ○ |
| Irregular or skipped menstrual cycles | ○ | ○ | ○ |
| Jaw pain | ○ | ○ | ○ |
| Joint pain | ○ | ○ | ○ |
| Kidney issues or protein in urine | ○ | ○ | ○ |
| Kidney pain | ○ | ○ | ○ |
| Low blood oxygen | ○ | ○ | ○ |
| Low blood pressure | ○ | ○ | ○ |
| Lower back pain | ○ | ○ | ○ |
| Loss of appetite | ○ | ○ | ○ |
| Loss of hearing | ○ | ○ | ○ |
| Loss or decrease in quality of vision | ○ | ○ | ○ |
| Lump in throat/difficulty swallowing | ○ | ○ | ○ |
| Memory problems | ○ | ○ | ○ |
| Menstrual cycles that are heavier or lighter than normal | ○ | ○ | ○ |
| Mid-back pain at base of ribs | ○ | ○ | ○ |
| Mouth sores or sore tongue | ○ | ○ | ○ |
| Muscle or body aches | ○ | ○ | ○ |
| Muscle twitching | ○ | ○ | ○ |
| Nausea or vomiting | ○ | ○ | ○ |
| Neck muscle pain | ○ | ○ | ○ |
| Nerve pain | ○ | ○ | ○ |
| Nerve sensations (tingling, pins and needles, numbness) | ○ | ○ | ○ |
| Neuropathy in feet and hands (weakness, numbness, and pain) | ○ | ○ | ○ |
| New allergies | ○ | ○ | ○ |
| Night sweats | ○ | ○ | ○ |
| Nightmares | ○ | ○ | ○ |
| Painful scalp | ○ | ○ | ○ |
| Partial or complete loss of sense of smell | ○ | ○ | ○ |
| Partial or complete loss of sense of taste | ○ | ○ | ○ |

| | | | |
|---|---|---|---|
| Persistent chest pain or pressure | ○ | ○ | ○ |
| Personality change (drastic) | ○ | ○ | ○ |
| Petechiae (pinpoint rash) | ○ | ○ | ○ |
| Phantom smells | ○ | ○ | ○ |
| Phlegm in back of throat | ○ | ○ | ○ |
| Post-exertional malaise (worsened symptoms or flu-like symptoms after exertion) | ○ | ○ | ○ |
| Postnasal drip | ○ | ○ | ○ |
| Rash | ○ | ○ | ○ |
| Reflux or heartburn | ○ | ○ | ○ |
| Runny nose | ○ | ○ | ○ |
| Sadness | ○ | ○ | ○ |
| Seizures | ○ | ○ | ○ |
| Sharp or sudden chest pain | ○ | ○ | ○ |
| Shortness of breath or difficulty breathing | ○ | ○ | ○ |
| Shortness of breath or exhaustion from bending over | ○ | ○ | ○ |
| Sleeping more than normal | ○ | ○ | ○ |
| Sore throat | ○ | ○ | ○ |
| Spinal issues | ○ | ○ | ○ |
| Spikes in blood pressure | ○ | ○ | ○ |
| Swollen hands or feet | ○ | ○ | ○ |
| Swollen lymph nodes | ○ | ○ | ○ |
| Syncope (fainting) | ○ | ○ | ○ |
| Tachycardia (rapid heartbeat) at rest | ○ | ○ | ○ |
| Tachycardia (rapid heartbeat) after standing up | ○ | ○ | ○ |
| Thrush (white fungal infection in the mouth or throat) | ○ | ○ | ○ |
| Tinnitus or humming in ears | ○ | ○ | ○ |
| Tremors or shakiness | ○ | ○ | ○ |
| Upper back pain | ○ | ○ | ○ |
| UTI (urinary tract infection) | ○ | ○ | ○ |
| Weakened neck | ○ | ○ | ○ |
| Weight gain | ○ | ○ | ○ |

#### During the past month, I have experienced the symptom, on average

| | Never--it has gone away completely | Once a month | Once or twice a week | 3 or 4 times a week | Most days | All the time |
|---|---|---|---|---|---|---|
| Abdominal pain | ◯ | ◯ | ◯ | ◯ | ◯ | ◯ |
| Abnormally low temperature | ◯ | ◯ | ◯ | ◯ | ◯ | ◯ |
| Acid reflux | ◯ | ◯ | ◯ | ◯ | ◯ | ◯ |
| Afternoon or evening fevers/low-grade fevers | ◯ | ◯ | ◯ | ◯ | ◯ | ◯ |
| Anemia (low number of red blood cells) | ◯ | ◯ | ◯ | ◯ | ◯ | ◯ |
| Anxiety | ◯ | ◯ | ◯ | ◯ | ◯ | ◯ |
| Arrythmia (improper beating of the heart due to electrical impulse problems) | ◯ | ◯ | ◯ | ◯ | ◯ | ◯ |
| Bilateral neck throbbing around lymph nodes | ◯ | ◯ | ◯ | ◯ | ◯ | ◯ |
| Blurry vision | ◯ | ◯ | ◯ | ◯ | ◯ | ◯ |
| Bone aches in extremities | ◯ | ◯ | ◯ | ◯ | ◯ | ◯ |
| Brain fog | ◯ | ◯ | ◯ | ◯ | ◯ | ◯ |
| Brain pressure | ◯ | ◯ | ◯ | ◯ | ◯ | ◯ |
| Bulging veins | ◯ | ◯ | ◯ | ◯ | ◯ | ◯ |
| Burning sensations | ◯ | ◯ | ◯ | ◯ | ◯ | ◯ |
| Bruising of skin | ◯ | ◯ | ◯ | ◯ | ◯ | ◯ |
| Calf cramps | ◯ | ◯ | ◯ | ◯ | ◯ | ◯ |
| Change in nails (i.e. white spots, brittleness, change in moons) | ◯ | ◯ | ◯ | ◯ | ◯ | ◯ |
| Changes in voice | ◯ | ◯ | ◯ | ◯ | ◯ | ◯ |
| Changed sense of taste | ◯ | ◯ | ◯ | ◯ | ◯ | ◯ |
| Chills but no fever | ◯ | ◯ | ◯ | ◯ | ◯ | ◯ |
| Clogged ears | ◯ | ◯ | ◯ | ◯ | ◯ | ◯ |
| Cold burning feeling in lungs | ◯ | ◯ | ◯ | ◯ | ◯ | ◯ |
| Confusion | ◯ | ◯ | ◯ | ◯ | ◯ | ◯ |
| Congested or runny nose | ◯ | ◯ | ◯ | ◯ | ◯ | ◯ |
| Constant thirst | ◯ | ◯ | ◯ | ◯ | ◯ | ◯ |
| Costochondritis (inflammation of the cartilage that connects a rib to the breastbone) | ◯ | ◯ | ◯ | ◯ | ◯ | ◯ |
| Cough | ◯ | ◯ | ◯ | ◯ | ◯ | ◯ |
| Coughing up blood | ◯ | ◯ | ◯ | ◯ | ◯ | ◯ |

| | | | | | | |
|---|---|---|---|---|---|---|
| Covid toes (tender or itchy rash or chilblains on the toes or foot) | ○ | ○ | ○ | ○ | ○ | ○ |
| Cracked or dry lips | ○ | ○ | ○ | ○ | ○ | ○ |
| Dental problems (i.e. chipped tooth, tooth loss) | ○ | ○ | ○ | ○ | ○ | ○ |
| Diarrhea | ○ | ○ | ○ | ○ | ○ | ○ |
| Difficulty concentrating or focusing | ○ | ○ | ○ | ○ | ○ | ○ |
| Difficulty sleeping | ○ | ○ | ○ | ○ | ○ | ○ |
| Difficulty speaking properly | ○ | ○ | ○ | ○ | ○ | ○ |
| Discoloration of the skin (for example: purple or blue on the hands or feet, no blistering) | ○ | ○ | ○ | ○ | ○ | ○ |
| Dizziness | ○ | ○ | ○ | ○ | ○ | ○ |
| Dry eyes | ○ | ○ | ○ | ○ | ○ | ○ |
| Dry or peeling skin | ○ | ○ | ○ | ○ | ○ | ○ |
| Dry scalp or dandruff | ○ | ○ | ○ | ○ | ○ | ○ |
| Dry throat | ○ | ○ | ○ | ○ | ○ | ○ |
| Ear pain/earache | ○ | ○ | ○ | ○ | ○ | ○ |
| Elevated thyroid hormones | ○ | ○ | ○ | ○ | ○ | ○ |
| Extreme pressure at base of head or occipital nerve | ○ | ○ | ○ | ○ | ○ | ○ |
| Eye stye or infection | ○ | ○ | ○ | ○ | ○ | ○ |
| Fatigue | ○ | ○ | ○ | ○ | ○ | ○ |
| Feeling irritable | ○ | ○ | ○ | ○ | ○ | ○ |
| Feeling of burning skin | ○ | ○ | ○ | ○ | ○ | ○ |
| Fever or chills | ○ | ○ | ○ | ○ | ○ | ○ |
| Floaters or flashes of light in vision | ○ | ○ | ○ | ○ | ○ | ○ |
| Foot pain | ○ | ○ | ○ | ○ | ○ | ○ |
| GERD (acid reflux) with excessive salivation | ○ | ○ | ○ | ○ | ○ | ○ |
| Goiter or lump in throat | ○ | ○ | ○ | ○ | ○ | ○ |
| Hair loss | ○ | ○ | ○ | ○ | ○ | ○ |
| Hand or wrist pain | ○ | ○ | ○ | ○ | ○ | ○ |
| Headache | ○ | ○ | ○ | ○ | ○ | ○ |
| Heart palpitations (heart skipping a beat or racing) | ○ | ○ | ○ | ○ | ○ | ○ |
| Heat intolerance | ○ | ○ | ○ | ○ | ○ | ○ |
| High blood pressure | ○ | ○ | ○ | ○ | ○ | ○ |
| Hormone imbalances | ○ | ○ | ○ | ○ | ○ | ○ |

| | | | | | | |
|---|---|---|---|---|---|---|
| "Hot" blood rush | ○ | ○ | ○ | ○ | ○ | ○ |
| Inability to cry | ○ | ○ | ○ | ○ | ○ | ○ |
| Inability to exercise or be active | ○ | ○ | ○ | ○ | ○ | ○ |
| Inability to yawn | ○ | ○ | ○ | ○ | ○ | ○ |
| Internal tremors or buzzing/vibration | ○ | ○ | ○ | ○ | ○ | ○ |
| Irregular or skipped menstrual cycles | ○ | ○ | ○ | ○ | ○ | ○ |
| Jaw pain | ○ | ○ | ○ | ○ | ○ | ○ |
| Joint pain | ○ | ○ | ○ | ○ | ○ | ○ |
| Kidney issues or protein in urine | ○ | ○ | ○ | ○ | ○ | ○ |
| Kidney pain | ○ | ○ | ○ | ○ | ○ | ○ |
| Low blood oxygen | ○ | ○ | ○ | ○ | ○ | ○ |
| Low blood pressure | ○ | ○ | ○ | ○ | ○ | ○ |
| Lower back pain | ○ | ○ | ○ | ○ | ○ | ○ |
| Loss of appetite | ○ | ○ | ○ | ○ | ○ | ○ |
| Loss of hearing | ○ | ○ | ○ | ○ | ○ | ○ |
| Loss or decrease in quality of vision | ○ | ○ | ○ | ○ | ○ | ○ |
| Lump in throat/difficulty swallowing | ○ | ○ | ○ | ○ | ○ | ○ |
| Memory problems | ○ | ○ | ○ | ○ | ○ | ○ |
| Menstrual cycles that are heavier or lighter than normal | ○ | ○ | ○ | ○ | ○ | ○ |
| Mid-back pain at base of ribs | ○ | ○ | ○ | ○ | ○ | ○ |
| Mouth sores or sore tongue | ○ | ○ | ○ | ○ | ○ | ○ |
| Muscle or body aches | ○ | ○ | ○ | ○ | ○ | ○ |
| Muscle twitching | ○ | ○ | ○ | ○ | ○ | ○ |
| Nausea or vomiting | ○ | ○ | ○ | ○ | ○ | ○ |
| Neck muscle pain | ○ | ○ | ○ | ○ | ○ | ○ |
| Nerve pain | ○ | ○ | ○ | ○ | ○ | ○ |
| Nerve sensations (tingling, pins and needles, numbness) | ○ | ○ | ○ | ○ | ○ | ○ |
| Neuropathy in feet and hands (weakness, numbness, and pain) | ○ | ○ | ○ | ○ | ○ | ○ |
| New allergies | ○ | ○ | ○ | ○ | ○ | ○ |
| Night sweats | ○ | ○ | ○ | ○ | ○ | ○ |
| Nightmares | ○ | ○ | ○ | ○ | ○ | ○ |
| Painful scalp | ○ | ○ | ○ | ○ | ○ | ○ |
| Partial or complete loss of sense of smell | ○ | ○ | ○ | ○ | ○ | ○ |

| | | | | | | |
|---|---|---|---|---|---|---|
| Partial or complete loss of sense of taste | ○ | ○ | ○ | ○ | ○ | ○ |
| Persistent chest pain or pressure | ○ | ○ | ○ | ○ | ○ | ○ |
| Personality change (drastic) | ○ | ○ | ○ | ○ | ○ | ○ |
| Petechiae (pinpoint rash) | ○ | ○ | ○ | ○ | ○ | ○ |
| Phantom smells | ○ | ○ | ○ | ○ | ○ | ○ |
| Phlegm in back of throat | ○ | ○ | ○ | ○ | ○ | ○ |
| Post-exertional malaise (worsened symptoms or flu-like symptoms after exertion) | ○ | ○ | ○ | ○ | ○ | ○ |
| Postnasal drip | ○ | ○ | ○ | ○ | ○ | ○ |
| Rash | ○ | ○ | ○ | ○ | ○ | ○ |
| Reflux or heartburn | ○ | ○ | ○ | ○ | ○ | ○ |
| Runny nose | ○ | ○ | ○ | ○ | ○ | ○ |
| Sadness | ○ | ○ | ○ | ○ | ○ | ○ |
| Seizures | ○ | ○ | ○ | ○ | ○ | ○ |
| Sharp or sudden chest pain | ○ | ○ | ○ | ○ | ○ | ○ |
| Shortness of breath or difficulty breathing | ○ | ○ | ○ | ○ | ○ | ○ |
| Shortness of breath or exhaustion from bending over | ○ | ○ | ○ | ○ | ○ | ○ |
| Sleeping more than normal | ○ | ○ | ○ | ○ | ○ | ○ |
| Sore throat | ○ | ○ | ○ | ○ | ○ | ○ |
| Spinal issues | ○ | ○ | ○ | ○ | ○ | ○ |
| Spikes in blood pressure | ○ | ○ | ○ | ○ | ○ | ○ |
| Swollen hands or feet | ○ | ○ | ○ | ○ | ○ | ○ |
| Swollen lymph nodes | ○ | ○ | ○ | ○ | ○ | ○ |
| Syncope (fainting) | ○ | ○ | ○ | ○ | ○ | ○ |
| Tachycardia (rapid heartbeat) at rest | ○ | ○ | ○ | ○ | ○ | ○ |
| Tachycardia (rapid heartbeat) after standing up | ○ | ○ | ○ | ○ | ○ | ○ |
| Thrush (white fungal infection in the mouth or throat) | ○ | ○ | ○ | ○ | ○ | ○ |
| Tinnitus or humming in ears | ○ | ○ | ○ | ○ | ○ | ○ |
| Tremors or shakiness | ○ | ○ | ○ | ○ | ○ | ○ |
| Upper back pain | ○ | ○ | ○ | ○ | ○ | ○ |
| UTI (urinary tract infection) | ○ | ○ | ○ | ○ | ○ | ○ |
| Weakened neck | ○ | ○ | ○ | ○ | ○ | ○ |
| Weight gain | ○ | ○ | ○ | ○ | ○ | ○ |

---

The next set of questions will ask about the same symptoms. Please answer based on your experience during the past month. If you have not experienced any of the symptoms at all, please select "Not at all" from the choices.

**During the past month, while experiencing the symptom, how much did it bothered you in terms of discomfort or pain?**

**If you have not experienced the symptom during the past month, please choose "Not at all."**

| | Not at all | A little bit | Somewhat | Quite a bit | Very much |
|---|---|---|---|---|---|
| Abdominal pain | ◯ | ◯ | ◯ | ◯ | ◯ |
| Abnormally low temperature | ◯ | ◯ | ◯ | ◯ | ◯ |
| Acid reflux | ◯ | ◯ | ◯ | ◯ | ◯ |
| Afternoon or evening fevers/low-grade fevers | ◯ | ◯ | ◯ | ◯ | ◯ |
| Anemia (low number of red blood cells) | ◯ | ◯ | ◯ | ◯ | ◯ |
| Anxiety | ◯ | ◯ | ◯ | ◯ | ◯ |
| Arrythmia (improper beating of the heart due to electrical impulse problems) | ◯ | ◯ | ◯ | ◯ | ◯ |
| Bilateral neck throbbing around lymph nodes | ◯ | ◯ | ◯ | ◯ | ◯ |
| Blurry vision | ◯ | ◯ | ◯ | ◯ | ◯ |
| Bone aches in extremities | ◯ | ◯ | ◯ | ◯ | ◯ |
| Brain fog | ◯ | ◯ | ◯ | ◯ | ◯ |
| Brain pressure | ◯ | ◯ | ◯ | ◯ | ◯ |
| Bulging veins | ◯ | ◯ | ◯ | ◯ | ◯ |
| Burning sensations | ◯ | ◯ | ◯ | ◯ | ◯ |
| Bruising of skin | ◯ | ◯ | ◯ | ◯ | ◯ |
| Calf cramps | ◯ | ◯ | ◯ | ◯ | ◯ |
| Change in nails (i.e. white spots, brittleness, change in moons) | ◯ | ◯ | ◯ | ◯ | ◯ |
| Changes in voice | ◯ | ◯ | ◯ | ◯ | ◯ |
| Changed sense of taste | ◯ | ◯ | ◯ | ◯ | ◯ |
| Chills but no fever | ◯ | ◯ | ◯ | ◯ | ◯ |
| Clogged ears | ◯ | ◯ | ◯ | ◯ | ◯ |
| Cold burning feeling in lungs | ◯ | ◯ | ◯ | ◯ | ◯ |
| Confusion | ◯ | ◯ | ◯ | ◯ | ◯ |
| Congested or runny nose | ◯ | ◯ | ◯ | ◯ | ◯ |
| Constant thirst | ◯ | ◯ | ◯ | ◯ | ◯ |
| Costochondritis (inflammation of the cartilage that connects a rib to the breastbone) | ◯ | ◯ | ◯ | ◯ | ◯ |
| Cough | ◯ | ◯ | ◯ | ◯ | ◯ |

| | | | | | |
|---|---|---|---|---|---|
| Coughing up blood | ○ | ○ | ○ | ○ | ○ |
| Covid toes (tender or itchy rash or chilblains on the toes or foot) | ○ | ○ | ○ | ○ | ○ |
| Cracked or dry lips | ○ | ○ | ○ | ○ | ○ |
| Dental problems (i.e. chipped tooth, tooth loss) | ○ | ○ | ○ | ○ | ○ |
| Diarrhea | ○ | ○ | ○ | ○ | ○ |
| Difficulty concentrating or focusing | ○ | ○ | ○ | ○ | ○ |
| Difficulty sleeping | ○ | ○ | ○ | ○ | ○ |
| Difficulty speaking properly | ○ | ○ | ○ | ○ | ○ |
| Discoloration of the skin (for example: purple or blue on the hands or feet, no blistering) | ○ | ○ | ○ | ○ | ○ |
| Dizziness | ○ | ○ | ○ | ○ | ○ |
| Dry eyes | ○ | ○ | ○ | ○ | ○ |
| Dry or peeling skin | ○ | ○ | ○ | ○ | ○ |
| Dry scalp or dandruff | ○ | ○ | ○ | ○ | ○ |
| Dry throat | ○ | ○ | ○ | ○ | ○ |
| Ear pain/earache | ○ | ○ | ○ | ○ | ○ |
| Elevated thyroid hormones | ○ | ○ | ○ | ○ | ○ |
| Extreme pressure at base of head or occipital nerve | ○ | ○ | ○ | ○ | ○ |
| Eye stye or infection | ○ | ○ | ○ | ○ | ○ |
| Fatigue | ○ | ○ | ○ | ○ | ○ |
| Feeling irritable | ○ | ○ | ○ | ○ | ○ |
| Feeling of burning skin | ○ | ○ | ○ | ○ | ○ |
| Fever or chills | ○ | ○ | ○ | ○ | ○ |
| Floaters or flashes of light in vision | ○ | ○ | ○ | ○ | ○ |
| Foot pain | ○ | ○ | ○ | ○ | ○ |
| GERD (acid reflux) with excessive salivation | ○ | ○ | ○ | ○ | ○ |
| Goiter or lump in throat | ○ | ○ | ○ | ○ | ○ |
| Hair loss | ○ | ○ | ○ | ○ | ○ |
| Hand or wrist pain | ○ | ○ | ○ | ○ | ○ |
| Headache | ○ | ○ | ○ | ○ | ○ |
| Heart palpitations (heart skipping a beat or racing) | ○ | ○ | ○ | ○ | ○ |
| Heat intolerance | ○ | ○ | ○ | ○ | ○ |
| High blood pressure | ○ | ○ | ○ | ○ | ○ |

| | | | | | |
|---|---|---|---|---|---|
| Hormone imbalances | ○ | ○ | ○ | ○ | ○ |
| "Hot" blood rush | ○ | ○ | ○ | ○ | ○ |
| Inability to cry | ○ | ○ | ○ | ○ | ○ |
| Inability to exercise or be active | ○ | ○ | ○ | ○ | ○ |
| Inability to yawn | ○ | ○ | ○ | ○ | ○ |
| Internal tremors or buzzing/vibration | ○ | ○ | ○ | ○ | ○ |
| Irregular or skipped menstrual cycles | ○ | ○ | ○ | ○ | ○ |
| Jaw pain | ○ | ○ | ○ | ○ | ○ |
| Joint pain | ○ | ○ | ○ | ○ | ○ |
| Kidney issues or protein in urine | ○ | ○ | ○ | ○ | ○ |
| Kidney pain | ○ | ○ | ○ | ○ | ○ |
| Low blood oxygen | ○ | ○ | ○ | ○ | ○ |
| Low blood pressure | ○ | ○ | ○ | ○ | ○ |
| Lower back pain | ○ | ○ | ○ | ○ | ○ |
| Loss of appetite | ○ | ○ | ○ | ○ | ○ |
| Loss of hearing | ○ | ○ | ○ | ○ | ○ |
| Loss or decrease in quality of vision | ○ | ○ | ○ | ○ | ○ |
| Lump in throat/difficulty swallowing | ○ | ○ | ○ | ○ | ○ |
| Memory problems | ○ | ○ | ○ | ○ | ○ |
| Menstrual cycles that are heavier or lighter than normal | ○ | ○ | ○ | ○ | ○ |
| Mid-back pain at base of ribs | ○ | ○ | ○ | ○ | ○ |
| Mouth sores or sore tongue | ○ | ○ | ○ | ○ | ○ |
| Muscle or body aches | ○ | ○ | ○ | ○ | ○ |
| Muscle twitching | ○ | ○ | ○ | ○ | ○ |
| Nausea or vomiting | ○ | ○ | ○ | ○ | ○ |
| Neck muscle pain | ○ | ○ | ○ | ○ | ○ |
| Nerve pain | ○ | ○ | ○ | ○ | ○ |
| Nerve sensations (tingling, pins and needles, numbness) | ○ | ○ | ○ | ○ | ○ |
| Neuropathy in feet and hands (weakness, numbness, and pain) | ○ | ○ | ○ | ○ | ○ |
| New allergies | ○ | ○ | ○ | ○ | ○ |
| Night sweats | ○ | ○ | ○ | ○ | ○ |
| Nightmares | ○ | ○ | ○ | ○ | ○ |
| Painful scalp | ○ | ○ | ○ | ○ | ○ |

| | | | | | |
|---|---|---|---|---|---|
| Partial or complete loss of sense of smell | ○ | ○ | ○ | ○ | ○ |
| Partial or complete loss of sense of taste | ○ | ○ | ○ | ○ | ○ |
| Persistent chest pain or pressure | ○ | ○ | ○ | ○ | ○ |
| Personality change (drastic) | ○ | ○ | ○ | ○ | ○ |
| Petechiae (pinpoint rash) | ○ | ○ | ○ | ○ | ○ |
| Phantom smells | ○ | ○ | ○ | ○ | ○ |
| Phlegm in back of throat | ○ | ○ | ○ | ○ | ○ |
| Post-exertional malaise (worsened symptoms or flu-like symptoms after exertion) | ○ | ○ | ○ | ○ | ○ |
| Postnasal drip | ○ | ○ | ○ | ○ | ○ |
| Rash | ○ | ○ | ○ | ○ | ○ |
| Reflux or heartburn | ○ | ○ | ○ | ○ | ○ |
| Runny nose | ○ | ○ | ○ | ○ | ○ |
| Sadness | ○ | ○ | ○ | ○ | ○ |
| Seizures | ○ | ○ | ○ | ○ | ○ |
| Sharp or sudden chest pain | ○ | ○ | ○ | ○ | ○ |
| Shortness of breath or difficulty breathing | ○ | ○ | ○ | ○ | ○ |
| Shortness of breath or exhaustion from bending over | ○ | ○ | ○ | ○ | ○ |
| Sleeping more than normal | ○ | ○ | ○ | ○ | ○ |
| Sore throat | ○ | ○ | ○ | ○ | ○ |
| Spinal issues | ○ | ○ | ○ | ○ | ○ |
| Spikes in blood pressure | ○ | ○ | ○ | ○ | ○ |
| Swollen hands or feet | ○ | ○ | ○ | ○ | ○ |
| Swollen lymph nodes | ○ | ○ | ○ | ○ | ○ |
| Syncope (fainting) | ○ | ○ | ○ | ○ | ○ |
| Tachycardia (rapid heartbeat) at rest | ○ | ○ | ○ | ○ | ○ |
| Tachycardia (rapid heartbeat) after standing up | ○ | ○ | ○ | ○ | ○ |
| Thrush (white fungal infection in the mouth or throat) | ○ | ○ | ○ | ○ | ○ |
| Tinnitus or humming in ears | ○ | ○ | ○ | ○ | ○ |
| Tremors or shakiness | ○ | ○ | ○ | ○ | ○ |
| Upper back pain | ○ | ○ | ○ | ○ | ○ |
| UTI (urinary tract infection) | ○ | ○ | ○ | ○ | ○ |
| Weakened neck | ○ | ○ | ○ | ○ | ○ |

| | | | | | |
|---|---|---|---|---|---|
| Weight gain | ○ | ○ | ○ | ○ | ○ |

**During the past month, how much did the symptom impair your ability to work compared to pre-COVID?**

**If you have not experienced the symptom during the past month, please choose "Not at all."**

| | Not at all | A little bit | Somewhat | Quite a bit | Very much |
|---|---|---|---|---|---|
| Abdominal pain | ○ | ○ | ○ | ○ | ○ |
| Abnormally low temperature | ○ | ○ | ○ | ○ | ○ |
| Acid reflux | ○ | ○ | ○ | ○ | ○ |
| Afternoon or evening fevers/low-grade fevers | ○ | ○ | ○ | ○ | ○ |
| Anemia (low number of red blood cells) | ○ | ○ | ○ | ○ | ○ |
| Anxiety | ○ | ○ | ○ | ○ | ○ |
| Arrythmia (improper beating of the heart due to electrical impulse problems) | ○ | ○ | ○ | ○ | ○ |
| Bilateral neck throbbing around lymph nodes | ○ | ○ | ○ | ○ | ○ |
| Blurry vision | ○ | ○ | ○ | ○ | ○ |
| Bone aches in extremities | ○ | ○ | ○ | ○ | ○ |
| Brain fog | ○ | ○ | ○ | ○ | ○ |
| Brain pressure | ○ | ○ | ○ | ○ | ○ |
| Bulging veins | ○ | ○ | ○ | ○ | ○ |
| Burning sensations | ○ | ○ | ○ | ○ | ○ |
| Bruising of skin | ○ | ○ | ○ | ○ | ○ |
| Calf cramps | ○ | ○ | ○ | ○ | ○ |
| Change in nails (i.e. white spots, brittleness, change in moons) | ○ | ○ | ○ | ○ | ○ |
| Changes in voice | ○ | ○ | ○ | ○ | ○ |
| Changed sense of taste | ○ | ○ | ○ | ○ | ○ |
| Chills but no fever | ○ | ○ | ○ | ○ | ○ |
| Clogged ears | ○ | ○ | ○ | ○ | ○ |
| Cold burning feeling in lungs | ○ | ○ | ○ | ○ | ○ |
| Confusion | ○ | ○ | ○ | ○ | ○ |
| Congested or runny nose | ○ | ○ | ○ | ○ | ○ |
| Constant thirst | ○ | ○ | ○ | ○ | ○ |
| Costochondritis (inflammation of the cartilage that connects a rib to the breastbone) | ○ | ○ | ○ | ○ | ○ |
| Cough | ○ | ○ | ○ | ○ | ○ |

| | | | | | |
|---|---|---|---|---|---|
| Coughing up blood | ○ | ○ | ○ | ○ | ○ |
| Covid toes (tender or itchy rash or chilblains on the toes or foot) | ○ | ○ | ○ | ○ | ○ |
| Cracked or dry lips | ○ | ○ | ○ | ○ | ○ |
| Dental problems (i.e. chipped tooth, tooth loss) | ○ | ○ | ○ | ○ | ○ |
| Diarrhea | ○ | ○ | ○ | ○ | ○ |
| Difficulty concentrating or focusing | ○ | ○ | ○ | ○ | ○ |
| Difficulty sleeping | ○ | ○ | ○ | ○ | ○ |
| Difficulty speaking properly | ○ | ○ | ○ | ○ | ○ |
| Discoloration of the skin (for example: purple or blue on the hands or feet, no blistering) | ○ | ○ | ○ | ○ | ○ |
| Dizziness | ○ | ○ | ○ | ○ | ○ |
| Dry eyes | ○ | ○ | ○ | ○ | ○ |
| Dry or peeling skin | ○ | ○ | ○ | ○ | ○ |
| Dry scalp or dandruff | ○ | ○ | ○ | ○ | ○ |
| Dry throat | ○ | ○ | ○ | ○ | ○ |
| Ear pain/earache | ○ | ○ | ○ | ○ | ○ |
| Elevated thyroid hormones | ○ | ○ | ○ | ○ | ○ |
| Extreme pressure at base of head or occipital nerve | ○ | ○ | ○ | ○ | ○ |
| Eye stye or infection | ○ | ○ | ○ | ○ | ○ |
| Fatigue | ○ | ○ | ○ | ○ | ○ |
| Feeling irritable | ○ | ○ | ○ | ○ | ○ |
| Feeling of burning skin | ○ | ○ | ○ | ○ | ○ |
| Fever or chills | ○ | ○ | ○ | ○ | ○ |
| Floaters or flashes of light in vision | ○ | ○ | ○ | ○ | ○ |
| Foot pain | ○ | ○ | ○ | ○ | ○ |
| GERD (acid reflux) with excessive salivation | ○ | ○ | ○ | ○ | ○ |
| Goiter or lump in throat | ○ | ○ | ○ | ○ | ○ |
| Hair loss | ○ | ○ | ○ | ○ | ○ |
| Hand or wrist pain | ○ | ○ | ○ | ○ | ○ |
| Headache | ○ | ○ | ○ | ○ | ○ |
| Heart palpitations (heart skipping a beat or racing) | ○ | ○ | ○ | ○ | ○ |
| Heat intolerance | ○ | ○ | ○ | ○ | ○ |
| High blood pressure | ○ | ○ | ○ | ○ | ○ |

| | | | | | |
|---|---|---|---|---|---|
| Hormone imbalances | ○ | ○ | ○ | ○ | ○ |
| "Hot" blood rush | ○ | ○ | ○ | ○ | ○ |
| Inability to cry | ○ | ○ | ○ | ○ | ○ |
| Inability to exercise or be active | ○ | ○ | ○ | ○ | ○ |
| Inability to yawn | ○ | ○ | ○ | ○ | ○ |
| Internal tremors or buzzing/vibration | ○ | ○ | ○ | ○ | ○ |
| Irregular or skipped menstrual cycles | ○ | ○ | ○ | ○ | ○ |
| Jaw pain | ○ | ○ | ○ | ○ | ○ |
| Joint pain | ○ | ○ | ○ | ○ | ○ |
| Kidney issues or protein in urine | ○ | ○ | ○ | ○ | ○ |
| Kidney pain | ○ | ○ | ○ | ○ | ○ |
| Low blood oxygen | ○ | ○ | ○ | ○ | ○ |
| Low blood pressure | ○ | ○ | ○ | ○ | ○ |
| Lower back pain | ○ | ○ | ○ | ○ | ○ |
| Loss of appetite | ○ | ○ | ○ | ○ | ○ |
| Loss of hearing | ○ | ○ | ○ | ○ | ○ |
| Loss or decrease in quality of vision | ○ | ○ | ○ | ○ | ○ |
| Lump in throat/difficulty swallowing | ○ | ○ | ○ | ○ | ○ |
| Memory problems | ○ | ○ | ○ | ○ | ○ |
| Menstrual cycles that are heavier or lighter than normal | ○ | ○ | ○ | ○ | ○ |
| Mid-back pain at base of ribs | ○ | ○ | ○ | ○ | ○ |
| Mouth sores or sore tongue | ○ | ○ | ○ | ○ | ○ |
| Muscle or body aches | ○ | ○ | ○ | ○ | ○ |
| Muscle twitching | ○ | ○ | ○ | ○ | ○ |
| Nausea or vomiting | ○ | ○ | ○ | ○ | ○ |
| Neck muscle pain | ○ | ○ | ○ | ○ | ○ |
| Nerve pain | ○ | ○ | ○ | ○ | ○ |
| Nerve sensations (tingling, pins and needles, numbness) | ○ | ○ | ○ | ○ | ○ |
| Neuropathy in feet and hands (weakness, numbness, and pain) | ○ | ○ | ○ | ○ | ○ |
| New allergies | ○ | ○ | ○ | ○ | ○ |
| Night sweats | ○ | ○ | ○ | ○ | ○ |
| Nightmares | ○ | ○ | ○ | ○ | ○ |
| Painful scalp | ○ | ○ | ○ | ○ | ○ |

| | | | | | |
|---|---|---|---|---|---|
| Partial or complete loss of sense of smell | ○ | ○ | ○ | ○ | ○ |
| Partial or complete loss of sense of taste | ○ | ○ | ○ | ○ | ○ |
| Persistent chest pain or pressure | ○ | ○ | ○ | ○ | ○ |
| Personality change (drastic) | ○ | ○ | ○ | ○ | ○ |
| Petechiae (pinpoint rash) | ○ | ○ | ○ | ○ | ○ |
| Phantom smells | ○ | ○ | ○ | ○ | ○ |
| Phlegm in back of throat | ○ | ○ | ○ | ○ | ○ |
| Post-exertional malaise (worsened symptoms or flu-like symptoms after exertion) | ○ | ○ | ○ | ○ | ○ |
| Postnasal drip | ○ | ○ | ○ | ○ | ○ |
| Rash | ○ | ○ | ○ | ○ | ○ |
| Reflux or heartburn | ○ | ○ | ○ | ○ | ○ |
| Runny nose | ○ | ○ | ○ | ○ | ○ |
| Sadness | ○ | ○ | ○ | ○ | ○ |
| Seizures | ○ | ○ | ○ | ○ | ○ |
| Sharp or sudden chest pain | ○ | ○ | ○ | ○ | ○ |
| Shortness of breath or difficulty breathing | ○ | ○ | ○ | ○ | ○ |
| Shortness of breath or exhaustion from bending over | ○ | ○ | ○ | ○ | ○ |
| Sleeping more than normal | ○ | ○ | ○ | ○ | ○ |
| Sore throat | ○ | ○ | ○ | ○ | ○ |
| Spinal issues | ○ | ○ | ○ | ○ | ○ |
| Spikes in blood pressure | ○ | ○ | ○ | ○ | ○ |
| Swollen hands or feet | ○ | ○ | ○ | ○ | ○ |
| Swollen lymph nodes | ○ | ○ | ○ | ○ | ○ |
| Syncope (fainting) | ○ | ○ | ○ | ○ | ○ |
| Tachycardia (rapid heartbeat) at rest | ○ | ○ | ○ | ○ | ○ |
| Tachycardia (rapid heartbeat) after standing up | ○ | ○ | ○ | ○ | ○ |
| Thrush (white fungal infection in the mouth or throat) | ○ | ○ | ○ | ○ | ○ |
| Tinnitus or humming in ears | ○ | ○ | ○ | ○ | ○ |
| Tremors or shakiness | ○ | ○ | ○ | ○ | ○ |
| Upper back pain | ○ | ○ | ○ | ○ | ○ |
| UTI (urinary tract infection) | ○ | ○ | ○ | ○ | ○ |
| Weakened neck | ○ | ○ | ○ | ○ | ○ |

| | | | | | |
|---|---|---|---|---|---|
| Weight gain | ◯ | ◯ | ◯ | ◯ | ◯ |

**During the past month, how much did the symptom impair your social or family functioning compared to pre-COVID?**

**If you have not experienced the symptom during the past month, please choose "Not at all."**

| | Not at all | A little bit | Somewhat | Quite a bit | Very much |
|---|---|---|---|---|---|
| Abdominal pain | ○ | ○ | ○ | ○ | ○ |
| Abnormally low temperature | ○ | ○ | ○ | ○ | ○ |
| Acid reflux | ○ | ○ | ○ | ○ | ○ |
| Afternoon or evening fevers/low-grade fevers | ○ | ○ | ○ | ○ | ○ |
| Anemia (low number of red blood cells) | ○ | ○ | ○ | ○ | ○ |
| Anxiety | ○ | ○ | ○ | ○ | ○ |
| Arrythmia (improper beating of the heart due to electrical impulse problems) | ○ | ○ | ○ | ○ | ○ |
| Bilateral neck throbbing around lymph nodes | ○ | ○ | ○ | ○ | ○ |
| Blurry vision | ○ | ○ | ○ | ○ | ○ |
| Bone aches in extremities | ○ | ○ | ○ | ○ | ○ |
| Brain fog | ○ | ○ | ○ | ○ | ○ |
| Brain pressure | ○ | ○ | ○ | ○ | ○ |
| Bulging veins | ○ | ○ | ○ | ○ | ○ |
| Burning sensations | ○ | ○ | ○ | ○ | ○ |
| Bruising of skin | ○ | ○ | ○ | ○ | ○ |
| Calf cramps | ○ | ○ | ○ | ○ | ○ |
| Change in nails (i.e. white spots, brittleness, change in moons) | ○ | ○ | ○ | ○ | ○ |
| Changes in voice | ○ | ○ | ○ | ○ | ○ |
| Changed sense of taste | ○ | ○ | ○ | ○ | ○ |
| Chills but no fever | ○ | ○ | ○ | ○ | ○ |
| Clogged ears | ○ | ○ | ○ | ○ | ○ |
| Cold burning feeling in lungs | ○ | ○ | ○ | ○ | ○ |
| Confusion | ○ | ○ | ○ | ○ | ○ |
| Congested or runny nose | ○ | ○ | ○ | ○ | ○ |
| Constant thirst | ○ | ○ | ○ | ○ | ○ |
| Costochondritis (inflammation of the cartilage that connects a rib to the breastbone) | ○ | ○ | ○ | ○ | ○ |
| Cough | ○ | ○ | ○ | ○ | ○ |

| | | | | | |
|---|---|---|---|---|---|
| Coughing up blood | ◯ | ◯ | ◯ | ◯ | ◯ |
| Covid toes (tender or itchy rash or chilblains on the toes or foot) | ◯ | ◯ | ◯ | ◯ | ◯ |
| Cracked or dry lips | ◯ | ◯ | ◯ | ◯ | ◯ |
| Dental problems (i.e. chipped tooth, tooth loss) | ◯ | ◯ | ◯ | ◯ | ◯ |
| Diarrhea | ◯ | ◯ | ◯ | ◯ | ◯ |
| Difficulty concentrating or focusing | ◯ | ◯ | ◯ | ◯ | ◯ |
| Difficulty sleeping | ◯ | ◯ | ◯ | ◯ | ◯ |
| Difficulty speaking properly | ◯ | ◯ | ◯ | ◯ | ◯ |
| Discoloration of the skin (for example: purple or blue on the hands or feet, no blistering) | ◯ | ◯ | ◯ | ◯ | ◯ |
| Dizziness | ◯ | ◯ | ◯ | ◯ | ◯ |
| Dry eyes | ◯ | ◯ | ◯ | ◯ | ◯ |
| Dry or peeling skin | ◯ | ◯ | ◯ | ◯ | ◯ |
| Dry scalp or dandruff | ◯ | ◯ | ◯ | ◯ | ◯ |
| Dry throat | ◯ | ◯ | ◯ | ◯ | ◯ |
| Ear pain/earache | ◯ | ◯ | ◯ | ◯ | ◯ |
| Elevated thyroid hormones | ◯ | ◯ | ◯ | ◯ | ◯ |
| Extreme pressure at base of head or occipital nerve | ◯ | ◯ | ◯ | ◯ | ◯ |
| Eye stye or infection | ◯ | ◯ | ◯ | ◯ | ◯ |
| Fatigue | ◯ | ◯ | ◯ | ◯ | ◯ |
| Feeling irritable | ◯ | ◯ | ◯ | ◯ | ◯ |
| Feeling of burning skin | ◯ | ◯ | ◯ | ◯ | ◯ |
| Fever or chills | ◯ | ◯ | ◯ | ◯ | ◯ |
| Floaters or flashes of light in vision | ◯ | ◯ | ◯ | ◯ | ◯ |
| Foot pain | ◯ | ◯ | ◯ | ◯ | ◯ |
| GERD (acid reflux) with excessive salivation | ◯ | ◯ | ◯ | ◯ | ◯ |
| Goiter or lump in throat | ◯ | ◯ | ◯ | ◯ | ◯ |
| Hair loss | ◯ | ◯ | ◯ | ◯ | ◯ |
| Hand or wrist pain | ◯ | ◯ | ◯ | ◯ | ◯ |
| Headache | ◯ | ◯ | ◯ | ◯ | ◯ |
| Heart palpitations (heart skipping a beat or racing) | ◯ | ◯ | ◯ | ◯ | ◯ |
| Heat intolerance | ◯ | ◯ | ◯ | ◯ | ◯ |
| High blood pressure | ◯ | ◯ | ◯ | ◯ | ◯ |

| | | | | | |
|---|---|---|---|---|---|
| Hormone imbalances | ◯ | ◯ | ◯ | ◯ | ◯ |
| "Hot" blood rush | ◯ | ◯ | ◯ | ◯ | ◯ |
| Inability to cry | ◯ | ◯ | ◯ | ◯ | ◯ |
| Inability to exercise or be active | ◯ | ◯ | ◯ | ◯ | ◯ |
| Inability to yawn | ◯ | ◯ | ◯ | ◯ | ◯ |
| Internal tremors or buzzing/vibration | ◯ | ◯ | ◯ | ◯ | ◯ |
| Irregular or skipped menstrual cycles | ◯ | ◯ | ◯ | ◯ | ◯ |
| Jaw pain | ◯ | ◯ | ◯ | ◯ | ◯ |
| Joint pain | ◯ | ◯ | ◯ | ◯ | ◯ |
| Kidney issues or protein in urine | ◯ | ◯ | ◯ | ◯ | ◯ |
| Kidney pain | ◯ | ◯ | ◯ | ◯ | ◯ |
| Low blood oxygen | ◯ | ◯ | ◯ | ◯ | ◯ |
| Low blood pressure | ◯ | ◯ | ◯ | ◯ | ◯ |
| Lower back pain | ◯ | ◯ | ◯ | ◯ | ◯ |
| Loss of appetite | ◯ | ◯ | ◯ | ◯ | ◯ |
| Loss of hearing | ◯ | ◯ | ◯ | ◯ | ◯ |
| Loss or decrease in quality of vision | ◯ | ◯ | ◯ | ◯ | ◯ |
| Lump in throat/difficulty swallowing | ◯ | ◯ | ◯ | ◯ | ◯ |
| Memory problems | ◯ | ◯ | ◯ | ◯ | ◯ |
| Menstrual cycles that are heavier or lighter than normal | ◯ | ◯ | ◯ | ◯ | ◯ |
| Mid-back pain at base of ribs | ◯ | ◯ | ◯ | ◯ | ◯ |
| Mouth sores or sore tongue | ◯ | ◯ | ◯ | ◯ | ◯ |
| Muscle or body aches | ◯ | ◯ | ◯ | ◯ | ◯ |
| Muscle twitching | ◯ | ◯ | ◯ | ◯ | ◯ |
| Nausea or vomiting | ◯ | ◯ | ◯ | ◯ | ◯ |
| Neck muscle pain | ◯ | ◯ | ◯ | ◯ | ◯ |
| Nerve pain | ◯ | ◯ | ◯ | ◯ | ◯ |
| Nerve sensations (tingling, pins and needles, numbness) | ◯ | ◯ | ◯ | ◯ | ◯ |
| Neuropathy in feet and hands (weakness, numbness, and pain) | ◯ | ◯ | ◯ | ◯ | ◯ |
| New allergies | ◯ | ◯ | ◯ | ◯ | ◯ |
| Night sweats | ◯ | ◯ | ◯ | ◯ | ◯ |
| Nightmares | ◯ | ◯ | ◯ | ◯ | ◯ |
| Painful scalp | ◯ | ◯ | ◯ | ◯ | ◯ |

| | | | | | |
|---|---|---|---|---|---|
| Partial or complete loss of sense of smell | ○ | ○ | ○ | ○ | ○ |
| Partial or complete loss of sense of taste | ○ | ○ | ○ | ○ | ○ |
| Persistent chest pain or pressure | ○ | ○ | ○ | ○ | ○ |
| Personality change (drastic) | ○ | ○ | ○ | ○ | ○ |
| Petechiae (pinpoint rash) | ○ | ○ | ○ | ○ | ○ |
| Phantom smells | ○ | ○ | ○ | ○ | ○ |
| Phlegm in back of throat | ○ | ○ | ○ | ○ | ○ |
| Post-exertional malaise (worsened symptoms or flu-like symptoms after exertion) | ○ | ○ | ○ | ○ | ○ |
| Postnasal drip | ○ | ○ | ○ | ○ | ○ |
| Rash | ○ | ○ | ○ | ○ | ○ |
| Reflux or heartburn | ○ | ○ | ○ | ○ | ○ |
| Runny nose | ○ | ○ | ○ | ○ | ○ |
| Sadness | ○ | ○ | ○ | ○ | ○ |
| Seizures | ○ | ○ | ○ | ○ | ○ |
| Sharp or sudden chest pain | ○ | ○ | ○ | ○ | ○ |
| Shortness of breath or difficulty breathing | ○ | ○ | ○ | ○ | ○ |
| Shortness of breath or exhaustion from bending over | ○ | ○ | ○ | ○ | ○ |
| Sleeping more than normal | ○ | ○ | ○ | ○ | ○ |
| Sore throat | ○ | ○ | ○ | ○ | ○ |
| Spinal issues | ○ | ○ | ○ | ○ | ○ |
| Spikes in blood pressure | ○ | ○ | ○ | ○ | ○ |
| Swollen hands or feet | ○ | ○ | ○ | ○ | ○ |
| Swollen lymph nodes | ○ | ○ | ○ | ○ | ○ |
| Syncope (fainting) | ○ | ○ | ○ | ○ | ○ |
| Tachycardia (rapid heartbeat) at rest | ○ | ○ | ○ | ○ | ○ |
| Tachycardia (rapid heartbeat) after standing up | ○ | ○ | ○ | ○ | ○ |
| Thrush (white fungal infection in the mouth or throat) | ○ | ○ | ○ | ○ | ○ |
| Tinnitus or humming in ears | ○ | ○ | ○ | ○ | ○ |
| Tremors or shakiness | ○ | ○ | ○ | ○ | ○ |
| Upper back pain | ○ | ○ | ○ | ○ | ○ |
| UTI (urinary tract infection) | ○ | ○ | ○ | ○ | ○ |
| Weakened neck | ○ | ○ | ○ | ○ | ○ |

| | | | | | |
|---|---|---|---|---|---|
| Weight gain | ○ | ○ | ○ | ○ | ○ |

Are there any symptoms not listed above that you have experienced within the past month?

*Please list all if so

__________________________________

Have you visited the hospital or been hospitalized in the past month for COVID-19 symptoms?

○ Yes ______
○ No

Please elaborate on your hospitalization (cause of hospitalization; relation, if any, to COVID-19; and health since hospitalization).

__________________________________

#### During the past month

| | 76-100% of health before COVID-19 | 51-75% of health before COVID-19 | 26-50% of health before COVID-19 | 0-25% of health before COVID-19 |
|---|---|---|---|---|
| On your best days, would you say that you are at | ◯ | ◯ | ◯ | ◯ |
| On your worst days, would you say that you are at | ◯ | ◯ | ◯ | ◯ |

Compared to your health before you received the first dose of your vaccine on [vaccine_arm_1][v_v_vax_date], would you say that your health is currently

◯ Better
◯ Worse
◯ The same
◯ Don't know

Please, in the space below, share any information about your experience with COVID-19 or the vaccine that we might have missed.

__________________________________

### Survey 4

Thank you for taking Survey 4, the final survey for the Yale COVID Recovery Study!

Please remember that you may stop this survey and come back to it at any point.

You may also find it helpful to have the below items ready as you complete the survey. If you do not have these items, please still fill in the information as best you can.

Testing results: dates, type (PCR, antigen) and result of tests. If you do not remember the exact date, the estimated date is enough.Symptom time and severity: your symptom log. You will be asked to pick symptoms you have had since you began having COVID-19 symptoms.

| | |
|---|---|
| During the past 6 weeks, have you visited the hospital or been hospitalized for COVID-19 symptoms? | ◯ Yes ______<br>◯ No |
| What were you hospitalized for? | __________________________________ |

On your previous survey, you responded that you have experienced the following symptoms due to COVID-19 at some point in the past.

The following questions will ask about how these symptoms have changed, if at all. Please respond based on your experiences during the past 6 weeks.

#### During the past 6 weeks, as compared to before, this symptom has been

| | Better | Worse | The same |
|---|---|---|---|
| Abdominal pain | ◯ | ◯ | ◯ |
| Abnormally low temperature | ◯ | ◯ | ◯ |
| Acid reflux | ◯ | ◯ | ◯ |
| Afternoon or evening fevers/low-grade fevers | ◯ | ◯ | ◯ |
| Anemia (low number of red blood cells) | ◯ | ◯ | ◯ |
| Anxiety | ◯ | ◯ | ◯ |
| Arrythmia (improper beating of the heart due to electrical impulse problems) | ◯ | ◯ | ◯ |
| Bilateral neck throbbing around lymph nodes | ◯ | ◯ | ◯ |
| Blurry vision | ◯ | ◯ | ◯ |
| Bone aches in extremities | ◯ | ◯ | ◯ |
| Brain fog | ◯ | ◯ | ◯ |
| Brain pressure | ◯ | ◯ | ◯ |
| Bulging veins | ◯ | ◯ | ◯ |
| Burning sensations | ◯ | ◯ | ◯ |
| Bruising of skin | ◯ | ◯ | ◯ |
| Calf cramps | ◯ | ◯ | ◯ |
| Change in nails (i.e. white spots, brittleness, change in moons) | ◯ | ◯ | ◯ |
| Changes in voice | ◯ | ◯ | ◯ |
| Changed sense of taste | ◯ | ◯ | ◯ |
| Chills but no fever | ◯ | ◯ | ◯ |
| Clogged ears | ◯ | ◯ | ◯ |
| Cold burning feeling in lungs | ◯ | ◯ | ◯ |
| Confusion | ◯ | ◯ | ◯ |
| Congested or runny nose | ◯ | ◯ | ◯ |
| Constant thirst | ◯ | ◯ | ◯ |
| Costochondritis (inflammation of the cartilage that connects a rib to the breastbone) | ◯ | ◯ | ◯ |
| Cough | ◯ | ◯ | ◯ |
| Coughing up blood | ◯ | ◯ | ◯ |
| Covid toes (tender or itchy rash or chilblains on the toes or foot) | ◯ | ◯ | ◯ |

| | | | |
|---|---|---|---|
| Cracked or dry lips | ○ | ○ | ○ |
| Dental problems (i.e. chipped tooth, tooth loss) | ○ | ○ | ○ |
| Diarrhea | ○ | ○ | ○ |
| Difficulty concentrating or focusing | ○ | ○ | ○ |
| Difficulty sleeping | ○ | ○ | ○ |
| Difficulty speaking properly | ○ | ○ | ○ |
| Discoloration of the skin (for example: purple or blue on the hands or feet, no blistering) | ○ | ○ | ○ |
| Dizziness | ○ | ○ | ○ |
| Dry eyes | ○ | ○ | ○ |
| Dry or peeling skin | ○ | ○ | ○ |
| Dry scalp or dandruff | ○ | ○ | ○ |
| Dry throat | ○ | ○ | ○ |
| Ear pain/earache | ○ | ○ | ○ |
| Elevated thyroid hormones | ○ | ○ | ○ |
| Extreme pressure at base of head or occipital nerve | ○ | ○ | ○ |
| Eye stye or infection | ○ | ○ | ○ |
| Fatigue | ○ | ○ | ○ |
| Feeling irritable | ○ | ○ | ○ |
| Feeling of burning skin | ○ | ○ | ○ |
| Fever or chills | ○ | ○ | ○ |
| Floaters or flashes of light in vision | ○ | ○ | ○ |
| Foot pain | ○ | ○ | ○ |
| GERD (acid reflux) with excessive salivation | ○ | ○ | ○ |
| Goiter or lump in throat | ○ | ○ | ○ |
| Hair loss | ○ | ○ | ○ |
| Hand or wrist pain | ○ | ○ | ○ |
| Headache | ○ | ○ | ○ |
| Heart palpitations (heart skipping a beat or racing) | ○ | ○ | ○ |
| Heat intolerance | ○ | ○ | ○ |
| High blood pressure | ○ | ○ | ○ |
| Hormone imbalances | ○ | ○ | ○ |
| "Hot" blood rush | ○ | ○ | ○ |
| Inability to cry | ○ | ○ | ○ |

| | | | |
|---|---|---|---|
| Inability to exercise or be active | ○ | ○ | ○ |
| Inability to yawn | ○ | ○ | ○ |
| Internal tremors or buzzing/vibration | ○ | ○ | ○ |
| Irregular or skipped menstrual cycles | ○ | ○ | ○ |
| Jaw pain | ○ | ○ | ○ |
| Joint pain | ○ | ○ | ○ |
| Kidney issues or protein in urine | ○ | ○ | ○ |
| Kidney pain | ○ | ○ | ○ |
| Low blood oxygen | ○ | ○ | ○ |
| Low blood pressure | ○ | ○ | ○ |
| Lower back pain | ○ | ○ | ○ |
| Loss of appetite | ○ | ○ | ○ |
| Loss of hearing | ○ | ○ | ○ |
| Loss or decrease in quality of vision | ○ | ○ | ○ |
| Lump in throat/difficulty swallowing | ○ | ○ | ○ |
| Memory problems | ○ | ○ | ○ |
| Menstrual cycles that are heavier or lighter than normal | ○ | ○ | ○ |
| Mid-back pain at base of ribs | ○ | ○ | ○ |
| Mouth sores or sore tongue | ○ | ○ | ○ |
| Muscle or body aches | ○ | ○ | ○ |
| Muscle twitching | ○ | ○ | ○ |
| Nausea or vomiting | ○ | ○ | ○ |
| Neck muscle pain | ○ | ○ | ○ |
| Nerve pain | ○ | ○ | ○ |
| Nerve sensations (tingling, pins and needles, numbness) | ○ | ○ | ○ |
| Neuropathy in feet and hands (weakness, numbness, and pain) | ○ | ○ | ○ |
| New allergies | ○ | ○ | ○ |
| Night sweats | ○ | ○ | ○ |
| Nightmares | ○ | ○ | ○ |
| Painful scalp | ○ | ○ | ○ |
| Partial or complete loss of sense of smell | ○ | ○ | ○ |
| Partial or complete loss of sense of taste | ○ | ○ | ○ |

| | | | |
|---|---|---|---|
| Persistent chest pain or pressure | ○ | ○ | ○ |
| Personality change (drastic) | ○ | ○ | ○ |
| Petechiae (pinpoint rash) | ○ | ○ | ○ |
| Phantom smells | ○ | ○ | ○ |
| Phlegm in back of throat | ○ | ○ | ○ |
| Post-exertional malaise (worsened symptoms or flu-like symptoms after exertion) | ○ | ○ | ○ |
| Postnasal drip | ○ | ○ | ○ |
| Rash | ○ | ○ | ○ |
| Reflux or heartburn | ○ | ○ | ○ |
| Runny nose | ○ | ○ | ○ |
| Sadness | ○ | ○ | ○ |
| Seizures | ○ | ○ | ○ |
| Sharp or sudden chest pain | ○ | ○ | ○ |
| Shortness of breath or difficulty breathing | ○ | ○ | ○ |
| Shortness of breath or exhaustion from bending over | ○ | ○ | ○ |
| Sleeping more than normal | ○ | ○ | ○ |
| Sore throat | ○ | ○ | ○ |
| Spinal issues | ○ | ○ | ○ |
| Spikes in blood pressure | ○ | ○ | ○ |
| Swollen hands or feet | ○ | ○ | ○ |
| Swollen lymph nodes | ○ | ○ | ○ |
| Syncope (fainting) | ○ | ○ | ○ |
| Tachycardia (rapid heartbeat) at rest | ○ | ○ | ○ |
| Tachycardia (rapid heartbeat) after standing up | ○ | ○ | ○ |
| Thrush (white fungal infection in the mouth or throat) | ○ | ○ | ○ |
| Tinnitus or humming in ears | ○ | ○ | ○ |
| Tremors or shakiness | ○ | ○ | ○ |
| Upper back pain | ○ | ○ | ○ |
| UTI (urinary tract infection) | ○ | ○ | ○ |
| Weakened neck | ○ | ○ | ○ |
| Weight gain | ○ | ○ | ○ |

#### During the past 6 weeks, I have experienced this symptom, on average

| | Never--it has gone away completely | Once a month | Once or twice a week | 3 or 4 times a week | Most days | All the time |
|---|---|---|---|---|---|---|
| Abdominal pain | ○ | ○ | ○ | ○ | ○ | ○ |
| Abnormally low temperature | ○ | ○ | ○ | ○ | ○ | ○ |
| Acid reflux | ○ | ○ | ○ | ○ | ○ | ○ |
| Afternoon or evening fevers/low-grade fevers | ○ | ○ | ○ | ○ | ○ | ○ |
| Anemia (low number of red blood cells) | ○ | ○ | ○ | ○ | ○ | ○ |
| Anxiety | ○ | ○ | ○ | ○ | ○ | ○ |
| Arrythmia (improper beating of the heart due to electrical impulse problems) | ○ | ○ | ○ | ○ | ○ | ○ |
| Bilateral neck throbbing around lymph nodes | ○ | ○ | ○ | ○ | ○ | ○ |
| Blurry vision | ○ | ○ | ○ | ○ | ○ | ○ |
| Bone aches in extremities | ○ | ○ | ○ | ○ | ○ | ○ |
| Brain fog | ○ | ○ | ○ | ○ | ○ | ○ |
| Brain pressure | ○ | ○ | ○ | ○ | ○ | ○ |
| Bulging veins | ○ | ○ | ○ | ○ | ○ | ○ |
| Burning sensations | ○ | ○ | ○ | ○ | ○ | ○ |
| Bruising of skin | ○ | ○ | ○ | ○ | ○ | ○ |
| Calf cramps | ○ | ○ | ○ | ○ | ○ | ○ |
| Change in nails (i.e. white spots, brittleness, change in moons) | ○ | ○ | ○ | ○ | ○ | ○ |
| Changes in voice | ○ | ○ | ○ | ○ | ○ | ○ |
| Changed sense of taste | ○ | ○ | ○ | ○ | ○ | ○ |
| Chills but no fever | ○ | ○ | ○ | ○ | ○ | ○ |
| Clogged ears | ○ | ○ | ○ | ○ | ○ | ○ |
| Cold burning feeling in lungs | ○ | ○ | ○ | ○ | ○ | ○ |
| Confusion | ○ | ○ | ○ | ○ | ○ | ○ |
| Congested or runny nose | ○ | ○ | ○ | ○ | ○ | ○ |
| Constant thirst | ○ | ○ | ○ | ○ | ○ | ○ |
| Costochondritis (inflammation of the cartilage that connects a rib to the breastbone) | ○ | ○ | ○ | ○ | ○ | ○ |
| Cough | ○ | ○ | ○ | ○ | ○ | ○ |
| Coughing up blood | ○ | ○ | ○ | ○ | ○ | ○ |

| | | | | | | |
|---|---|---|---|---|---|---|
| Covid toes (tender or itchy rash or chilblains on the toes or foot) | ○ | ○ | ○ | ○ | ○ | ○ |
| Cracked or dry lips | ○ | ○ | ○ | ○ | ○ | ○ |
| Dental problems (i.e. chipped tooth, tooth loss) | ○ | ○ | ○ | ○ | ○ | ○ |
| Diarrhea | ○ | ○ | ○ | ○ | ○ | ○ |
| Difficulty concentrating or focusing | ○ | ○ | ○ | ○ | ○ | ○ |
| Difficulty sleeping | ○ | ○ | ○ | ○ | ○ | ○ |
| Difficulty speaking properly | ○ | ○ | ○ | ○ | ○ | ○ |
| Discoloration of the skin (for example: purple or blue on the hands or feet, no blistering) | ○ | ○ | ○ | ○ | ○ | ○ |
| Dizziness | ○ | ○ | ○ | ○ | ○ | ○ |
| Dry eyes | ○ | ○ | ○ | ○ | ○ | ○ |
| Dry or peeling skin | ○ | ○ | ○ | ○ | ○ | ○ |
| Dry scalp or dandruff | ○ | ○ | ○ | ○ | ○ | ○ |
| Dry throat | ○ | ○ | ○ | ○ | ○ | ○ |
| Ear pain/earache | ○ | ○ | ○ | ○ | ○ | ○ |
| Elevated thyroid hormones | ○ | ○ | ○ | ○ | ○ | ○ |
| Extreme pressure at base of head or occipital nerve | ○ | ○ | ○ | ○ | ○ | ○ |
| Eye stye or infection | ○ | ○ | ○ | ○ | ○ | ○ |
| Fatigue | ○ | ○ | ○ | ○ | ○ | ○ |
| Feeling irritable | ○ | ○ | ○ | ○ | ○ | ○ |
| Feeling of burning skin | ○ | ○ | ○ | ○ | ○ | ○ |
| Fever or chills | ○ | ○ | ○ | ○ | ○ | ○ |
| Floaters or flashes of light in vision | ○ | ○ | ○ | ○ | ○ | ○ |
| Foot pain | ○ | ○ | ○ | ○ | ○ | ○ |
| GERD (acid reflux) with excessive salivation | ○ | ○ | ○ | ○ | ○ | ○ |
| Goiter or lump in throat | ○ | ○ | ○ | ○ | ○ | ○ |
| Hair loss | ○ | ○ | ○ | ○ | ○ | ○ |
| Hand or wrist pain | ○ | ○ | ○ | ○ | ○ | ○ |
| Headache | ○ | ○ | ○ | ○ | ○ | ○ |
| Heart palpitations (heart skipping a beat or racing) | ○ | ○ | ○ | ○ | ○ | ○ |
| Heat intolerance | ○ | ○ | ○ | ○ | ○ | ○ |
| High blood pressure | ○ | ○ | ○ | ○ | ○ | ○ |
| Hormone imbalances | ○ | ○ | ○ | ○ | ○ | ○ |

| | | | | | | |
|---|---|---|---|---|---|---|
| "Hot" blood rush | ○ | ○ | ○ | ○ | ○ | ○ |
| Inability to cry | ○ | ○ | ○ | ○ | ○ | ○ |
| Inability to exercise or be active | ○ | ○ | ○ | ○ | ○ | ○ |
| Inability to yawn | ○ | ○ | ○ | ○ | ○ | ○ |
| Internal tremors or buzzing/vibration | ○ | ○ | ○ | ○ | ○ | ○ |
| Irregular or skipped menstrual cycles | ○ | ○ | ○ | ○ | ○ | ○ |
| Jaw pain | ○ | ○ | ○ | ○ | ○ | ○ |
| Joint pain | ○ | ○ | ○ | ○ | ○ | ○ |
| Kidney issues or protein in urine | ○ | ○ | ○ | ○ | ○ | ○ |
| Kidney pain | ○ | ○ | ○ | ○ | ○ | ○ |
| Low blood oxygen | ○ | ○ | ○ | ○ | ○ | ○ |
| Low blood pressure | ○ | ○ | ○ | ○ | ○ | ○ |
| Lower back pain | ○ | ○ | ○ | ○ | ○ | ○ |
| Loss of appetite | ○ | ○ | ○ | ○ | ○ | ○ |
| Loss of hearing | ○ | ○ | ○ | ○ | ○ | ○ |
| Loss or decrease in quality of vision | ○ | ○ | ○ | ○ | ○ | ○ |
| Lump in throat/difficulty swallowing | ○ | ○ | ○ | ○ | ○ | ○ |
| Memory problems | ○ | ○ | ○ | ○ | ○ | ○ |
| Menstrual cycles that are heavier or lighter than normal | ○ | ○ | ○ | ○ | ○ | ○ |
| Mid-back pain at base of ribs | ○ | ○ | ○ | ○ | ○ | ○ |
| Mouth sores or sore tongue | ○ | ○ | ○ | ○ | ○ | ○ |
| Muscle or body aches | ○ | ○ | ○ | ○ | ○ | ○ |
| Muscle twitching | ○ | ○ | ○ | ○ | ○ | ○ |
| Nausea or vomiting | ○ | ○ | ○ | ○ | ○ | ○ |
| Neck muscle pain | ○ | ○ | ○ | ○ | ○ | ○ |
| Nerve pain | ○ | ○ | ○ | ○ | ○ | ○ |
| Nerve sensations (tingling, pins and needles, numbness) | ○ | ○ | ○ | ○ | ○ | ○ |
| Neuropathy in feet and hands (weakness, numbness, and pain) | ○ | ○ | ○ | ○ | ○ | ○ |
| New allergies | ○ | ○ | ○ | ○ | ○ | ○ |
| Night sweats | ○ | ○ | ○ | ○ | ○ | ○ |
| Nightmares | ○ | ○ | ○ | ○ | ○ | ○ |
| Painful scalp | ○ | ○ | ○ | ○ | ○ | ○ |
| Partial or complete loss of sense of smell | ○ | ○ | ○ | ○ | ○ | ○ |

| | | | | | | |
|---|---|---|---|---|---|---|
| Partial or complete loss of sense of taste | ○ | ○ | ○ | ○ | ○ | ○ |
| Persistent chest pain or pressure | ○ | ○ | ○ | ○ | ○ | ○ |
| Personality change (drastic) | ○ | ○ | ○ | ○ | ○ | ○ |
| Petechiae (pinpoint rash) | ○ | ○ | ○ | ○ | ○ | ○ |
| Phantom smells | ○ | ○ | ○ | ○ | ○ | ○ |
| Phlegm in back of throat | ○ | ○ | ○ | ○ | ○ | ○ |
| Post-exertional malaise (worsened symptoms or flu-like symptoms after exertion) | ○ | ○ | ○ | ○ | ○ | ○ |
| Postnasal drip | ○ | ○ | ○ | ○ | ○ | ○ |
| Rash | ○ | ○ | ○ | ○ | ○ | ○ |
| Reflux or heartburn | ○ | ○ | ○ | ○ | ○ | ○ |
| Runny nose | ○ | ○ | ○ | ○ | ○ | ○ |
| Sadness | ○ | ○ | ○ | ○ | ○ | ○ |
| Seizures | ○ | ○ | ○ | ○ | ○ | ○ |
| Sharp or sudden chest pain | ○ | ○ | ○ | ○ | ○ | ○ |
| Shortness of breath or difficulty breathing | ○ | ○ | ○ | ○ | ○ | ○ |
| Shortness of breath or exhaustion from bending over | ○ | ○ | ○ | ○ | ○ | ○ |
| Sleeping more than normal | ○ | ○ | ○ | ○ | ○ | ○ |
| Sore throat | ○ | ○ | ○ | ○ | ○ | ○ |
| Spinal issues | ○ | ○ | ○ | ○ | ○ | ○ |
| Spikes in blood pressure | ○ | ○ | ○ | ○ | ○ | ○ |
| Swollen hands or feet | ○ | ○ | ○ | ○ | ○ | ○ |
| Swollen lymph nodes | ○ | ○ | ○ | ○ | ○ | ○ |
| Syncope (fainting) | ○ | ○ | ○ | ○ | ○ | ○ |
| Tachycardia (rapid heartbeat) at rest | ○ | ○ | ○ | ○ | ○ | ○ |
| Tachycardia (rapid heartbeat) after standing up | ○ | ○ | ○ | ○ | ○ | ○ |
| Thrush (white fungal infection in the mouth or throat) | ○ | ○ | ○ | ○ | ○ | ○ |
| Tinnitus or humming in ears | ○ | ○ | ○ | ○ | ○ | ○ |
| Tremors or shakiness | ○ | ○ | ○ | ○ | ○ | ○ |
| Upper back pain | ○ | ○ | ○ | ○ | ○ | ○ |
| UTI (urinary tract infection) | ○ | ○ | ○ | ○ | ○ | ○ |
| Weakened neck | ○ | ○ | ○ | ○ | ○ | ○ |
| Weight gain | ○ | ○ | ○ | ○ | ○ | ○ |

The next set of questions will ask about the same symptoms. Please answer based on your experience during the past 6 weeks. If you have not experienced any of the symptoms at all, please select "Not at all" from the choices.

**During the past 6 weeks, while experiencing the symptom, how much did it bother you in terms of discomfort or pain?**

***If you have not experienced the symptom at all during the past 6 weeks, please select "Not at all."**

| | Not at all | A little bit | Somewhat | Quite a bit | Very much |
|---|---|---|---|---|---|
| Abdominal pain | ○ | ○ | ○ | ○ | ○ |
| Abnormally low temperature | ○ | ○ | ○ | ○ | ○ |
| Acid reflux | ○ | ○ | ○ | ○ | ○ |
| Afternoon or evening fevers/low-grade fevers | ○ | ○ | ○ | ○ | ○ |
| Anemia (low number of red blood cells) | ○ | ○ | ○ | ○ | ○ |
| Anxiety | ○ | ○ | ○ | ○ | ○ |
| Arrythmia (improper beating of the heart due to electrical impulse problems) | ○ | ○ | ○ | ○ | ○ |
| Bilateral neck throbbing around lymph nodes | ○ | ○ | ○ | ○ | ○ |
| Blurry vision | ○ | ○ | ○ | ○ | ○ |
| Bone aches in extremities | ○ | ○ | ○ | ○ | ○ |
| Brain fog | ○ | ○ | ○ | ○ | ○ |
| Brain pressure | ○ | ○ | ○ | ○ | ○ |
| Bulging veins | ○ | ○ | ○ | ○ | ○ |
| Burning sensations | ○ | ○ | ○ | ○ | ○ |
| Bruising of skin | ○ | ○ | ○ | ○ | ○ |
| Calf cramps | ○ | ○ | ○ | ○ | ○ |
| Change in nails (i.e. white spots, brittleness, change in moons) | ○ | ○ | ○ | ○ | ○ |
| Changes in voice | ○ | ○ | ○ | ○ | ○ |
| Changed sense of taste | ○ | ○ | ○ | ○ | ○ |
| Chills but no fever | ○ | ○ | ○ | ○ | ○ |
| Clogged ears | ○ | ○ | ○ | ○ | ○ |
| Cold burning feeling in lungs | ○ | ○ | ○ | ○ | ○ |
| Confusion | ○ | ○ | ○ | ○ | ○ |
| Congested or runny nose | ○ | ○ | ○ | ○ | ○ |
| Constant thirst | ○ | ○ | ○ | ○ | ○ |
| Costochondritis (inflammation of the cartilage that connects a rib to the breastbone) | ○ | ○ | ○ | ○ | ○ |

| | | | | | |
|---|---|---|---|---|---|
| Cough | ○ | ○ | ○ | ○ | ○ |
| Coughing up blood | ○ | ○ | ○ | ○ | ○ |
| Covid toes (tender or itchy rash or chilblains on the toes or foot) | ○ | ○ | ○ | ○ | ○ |
| Cracked or dry lips | ○ | ○ | ○ | ○ | ○ |
| Dental problems (i.e. chipped tooth, tooth loss) | ○ | ○ | ○ | ○ | ○ |
| Diarrhea | ○ | ○ | ○ | ○ | ○ |
| Difficulty concentrating or focusing | ○ | ○ | ○ | ○ | ○ |
| Difficulty sleeping | ○ | ○ | ○ | ○ | ○ |
| Difficulty speaking properly | ○ | ○ | ○ | ○ | ○ |
| Discoloration of the skin (for example: purple or blue on the hands or feet, no blistering) | ○ | ○ | ○ | ○ | ○ |
| Dizziness | ○ | ○ | ○ | ○ | ○ |
| Dry eyes | ○ | ○ | ○ | ○ | ○ |
| Dry or peeling skin | ○ | ○ | ○ | ○ | ○ |
| Dry scalp or dandruff | ○ | ○ | ○ | ○ | ○ |
| Dry throat | ○ | ○ | ○ | ○ | ○ |
| Ear pain/earache | ○ | ○ | ○ | ○ | ○ |
| Elevated thyroid hormones | ○ | ○ | ○ | ○ | ○ |
| Extreme pressure at base of head or occipital nerve | ○ | ○ | ○ | ○ | ○ |
| Eye stye or infection | ○ | ○ | ○ | ○ | ○ |
| Fatigue | ○ | ○ | ○ | ○ | ○ |
| Feeling irritable | ○ | ○ | ○ | ○ | ○ |
| Feeling of burning skin | ○ | ○ | ○ | ○ | ○ |
| Fever or chills | ○ | ○ | ○ | ○ | ○ |
| Floaters or flashes of light in vision | ○ | ○ | ○ | ○ | ○ |
| Foot pain | ○ | ○ | ○ | ○ | ○ |
| GERD (acid reflux) with excessive salivation | ○ | ○ | ○ | ○ | ○ |
| Goiter or lump in throat | ○ | ○ | ○ | ○ | ○ |
| Hair loss | ○ | ○ | ○ | ○ | ○ |
| Hand or wrist pain | ○ | ○ | ○ | ○ | ○ |
| Headache | ○ | ○ | ○ | ○ | ○ |
| Heart palpitations (heart skipping a beat or racing) | ○ | ○ | ○ | ○ | ○ |
| Heat intolerance | ○ | ○ | ○ | ○ | ○ |

| | | | | | |
|---|---|---|---|---|---|
| High blood pressure | ○ | ○ | ○ | ○ | ○ |
| Hormone imbalances | ○ | ○ | ○ | ○ | ○ |
| "Hot" blood rush | ○ | ○ | ○ | ○ | ○ |
| Inability to cry | ○ | ○ | ○ | ○ | ○ |
| Inability to exercise or be active | ○ | ○ | ○ | ○ | ○ |
| Inability to yawn | ○ | ○ | ○ | ○ | ○ |
| Internal tremors or buzzing/vibration | ○ | ○ | ○ | ○ | ○ |
| Irregular or skipped menstrual cycles | ○ | ○ | ○ | ○ | ○ |
| Jaw pain | ○ | ○ | ○ | ○ | ○ |
| Joint pain | ○ | ○ | ○ | ○ | ○ |
| Kidney issues or protein in urine | ○ | ○ | ○ | ○ | ○ |
| Kidney pain | ○ | ○ | ○ | ○ | ○ |
| Low blood oxygen | ○ | ○ | ○ | ○ | ○ |
| Low blood pressure | ○ | ○ | ○ | ○ | ○ |
| Lower back pain | ○ | ○ | ○ | ○ | ○ |
| Loss of appetite | ○ | ○ | ○ | ○ | ○ |
| Loss of hearing | ○ | ○ | ○ | ○ | ○ |
| Loss or decrease in quality of vision | ○ | ○ | ○ | ○ | ○ |
| Lump in throat/difficulty swallowing | ○ | ○ | ○ | ○ | ○ |
| Memory problems | ○ | ○ | ○ | ○ | ○ |
| Menstrual cycles that are heavier or lighter than normal | ○ | ○ | ○ | ○ | ○ |
| Mid-back pain at base of ribs | ○ | ○ | ○ | ○ | ○ |
| Mouth sores or sore tongue | ○ | ○ | ○ | ○ | ○ |
| Muscle or body aches | ○ | ○ | ○ | ○ | ○ |
| Muscle twitching | ○ | ○ | ○ | ○ | ○ |
| Nausea or vomiting | ○ | ○ | ○ | ○ | ○ |
| Neck muscle pain | ○ | ○ | ○ | ○ | ○ |
| Nerve pain | ○ | ○ | ○ | ○ | ○ |
| Nerve sensations (tingling, pins and needles, numbness) | ○ | ○ | ○ | ○ | ○ |
| Neuropathy in feet and hands (weakness, numbness, and pain) | ○ | ○ | ○ | ○ | ○ |
| New allergies | ○ | ○ | ○ | ○ | ○ |
| Night sweats | ○ | ○ | ○ | ○ | ○ |
| Nightmares | ○ | ○ | ○ | ○ | ○ |
| Painful scalp | ○ | ○ | ○ | ○ | ○ |

| | | | | | |
|---|---|---|---|---|---|
| Partial or complete loss of sense of smell | ○ | ○ | ○ | ○ | ○ |
| Partial or complete loss of sense of taste | ○ | ○ | ○ | ○ | ○ |
| Persistent chest pain or pressure | ○ | ○ | ○ | ○ | ○ |
| Personality change (drastic) | ○ | ○ | ○ | ○ | ○ |
| Petechiae (pinpoint rash) | ○ | ○ | ○ | ○ | ○ |
| Phantom smells | ○ | ○ | ○ | ○ | ○ |
| Phlegm in back of throat | ○ | ○ | ○ | ○ | ○ |
| Post-exertional malaise (worsened symptoms or flu-like symptoms after exertion) | ○ | ○ | ○ | ○ | ○ |
| Postnasal drip | ○ | ○ | ○ | ○ | ○ |
| Rash | ○ | ○ | ○ | ○ | ○ |
| Reflux or heartburn | ○ | ○ | ○ | ○ | ○ |
| Runny nose | ○ | ○ | ○ | ○ | ○ |
| Sadness | ○ | ○ | ○ | ○ | ○ |
| Seizures | ○ | ○ | ○ | ○ | ○ |
| Sharp or sudden chest pain | ○ | ○ | ○ | ○ | ○ |
| Shortness of breath or difficulty breathing | ○ | ○ | ○ | ○ | ○ |
| Shortness of breath or exhaustion from bending over | ○ | ○ | ○ | ○ | ○ |
| Sleeping more than normal | ○ | ○ | ○ | ○ | ○ |
| Sore throat | ○ | ○ | ○ | ○ | ○ |
| Spinal issues | ○ | ○ | ○ | ○ | ○ |
| Spikes in blood pressure | ○ | ○ | ○ | ○ | ○ |
| Swollen hands or feet | ○ | ○ | ○ | ○ | ○ |
| Swollen lymph nodes | ○ | ○ | ○ | ○ | ○ |
| Syncope (fainting) | ○ | ○ | ○ | ○ | ○ |
| Tachycardia (rapid heartbeat) at rest | ○ | ○ | ○ | ○ | ○ |
| Tachycardia (rapid heartbeat) after standing up | ○ | ○ | ○ | ○ | ○ |
| Thrush (white fungal infection in the mouth or throat) | ○ | ○ | ○ | ○ | ○ |
| Tinnitus or humming in ears | ○ | ○ | ○ | ○ | ○ |
| Tremors or shakiness | ○ | ○ | ○ | ○ | ○ |
| Upper back pain | ○ | ○ | ○ | ○ | ○ |
| UTI (urinary tract infection) | ○ | ○ | ○ | ○ | ○ |
| Weakened neck | ○ | ○ | ○ | ○ | ○ |

| | | | | | |
|---|---|---|---|---|---|
| Weight gain | ○ | ○ | ○ | ○ | ○ |

**During the past 6 weeks, ow much did the symptom impair your ability to work compared to pre-COVID?**

***If you have not experienced the symptom at all during the past 6 weeks, please select "Not at all."**

| | Not at all | A little bit | Somewhat | Quite a bit | Very much |
|---|---|---|---|---|---|
| Abdominal pain | ○ | ○ | ○ | ○ | ○ |
| Abnormally low temperature | ○ | ○ | ○ | ○ | ○ |
| Acid reflux | ○ | ○ | ○ | ○ | ○ |
| Afternoon or evening fevers/low-grade fevers | ○ | ○ | ○ | ○ | ○ |
| Anemia (low number of red blood cells) | ○ | ○ | ○ | ○ | ○ |
| Anxiety | ○ | ○ | ○ | ○ | ○ |
| Arrythmia (improper beating of the heart due to electrical impulse problems) | ○ | ○ | ○ | ○ | ○ |
| Bilateral neck throbbing around lymph nodes | ○ | ○ | ○ | ○ | ○ |
| Blurry vision | ○ | ○ | ○ | ○ | ○ |
| Bone aches in extremities | ○ | ○ | ○ | ○ | ○ |
| Brain fog | ○ | ○ | ○ | ○ | ○ |
| Brain pressure | ○ | ○ | ○ | ○ | ○ |
| Bulging veins | ○ | ○ | ○ | ○ | ○ |
| Burning sensations | ○ | ○ | ○ | ○ | ○ |
| Bruising of skin | ○ | ○ | ○ | ○ | ○ |
| Calf cramps | ○ | ○ | ○ | ○ | ○ |
| Change in nails (i.e. white spots, brittleness, change in moons) | ○ | ○ | ○ | ○ | ○ |
| Changes in voice | ○ | ○ | ○ | ○ | ○ |
| Changed sense of taste | ○ | ○ | ○ | ○ | ○ |
| Chills but no fever | ○ | ○ | ○ | ○ | ○ |
| Clogged ears | ○ | ○ | ○ | ○ | ○ |
| Cold burning feeling in lungs | ○ | ○ | ○ | ○ | ○ |
| Confusion | ○ | ○ | ○ | ○ | ○ |
| Congested or runny nose | ○ | ○ | ○ | ○ | ○ |
| Constant thirst | ○ | ○ | ○ | ○ | ○ |
| Costochondritis (inflammation of the cartilage that connects a rib to the breastbone) | ○ | ○ | ○ | ○ | ○ |

| | | | | | |
|---|---|---|---|---|---|
| Cough | ○ | ○ | ○ | ○ | ○ |
| Coughing up blood | ○ | ○ | ○ | ○ | ○ |
| Covid toes (tender or itchy rash or chilblains on the toes or foot) | ○ | ○ | ○ | ○ | ○ |
| Cracked or dry lips | ○ | ○ | ○ | ○ | ○ |
| Dental problems (i.e. chipped tooth, tooth loss) | ○ | ○ | ○ | ○ | ○ |
| Diarrhea | ○ | ○ | ○ | ○ | ○ |
| Difficulty concentrating or focusing | ○ | ○ | ○ | ○ | ○ |
| Difficulty sleeping | ○ | ○ | ○ | ○ | ○ |
| Difficulty speaking properly | ○ | ○ | ○ | ○ | ○ |
| Discoloration of the skin (for example: purple or blue on the hands or feet, no blistering) | ○ | ○ | ○ | ○ | ○ |
| Dizziness | ○ | ○ | ○ | ○ | ○ |
| Dry eyes | ○ | ○ | ○ | ○ | ○ |
| Dry or peeling skin | ○ | ○ | ○ | ○ | ○ |
| Dry scalp or dandruff | ○ | ○ | ○ | ○ | ○ |
| Dry throat | ○ | ○ | ○ | ○ | ○ |
| Ear pain/earache | ○ | ○ | ○ | ○ | ○ |
| Elevated thyroid hormones | ○ | ○ | ○ | ○ | ○ |
| Extreme pressure at base of head or occipital nerve | ○ | ○ | ○ | ○ | ○ |
| Eye stye or infection | ○ | ○ | ○ | ○ | ○ |
| Fatigue | ○ | ○ | ○ | ○ | ○ |
| Feeling irritable | ○ | ○ | ○ | ○ | ○ |
| Feeling of burning skin | ○ | ○ | ○ | ○ | ○ |
| Fever or chills | ○ | ○ | ○ | ○ | ○ |
| Floaters or flashes of light in vision | ○ | ○ | ○ | ○ | ○ |
| Foot pain | ○ | ○ | ○ | ○ | ○ |
| GERD (acid reflux) with excessive salivation | ○ | ○ | ○ | ○ | ○ |
| Goiter or lump in throat | ○ | ○ | ○ | ○ | ○ |
| Hair loss | ○ | ○ | ○ | ○ | ○ |
| Hand or wrist pain | ○ | ○ | ○ | ○ | ○ |
| Headache | ○ | ○ | ○ | ○ | ○ |
| Heart palpitations (heart skipping a beat or racing) | ○ | ○ | ○ | ○ | ○ |
| Heat intolerance | ○ | ○ | ○ | ○ | ○ |

| | | | | | |
|---|---|---|---|---|---|
| High blood pressure | ○ | ○ | ○ | ○ | ○ |
| Hormone imbalances | ○ | ○ | ○ | ○ | ○ |
| "Hot" blood rush | ○ | ○ | ○ | ○ | ○ |
| Inability to cry | ○ | ○ | ○ | ○ | ○ |
| Inability to exercise or be active | ○ | ○ | ○ | ○ | ○ |
| Inability to yawn | ○ | ○ | ○ | ○ | ○ |
| Internal tremors or buzzing/vibration | ○ | ○ | ○ | ○ | ○ |
| Irregular or skipped menstrual cycles | ○ | ○ | ○ | ○ | ○ |
| Jaw pain | ○ | ○ | ○ | ○ | ○ |
| Joint pain | ○ | ○ | ○ | ○ | ○ |
| Kidney issues or protein in urine | ○ | ○ | ○ | ○ | ○ |
| Kidney pain | ○ | ○ | ○ | ○ | ○ |
| Low blood oxygen | ○ | ○ | ○ | ○ | ○ |
| Low blood pressure | ○ | ○ | ○ | ○ | ○ |
| Lower back pain | ○ | ○ | ○ | ○ | ○ |
| Loss of appetite | ○ | ○ | ○ | ○ | ○ |
| Loss of hearing | ○ | ○ | ○ | ○ | ○ |
| Loss or decrease in quality of vision | ○ | ○ | ○ | ○ | ○ |
| Lump in throat/difficulty swallowing | ○ | ○ | ○ | ○ | ○ |
| Memory problems | ○ | ○ | ○ | ○ | ○ |
| Menstrual cycles that are heavier or lighter than normal | ○ | ○ | ○ | ○ | ○ |
| Mid-back pain at base of ribs | ○ | ○ | ○ | ○ | ○ |
| Mouth sores or sore tongue | ○ | ○ | ○ | ○ | ○ |
| Muscle or body aches | ○ | ○ | ○ | ○ | ○ |
| Muscle twitching | ○ | ○ | ○ | ○ | ○ |
| Nausea or vomiting | ○ | ○ | ○ | ○ | ○ |
| Neck muscle pain | ○ | ○ | ○ | ○ | ○ |
| Nerve pain | ○ | ○ | ○ | ○ | ○ |
| Nerve sensations (tingling, pins and needles, numbness) | ○ | ○ | ○ | ○ | ○ |
| Neuropathy in feet and hands (weakness, numbness, and pain) | ○ | ○ | ○ | ○ | ○ |
| New allergies | ○ | ○ | ○ | ○ | ○ |
| Night sweats | ○ | ○ | ○ | ○ | ○ |
| Nightmares | ○ | ○ | ○ | ○ | ○ |
| Painful scalp | ○ | ○ | ○ | ○ | ○ |

| | | | | | |
|---|---|---|---|---|---|
| Partial or complete loss of sense of smell | ◯ | ◯ | ◯ | ◯ | ◯ |
| Partial or complete loss of sense of taste | ◯ | ◯ | ◯ | ◯ | ◯ |
| Persistent chest pain or pressure | ◯ | ◯ | ◯ | ◯ | ◯ |
| Personality change (drastic) | ◯ | ◯ | ◯ | ◯ | ◯ |
| Petechiae (pinpoint rash) | ◯ | ◯ | ◯ | ◯ | ◯ |
| Phantom smells | ◯ | ◯ | ◯ | ◯ | ◯ |
| Phlegm in back of throat | ◯ | ◯ | ◯ | ◯ | ◯ |
| Post-exertional malaise (worsened symptoms or flu-like symptoms after exertion) | ◯ | ◯ | ◯ | ◯ | ◯ |
| Postnasal drip | ◯ | ◯ | ◯ | ◯ | ◯ |
| Rash | ◯ | ◯ | ◯ | ◯ | ◯ |
| Reflux or heartburn | ◯ | ◯ | ◯ | ◯ | ◯ |
| Runny nose | ◯ | ◯ | ◯ | ◯ | ◯ |
| Sadness | ◯ | ◯ | ◯ | ◯ | ◯ |
| Seizures | ◯ | ◯ | ◯ | ◯ | ◯ |
| Sharp or sudden chest pain | ◯ | ◯ | ◯ | ◯ | ◯ |
| Shortness of breath or difficulty breathing | ◯ | ◯ | ◯ | ◯ | ◯ |
| Shortness of breath or exhaustion from bending over | ◯ | ◯ | ◯ | ◯ | ◯ |
| Sleeping more than normal | ◯ | ◯ | ◯ | ◯ | ◯ |
| Sore throat | ◯ | ◯ | ◯ | ◯ | ◯ |
| Spinal issues | ◯ | ◯ | ◯ | ◯ | ◯ |
| Spikes in blood pressure | ◯ | ◯ | ◯ | ◯ | ◯ |
| Swollen hands or feet | ◯ | ◯ | ◯ | ◯ | ◯ |
| Swollen lymph nodes | ◯ | ◯ | ◯ | ◯ | ◯ |
| Syncope (fainting) | ◯ | ◯ | ◯ | ◯ | ◯ |
| Tachycardia (rapid heartbeat) at rest | ◯ | ◯ | ◯ | ◯ | ◯ |
| Tachycardia (rapid heartbeat) after standing up | ◯ | ◯ | ◯ | ◯ | ◯ |
| Thrush (white fungal infection in the mouth or throat) | ◯ | ◯ | ◯ | ◯ | ◯ |
| Tinnitus or humming in ears | ◯ | ◯ | ◯ | ◯ | ◯ |
| Tremors or shakiness | ◯ | ◯ | ◯ | ◯ | ◯ |
| Upper back pain | ◯ | ◯ | ◯ | ◯ | ◯ |
| UTI (urinary tract infection) | ◯ | ◯ | ◯ | ◯ | ◯ |
| Weakened neck | ◯ | ◯ | ◯ | ◯ | ◯ |

| | | | | | |
|---|---|---|---|---|---|
| Weight gain | ○ | ○ | ○ | ○ | ○ |

**During the past 6 weeks, how much did the symptom impair your social or family functioning compared to pre-COVID?**

***If you have not experienced the symptom at all during the past 6 weeks, please select "Not at all."**

| | Not at all | A little bit | Somewhat | Quite a bit | Very much |
|---|---|---|---|---|---|
| Abdominal pain | ○ | ○ | ○ | ○ | ○ |
| Abnormally low temperature | ○ | ○ | ○ | ○ | ○ |
| Acid reflux | ○ | ○ | ○ | ○ | ○ |
| Afternoon or evening fevers/low-grade fevers | ○ | ○ | ○ | ○ | ○ |
| Anemia (low number of red blood cells) | ○ | ○ | ○ | ○ | ○ |
| Anxiety | ○ | ○ | ○ | ○ | ○ |
| Arrythmia (improper beating of the heart due to electrical impulse problems) | ○ | ○ | ○ | ○ | ○ |
| Bilateral neck throbbing around lymph nodes | ○ | ○ | ○ | ○ | ○ |
| Blurry vision | ○ | ○ | ○ | ○ | ○ |
| Bone aches in extremities | ○ | ○ | ○ | ○ | ○ |
| Brain fog | ○ | ○ | ○ | ○ | ○ |
| Brain pressure | ○ | ○ | ○ | ○ | ○ |
| Bulging veins | ○ | ○ | ○ | ○ | ○ |
| Burning sensations | ○ | ○ | ○ | ○ | ○ |
| Bruising of skin | ○ | ○ | ○ | ○ | ○ |
| Calf cramps | ○ | ○ | ○ | ○ | ○ |
| Change in nails (i.e. white spots, brittleness, change in moons) | ○ | ○ | ○ | ○ | ○ |
| Changes in voice | ○ | ○ | ○ | ○ | ○ |
| Changed sense of taste | ○ | ○ | ○ | ○ | ○ |
| Chills but no fever | ○ | ○ | ○ | ○ | ○ |
| Clogged ears | ○ | ○ | ○ | ○ | ○ |
| Cold burning feeling in lungs | ○ | ○ | ○ | ○ | ○ |
| Confusion | ○ | ○ | ○ | ○ | ○ |
| Congested or runny nose | ○ | ○ | ○ | ○ | ○ |
| Constant thirst | ○ | ○ | ○ | ○ | ○ |
| Costochondritis (inflammation of the cartilage that connects a rib to the breastbone) | ○ | ○ | ○ | ○ | ○ |

| | | | | | |
|---|---|---|---|---|---|
| Cough | ○ | ○ | ○ | ○ | ○ |
| Coughing up blood | ○ | ○ | ○ | ○ | ○ |
| Covid toes (tender or itchy rash or chilblains on the toes or foot) | ○ | ○ | ○ | ○ | ○ |
| Cracked or dry lips | ○ | ○ | ○ | ○ | ○ |
| Dental problems (i.e. chipped tooth, tooth loss) | ○ | ○ | ○ | ○ | ○ |
| Diarrhea | ○ | ○ | ○ | ○ | ○ |
| Difficulty concentrating or focusing | ○ | ○ | ○ | ○ | ○ |
| Difficulty sleeping | ○ | ○ | ○ | ○ | ○ |
| Difficulty speaking properly | ○ | ○ | ○ | ○ | ○ |
| Discoloration of the skin (for example: purple or blue on the hands or feet, no blistering) | ○ | ○ | ○ | ○ | ○ |
| Dizziness | ○ | ○ | ○ | ○ | ○ |
| Dry eyes | ○ | ○ | ○ | ○ | ○ |
| Dry or peeling skin | ○ | ○ | ○ | ○ | ○ |
| Dry scalp or dandruff | ○ | ○ | ○ | ○ | ○ |
| Dry throat | ○ | ○ | ○ | ○ | ○ |
| Ear pain/earache | ○ | ○ | ○ | ○ | ○ |
| Elevated thyroid hormones | ○ | ○ | ○ | ○ | ○ |
| Extreme pressure at base of head or occipital nerve | ○ | ○ | ○ | ○ | ○ |
| Eye stye or infection | ○ | ○ | ○ | ○ | ○ |
| Fatigue | ○ | ○ | ○ | ○ | ○ |
| Feeling irritable | ○ | ○ | ○ | ○ | ○ |
| Feeling of burning skin | ○ | ○ | ○ | ○ | ○ |
| Fever or chills | ○ | ○ | ○ | ○ | ○ |
| Floaters or flashes of light in vision | ○ | ○ | ○ | ○ | ○ |
| Foot pain | ○ | ○ | ○ | ○ | ○ |
| GERD (acid reflux) with excessive salivation | ○ | ○ | ○ | ○ | ○ |
| Goiter or lump in throat | ○ | ○ | ○ | ○ | ○ |
| Hair loss | ○ | ○ | ○ | ○ | ○ |
| Hand or wrist pain | ○ | ○ | ○ | ○ | ○ |
| Headache | ○ | ○ | ○ | ○ | ○ |
| Heart palpitations (heart skipping a beat or racing) | ○ | ○ | ○ | ○ | ○ |
| Heat intolerance | ○ | ○ | ○ | ○ | ○ |

| | | | | | |
|---|---|---|---|---|---|
| High blood pressure | ○ | ○ | ○ | ○ | ○ |
| Hormone imbalances | ○ | ○ | ○ | ○ | ○ |
| "Hot" blood rush | ○ | ○ | ○ | ○ | ○ |
| Inability to cry | ○ | ○ | ○ | ○ | ○ |
| Inability to exercise or be active | ○ | ○ | ○ | ○ | ○ |
| Inability to yawn | ○ | ○ | ○ | ○ | ○ |
| Internal tremors or buzzing/vibration | ○ | ○ | ○ | ○ | ○ |
| Irregular or skipped menstrual cycles | ○ | ○ | ○ | ○ | ○ |
| Jaw pain | ○ | ○ | ○ | ○ | ○ |
| Joint pain | ○ | ○ | ○ | ○ | ○ |
| Kidney issues or protein in urine | ○ | ○ | ○ | ○ | ○ |
| Kidney pain | ○ | ○ | ○ | ○ | ○ |
| Low blood oxygen | ○ | ○ | ○ | ○ | ○ |
| Low blood pressure | ○ | ○ | ○ | ○ | ○ |
| Lower back pain | ○ | ○ | ○ | ○ | ○ |
| Loss of appetite | ○ | ○ | ○ | ○ | ○ |
| Loss of hearing | ○ | ○ | ○ | ○ | ○ |
| Loss or decrease in quality of vision | ○ | ○ | ○ | ○ | ○ |
| Lump in throat/difficulty swallowing | ○ | ○ | ○ | ○ | ○ |
| Memory problems | ○ | ○ | ○ | ○ | ○ |
| Menstrual cycles that are heavier or lighter than normal | ○ | ○ | ○ | ○ | ○ |
| Mid-back pain at base of ribs | ○ | ○ | ○ | ○ | ○ |
| Mouth sores or sore tongue | ○ | ○ | ○ | ○ | ○ |
| Muscle or body aches | ○ | ○ | ○ | ○ | ○ |
| Muscle twitching | ○ | ○ | ○ | ○ | ○ |
| Nausea or vomiting | ○ | ○ | ○ | ○ | ○ |
| Neck muscle pain | ○ | ○ | ○ | ○ | ○ |
| Nerve pain | ○ | ○ | ○ | ○ | ○ |
| Nerve sensations (tingling, pins and needles, numbness) | ○ | ○ | ○ | ○ | ○ |
| Neuropathy in feet and hands (weakness, numbness, and pain) | ○ | ○ | ○ | ○ | ○ |
| New allergies | ○ | ○ | ○ | ○ | ○ |
| Night sweats | ○ | ○ | ○ | ○ | ○ |
| Nightmares | ○ | ○ | ○ | ○ | ○ |
| Painful scalp | ○ | ○ | ○ | ○ | ○ |

| | | | | | |
|---|---|---|---|---|---|
| Partial or complete loss of sense of smell | ○ | ○ | ○ | ○ | ○ |
| Partial or complete loss of sense of taste | ○ | ○ | ○ | ○ | ○ |
| Persistent chest pain or pressure | ○ | ○ | ○ | ○ | ○ |
| Personality change (drastic) | ○ | ○ | ○ | ○ | ○ |
| Petechiae (pinpoint rash) | ○ | ○ | ○ | ○ | ○ |
| Phantom smells | ○ | ○ | ○ | ○ | ○ |
| Phlegm in back of throat | ○ | ○ | ○ | ○ | ○ |
| Post-exertional malaise (worsened symptoms or flu-like symptoms after exertion) | ○ | ○ | ○ | ○ | ○ |
| Postnasal drip | ○ | ○ | ○ | ○ | ○ |
| Rash | ○ | ○ | ○ | ○ | ○ |
| Reflux or heartburn | ○ | ○ | ○ | ○ | ○ |
| Runny nose | ○ | ○ | ○ | ○ | ○ |
| Sadness | ○ | ○ | ○ | ○ | ○ |
| Seizures | ○ | ○ | ○ | ○ | ○ |
| Sharp or sudden chest pain | ○ | ○ | ○ | ○ | ○ |
| Shortness of breath or difficulty breathing | ○ | ○ | ○ | ○ | ○ |
| Shortness of breath or exhaustion from bending over | ○ | ○ | ○ | ○ | ○ |
| Sleeping more than normal | ○ | ○ | ○ | ○ | ○ |
| Sore throat | ○ | ○ | ○ | ○ | ○ |
| Spinal issues | ○ | ○ | ○ | ○ | ○ |
| Spikes in blood pressure | ○ | ○ | ○ | ○ | ○ |
| Swollen hands or feet | ○ | ○ | ○ | ○ | ○ |
| Swollen lymph nodes | ○ | ○ | ○ | ○ | ○ |
| Syncope (fainting) | ○ | ○ | ○ | ○ | ○ |
| Tachycardia (rapid heartbeat) at rest | ○ | ○ | ○ | ○ | ○ |
| Tachycardia (rapid heartbeat) after standing up | ○ | ○ | ○ | ○ | ○ |
| Thrush (white fungal infection in the mouth or throat) | ○ | ○ | ○ | ○ | ○ |
| Tinnitus or humming in ears | ○ | ○ | ○ | ○ | ○ |
| Tremors or shakiness | ○ | ○ | ○ | ○ | ○ |
| Upper back pain | ○ | ○ | ○ | ○ | ○ |
| UTI (urinary tract infection) | ○ | ○ | ○ | ○ | ○ |
| Weakened neck | ○ | ○ | ○ | ○ | ○ |

| | | | | | |
|---|---|---|---|---|---|
| Weight gain | ○ | ○ | ○ | ○ | ○ |

Are there any symptoms not listed above that you have experienced within the past 6 weeks?

*Please list all if so

#### During the past 6 weeks

| | 76-100% of health before COVID-19 | 51-75% of health before COVID-19 | 26-50% of health before COVID-19 | 0-25% of health before COVID-19 |
|---|---|---|---|---|
| On your best day, would you say that you are at | ○ | ○ | ○ | ○ |
| On your worst day, would you say that you are at | ○ | ○ | ○ | ○ |

Compared to your health before you received the first dose of your vaccine on [vaccine_arm_1][v_v_vax_date], would you say that your health is currently

○ Better
○ Worse
○ The same
○ Don't know

Please, in the space below, share any information about your experience with COVID-19 or the vaccine that we might have missed.

__________________________________________
